# Targeting multicopy prophage genes for the differential diagnosis of Lyme disease

**DOI:** 10.1101/2020.12.02.20241687

**Authors:** Jinyu Shan, Ying Jia, Louis Teulières, Faizal Patel, Martha R.J. Clokie

## Abstract

The successful treatment of Lyme disease (LD) requires an accurate diagnostic test; however, most tests are insensitive and unspecific. To overcome these challenges, we developed and validated an internally-controlled quantitative PCR (Ter-qPCR) that targets the multicopy terminase large subunit (*terL*) gene encoded by prophages that are only found in LD-causing bacteria. The *terL* protein helps phages pack their DNA. Strikingly, the detection limit of the Ter-qPCR was analytically estimated to be 22 copies and one bacterial cell in bacteria spiked blood. Furthermore, significant quantitative differences in terms of the amount of *terL* detected in healthy individuals and patients with either early or late disease. Together, the data suggests that the prophage-targeting PCR has significant power to provide a differential diagnosis for LD. Prophage encoded markers are prevalent in many other pathogenic bacteria rendering this approach highly applicable to bacterial identification in general, potentially revolutionising the detection of disease.

## Introduction

Lyme disease (LD) is the most common tick-born disease with more than 300, 000 cases in the US and 100, 000 cases in Europe reported annually^1,2^. LD is caused by a group of bacteria classified together as the *Borrelia burgdorferi* sensu lato (s.l.) complex, that comprises a clade of more than 20 species including *B. burgdorferi* sensu stricto (s.s.) which dominates in US, and *B. garinii* and *B. afzelii* which are prevalent in Europe and Asia. The LD-causing bacteria are generally transmitted to humans after they are bitten by ticks of the *Ixodes* family infected with LD causing *Borrelia*. However, recent reports have raised concerns over *Borrelia* transmission through blood transfusion based on observations that *Borrelia* can survive and circulate in the human bloodstream^3^.

Currently, LD diagnosis is based on the overt clinical manifestation of disease in the form of erythema migrans (EM) skin lesions, commonly known as a ‘bull’s-eye’ rash^4^ and a history of tick exposure^5^. Although EM lesions occur in 70 to 80% of infected individuals^5,6^, only a third of these patients develop the classic ‘bull’s-eye’ rash^7^, and many other types of skin lesions can occur which are easily confused with EM^4,8,9^. In addition to the EM uncertainty, other common symptoms of LD such as fatigue, muscle pain, headache, and perceived cognitive dysfunction largely overlap with an array of other diseases, including other tick-borne diseases. One such example is Relapsing Fever (RF), which is caused by close relatives of the LD-causing bacteria, such as *Borrelia miyamotoi* ^9,10^. The two *Borrelia* ‘groups’ responsible for LD and RF have caused great concern and clinical confusion, as they are morphologically similar and present with almost indistinguishable clinical symptoms^11^. Despite this, they respond to different antibiotics and treatment regimens^12,13^. Another example of confusion surrounding LD is the co-infection caused by *Bartonella* spp. This genus of bacteria is emerging as an increasingly common human infection^14^. Much of the controversy surrounding LD and co-infections with *Bartonella* and/or *B. miyamotoi* is due to the lack of a reliable and sensitive diagnostic method to detect and distinguish between the three groups of bacteria, the LD and RF causing *Borrelia and Bartonella*^15^. Therefore, laboratory tests to determine and distinguish between LD and co-infections play a vital role in the correct diagnosis and consequent treatment with different antibiotics.

Scientists have faced several challenges with LD detection including patients presenting with a delayed antibody response and a low number of *Borrelia* cells typically found in human clinical samples^16-19^. Serological tests for LD are also hindered by the cross-reactivity present between LD and RF^16,18,20^. Although it is particularly difficult to diagnose LD early, it is critical, as it is far easier to treat the disease when it is detected at an early stage^19,21,22 4,6,23,24^. Bacteria-targeting approaches, such as polymerase chain reaction (PCR) detecting the *Borrelia chromosomal* DNA, can potentially identify early LD but is relatively insensitive detecting only between 30-50% of positive cases, and is therefore deemed to have little clinical utility ^15,17,19^. The reasons behind the poor sensitivity of the current PCR methods in Lyme diagnosis are twofold; first, the current PCRs target *Borrelia* genomic DNA regions that have only one copy in each bacterium, such as the bacterial 16S rRNA gene, *RecA* gene, and the 5S-23S intergenic regions^15,16,25-27^. Second, at least some *Borrelia* species are ‘tissue-bound’ and are only transiently found circulating in the blood ^28^.

In response to these diagnostic challenges, we adopted a novel approach, taking advantage of the fact that most pathogenic bacteria carry multiple complete or partial prophages (phages associated with bacteria)^29^. These prophage sequences can form the bases of a template from which quantitative PCR (qPCR) primers and probes can be designed. It is known that *Borrelia* carry a large number of linear and circular plasmids (comprising between 33-40% of the *Borrelia* genome), among which the cp26 and cp32, and the lp54 linear plasmid, are evolutionarily stable^30^. Of these paralogous plasmids, cp32 has been experimentally determined to be a *Borrelia burgdorferi* prophage thus it is highly likely that many of its homologues are also prophages^30-34^.

Each *Borrelia* species has a distinct amount of species specific variation in its prophage sequences; thus these prophages can be used as a proxy to identify the bacteria because of the tight correlation between them and the exact prophages found in each *Borrelia* host. As there are multiple prophages per *Borrelia* cell, the detectable signal is higher for prophages than bacteria. Furthermore, evidence suggests that *Borrelia* prophages can be released outside the *Borrelia* cells following encounters with stressors such as antibiotics or exposure to macrophages^31-33,35,36^. In this study, we confirmed that *Borrelia* prophages can escape from the bacterial host cell in a spontaneous manner. Taking advantage of the multicopy and free movement of *Borrelia* prophages, the approach to target prophages instead of bacteria will bypass the cryptic and tissue-bound feature that typifies human *Borrelia* infections^28^. Thus, we have a greater chance of detecting the prophages in blood even when the bacteria may not be present or present in extremely low numbers. In this sense, prophages are somewhat analogous to *Borrelia* ‘footprints’.

Another challenge in detecting bacteria from blood samples is the successful extraction of DNA that forms the PCR target. It is well-known that DNA extraction from blood samples plays a pivotal role in the success of PCR diagnosis of bacterial infections^37,38^. It is also true that PCR assays may fail to amplify if bacteria are sparse, or if an inappropriate DNA extraction method was adopted that favoured an overwhelming dominance of human DNA in the final DNA product^38-40^. To find the most suitable DNA extraction method, three blood DNA extraction methods were applied to blood samples spiked with serial dilutions of *Borrelia* cells and assessed by qPCR. We determined the best DNA extraction method by assessing each sample in order to determine the highest detectable phage copy number from the *Borrelia* spiked blood.

In this paper we have demonstrated for the first time in *Borrelia*-related diagnostics that it is possible to overcome the sensitivity challenges associated with LD detection. We highlight the enormous potential of our test to discriminate between healthy volunteers, early LD, and late LD patients. We present data from a systematic and comprehensive study that validates the use of the multicopy phage terminase large subunit (*terL*) gene as a molecular marker for the detection of *Borrelia* species. The analytical performance of the *terL*-targeting qPCR (referred to as Ter-qPCR) was thoroughly evaluated, and the test was shown to be able to detect one single *Borrelia* cell from blood samples. The diagnostic potential was evaluated using a set of blood and serum samples collected from healthy volunteers and individuals who were clinically diagnosed with LD.

In summary, we demonstrate that a quantitative phage-based PCR has the potential to revolutionise the differential diagnosis of LD from blood samples. This approach of looking for induced and specific phages may be useful for a plethora of blood-borne bacterial pathogens that cause sepsis, such as *Staphylococcus aureus, Escherichia coli*, and *Pseudomonas aeruginosa*41.

## Results and Discussion

### Multicopy, *terL* genes are widespread within *Borrelia* species

To determine which prophage gene to use as a marker the *B. burgdorferi* B31 genome was examined and shown to carry the multicopy *terL* gene (NC_000948.1). This gene encodes for the terL protein which is responsible for packing phage genomes and is essential for phage survival^42^. The gene was found to be present on seven of the circular plasmids from the cp32 series and on three linear plasmids within B31 genome (Supplementary information 1). Blastn analysis revealed that the *terL* homologs are widespread in LD and RF *Borrelia* spp., including *B. miyamotoi*. As summarised in Table 1, the *terL* homologs present in LD *Borrelia* species are mainly located on the cp32 plasmids with E values of zero and query cover of 100%; thus, primer design specific to species was carried out. There are 13 versions of terL-bearing cp32 plasmids (cp32-1 to cp32-13), all of which are present in the *B. burgdorferi* spp. In contrast, *B. afzelii* and *B. garinii* encode eight and four such cp32 plasmids, respectively.

**Table 1.** The prevalence of the *terL* homologs among plasmids residing *Borrelia* species causing Lyme disease (smiley face and ‘×’ to denote presence and absence of the *terL* homologs, respectively)

| Plasmid | *B. burgdorferi* | *B. afzelii* | *B. garinii* | *B. bissettii* | *B. mayonii* | *B. bavariensis* | *B. valaisiana* | *B. finlandensis* | *B. spielmanii* |
| --- | --- | --- | --- | --- | --- | --- | --- | --- | --- |
| cp32-1 | ☺ | ☺ | x | x | ☺ | ☺ | x | x | x |
| cp32-2 | ☺ | x | x | x | x | x | x | x | x |
| cp32-3 | ☺ | ☺ | ☺ | ☺ | ☺ | ☺ | x | ☺ | x |
| cp32-4 | ☺ | ☺ | x | ☺ | ☺ | x | x | ☺ | x |
| cp32-5 | ☺ | ☺ | ☺ | ☺ | x | ☺ | ☺ | x | x |
| cp32-6 | ☺ | x | ☺ | ☺ | ☺ | x | x | x | x |
| cp32-7 | ☺ | ☺ | x | ☺ | x | ☺ | x | ☺ | x |
| cp32-8 | ☺ | x | x | x | x | x | x | x | x |
| cp32-9 | ☺ | ☺ | x | x | x | x | x | x | x |
| cp32-10 | ☺ | x | ☺ | x | x | x | ☺ | x | x |
| cp32-11 | ☺ | ☺ | x | ☺ | x | x | x | x | x |
| cp32-12 | ☺ | ☺ | x | x | x | x | x | ☺ | x |
| cp32-13 | ☺ | x | x | x | ☺ | x | x | x | x |
| cp32-2-7<sup>a</sup> | x | x | x | x | x | x | ☺ | x | x |
| cp32-5+1<sup>b</sup> | ☺ | x | x | x | x | x | x | x | x |
| cp32-5-1<sup>c</sup> | ☺ | x | x | x | x | x | x | x | x |
| cp32-3+10<sup>d</sup> | ☺ | x | x | x | x | x | x | x | x |
| cp30 | x | ☺ | x | x | x | x | x | x | x |
| lp17 | ☺ | x | x | x | x | x | x | x | x |
| cp18 | ☺ | x | x | x | x | x | x | x | x |
| lp25 | x | x | x | x | x | ☺ | x | x | x |
| lp28-4 | x | x | x | x | x | ☺ | x | x | x |
| lp54 | ☺ | x | x | x | x | x | ☺ | ☺ | ☺ |
| lp56 | ☺ | x | x | x | x | x | x | x | x |
| Total hits | 147 | 25 | 14 | 8 | 8 | 6 | 5 | 5 | 1 |

- cp32-2-7 refers to cp32-2 and cp32-7 as both plasmids have the same compatibility<sup>30</sup>. - cp32-1+5 refers to a fusion plasmid discovered from *B. burgdorferi* JD1 that is made up of two different full-length cp32-1 and cp32-5 fused together to form a circular plasmid<sup>36</sup>. - cp32-1-5 is another version of cp32-1-5 referring to the fusion of cp32-1 and cp32-5 into a single circular replicon<sup>31</sup>. - cp32-3+10 refers to a fusion plasmid discovered from *B. burgdorferi* B31 that is made up of two different full-length cp32-3 and cp32-10 fused together to form a circular plasmid<sup>36,83</sup>.

*TerL* homologs are also found in RF and *B. miyamotoi* species (E values ranging from 5e-164 to 0.03 and a query cover ranging from 90% to 5%). Thus, the phage terL gene appears to be a useful marker; indeed, it has previously been used as a marker to reveal the evolutionary relationships within prophages of the environmental *Paraburkholderia* species and lytic phages of *Edwardsiella ictaluri*, the causative agent of enteric septicaemia of catfish^43,44^. In summary, the prevalence, amount of variability in the sequence, and multicopy nature of *terL* suggests that it could be a suitable marker to indicate *Borrelia* presence. Therefore, we predicted that a qPCR targeting *terL* homologs will detect *Borrelia* species to the strain level and distinguish LD *Borrelia* species from RF species and *B. miyamotoi*.

### Phylogenetic analysis of the *terL* gene

To determine the potential of the *terL* gene as a marker for specific *Borrelia* species, phylogenetic analyses were carried out using neighbour-joining (NJ) (Fig. 1) and likelihood (ML) methods (Supplementary Fig. 1). Both trees were concordant with each other and demonstrated three well-defined clades of LD, RF and *B. miyamotoi*, indicating that the *terL* gene is evolutionary stable and offers resolution at the *Borrelia* species level. As shown in Fig. 1, the *terL* gene resolves *Borrelia* into genospecies, revealing an independent sub-group of LD *Borrelia* species that is well-separated from the other RF *Borrelia* group (including *B. miyamotoi*) with statistically a significant bootstrap value.

**Fig. 1.**
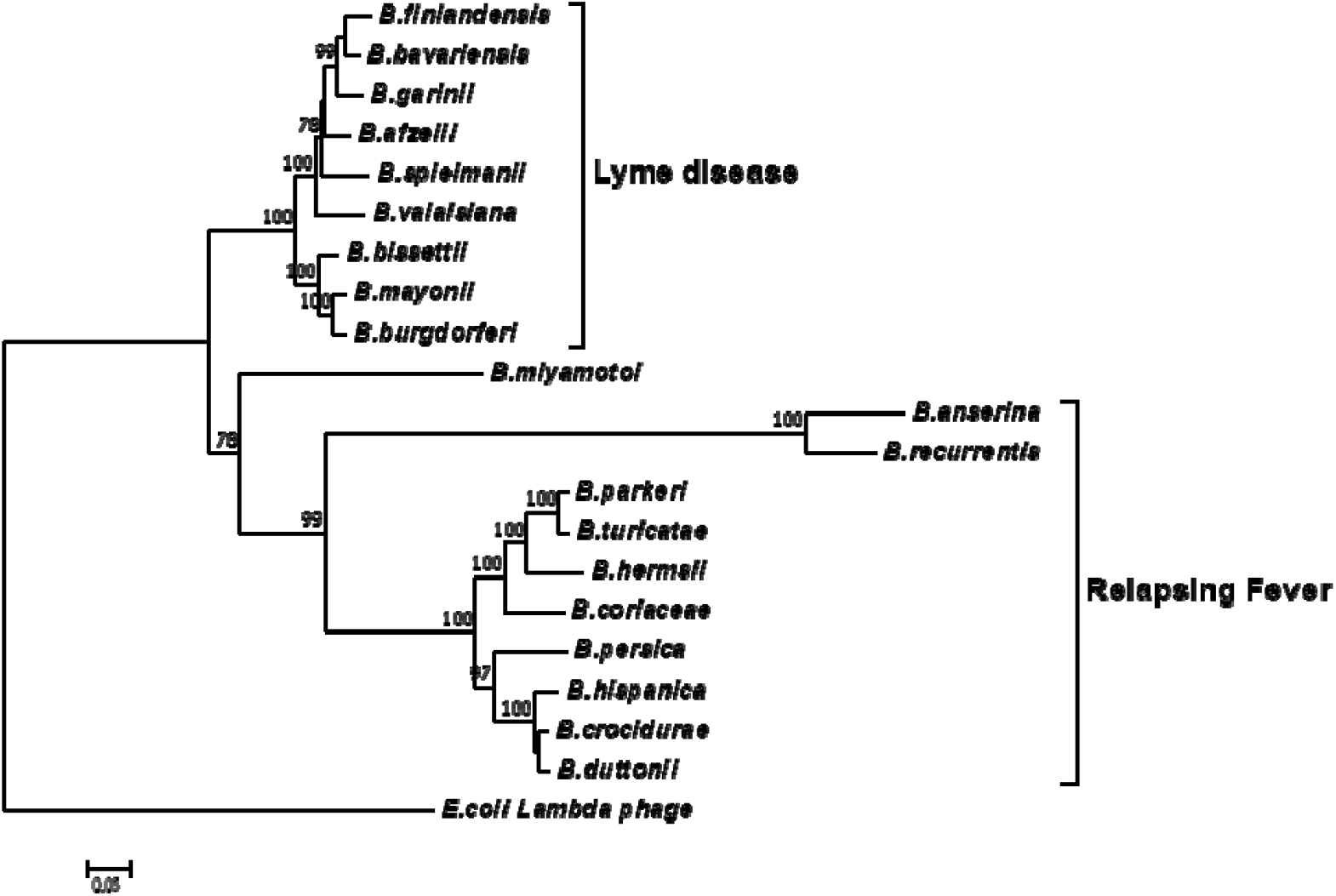
A phylogenetic tree constructed based on the *terL* nucleotide sequence. The LD and RF *Borrelia* species and *B. miyamotoi* are separated into three well-supported clades. The Molecular Evolutionary Genetics Analysis (MEGA) package version 7 was used, and the tree was constructed via the neighbour-joining method. Scale bar represents the units of the number of base substitutions per site. Support for the clades was estimated via bootstrap analysis in MEGA with 3,000 replicates, values are indicated at the nodes (only values greater than 75 are displayed). The *terL* sequence from phage lambda was used as an outgroup to root the tree.

Encouragingly, and of significant importance to diagnosis and treatment, our analyses show a well-supported resolution within the LD and RF lineages. Moreover, it is important to note that the *terL* phylogenetic tree largely agrees with the 16S rRNA gene based *Borrelia* phylogeny. The only exception being that the 16S tree placed *B. miyamotoi* within the RF clade, while the *terL* tree places it outside both the LD and RF clades^45-48^. This variable phylogenetic position of *B. miyamotoi* indicates that it is distantly related to both LD and RF, but is more closely related to RF than to LD *Borrelia*. This detailed resolution offered by the *terL* gene is useful on two fronts: 1) it reflects the fact that the pathogenesis of *B. miyamotoi* is distinct to RF^46,47^; and 2) as *B. miyamotoi* is the only RF *Borrelia* that can be co-transmitted with LD *Borrelia* species by hard-bodied ticks^47,49,50^, it is really useful to have a molecular marker that can distinguish between LD and RF-causing *Borrelia* spp., and *B. miyamotoi*.

To summarise this section, the *terL* based phylogenetic analysis tightly correlates the *Borrelia* species. The phylogenetic power of the *terL* gene combined with the multi-copy nature suggests that it can be developed as a diagnostic marker for accurate identification of LD, and can be used to differentiate LD from related infections and co-infections such as RF and diseases caused by *B. miyamotoi*.

### Ter-qPCR analytical specificity, sensitivity, and efficiency

To maximise the specificity and sensitivity of our test, we designed a set of primers and a probe to the most conserved regions of terL (to target nine out of 13 terL copies) (Fig. 2 and Supplementary information 2)^51^. The specificity of the primer/probe was confirmed by Blastn and *In silico*’ PCR (http://insilico.ehu.es/PCR/). Positive Ter-qPCR results were also obtained from all the LD *Borrelia* strains listed in Table 2. No positive Ter-qPCR was observed from *B. spielmanii*, or RF-causing *Borrelia* strains as listed in Table 2, or other non-*Borrelia* bacterial strains tested, along with and human DNA samples (detail in Supplementary information 3).

**Table 2.** *Borrelia* strains used in this study

| Lab number | Isolate name | Scientific name | Source | Disease |
| --- | --- | --- | --- | --- |
| B31 | B31 | *Borrelia burgdorferi* | Sven Bergström | LD |
| 1 | 1120 | *Borrelia duttonii* | Sven Bergström | RF |
| 2 | Her HS1 | *Borrelia hemsii* | Sven Bergström | RF |
| 3 | VS185 P9 | *Borrelia burgdorferi* | Sven Bergström | LD |
| 4 | NE218 | *Borrelia valasiana* | Sven Bergström | LD |
| 5 | ACA1 | *Borrelia afzelii* | Sven Bergström | LD |
| 6 | UK filtered | *Borrelia burgdorferi* | Sven Bergström | LD |
| 7 | 190 P9 | *Borrelia garinii* | Sven Bergström | LD |
| 8 | China23 | *Borrelia burgdorferi* | Sven Bergström | LD |
| 9 | CA128 | *Borrelia bisettii* | Sven Bergström | LD |
| 10 | FR64b | *Borrelia miyamotoi* | CDC, USA | RF |
| 11 | HT31 | *Borrelia miyamotoi* | CDC, USA | RF |
| 12 | CIP 109134 | *Borrelia burgdorferi* | Institut Pasteur | LD |
| 13 | CIP 108855T | *Borrelia spielmanii* | Institut Pasteur | LD |
| 14 | CIP 105366T | *Borrelia lusitaniae* | Institut Pasteur | LD |
| 15 | DSM 21467 | *Borrelia valasiana* | DSMZ | LD |
| 16 | DSM 10508 | *Borrelia afzelii* | DSMZ | LD |
| 17 | DSM 10534 | *Borrelia garinii* | DSMZ | LD |
| 18 | DSM 23469 | *Borrelia bavariensis* | DSMZ | LD |
| 19 | Pbi | *Borrelia bavariensis* | Cecilia Hizo-Teufel | LD |
| 20 | NT54 | *Borrelia bavariensis* | Cecilia Hizo-Teufel | LD |

LD: Lyme disease; RF: Relapsing fever

**Fig. 2.**
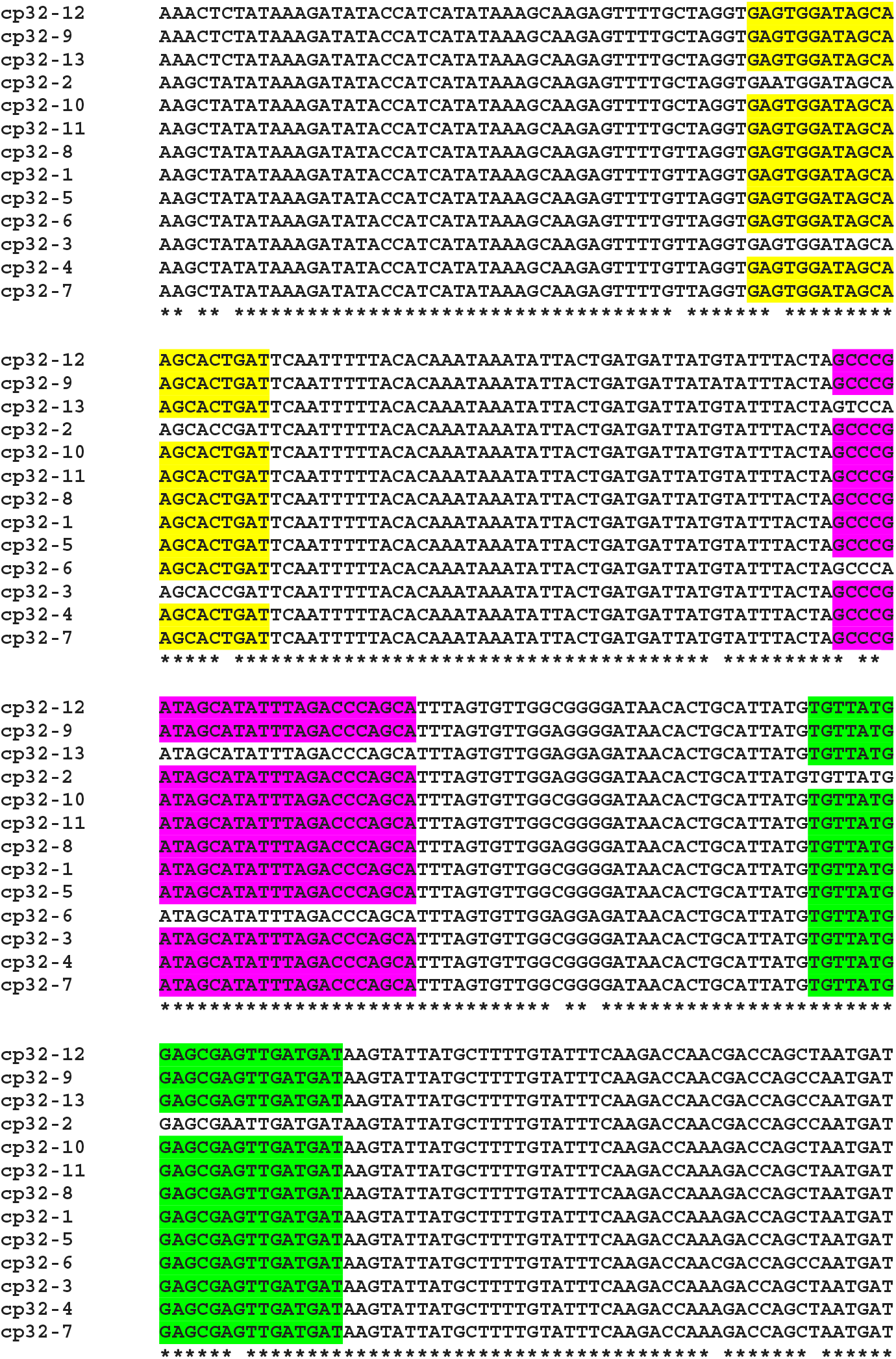
Alignment of the *terL* gene sequence located on the 13 cp32 plasmids (cp32-1 to cp32-13). Identical nucleotides are indicated with an asterisk. Conserved regions of the forward and reverse primers, and probe are highlighted in yellow, green and purple, respectively. Identical primer/probe sequences are present in nine of the *terL* genes, except for those of the cp32-2, cp32-3, cp32-6 and cp32-13 plasmids.

For clinical diagnosis and treatment, it is essential to understand the *Borrelia* load present in the patient, which requires absolute quantification. To develop an absolute quantification assay, we cloned the relevant *terL* fraction into a plasmid (Ter-plasmid) and carried out the Ter-qPCR assay with samples containing background human DNA, this in order to mimic the real clinical samples. We observed a strong linear relationship between the concentration of the Ter-plasmid and Cq (R^2^=0.99) with an amplification efficiency of 99.58% (Fig. 3A). B31 DNA dilutions also displayed a robust linear association with Cq values (R^2^=0.999) with an amplification efficiency of 98.64% (Fig. 3B). This demonstrates the high efficiency of the Ter-qPCR. A close to 100% amplification efficiency from both serial dilution experiments confirmed that our standard curve assay was robust and repeatable^52^.

**Fig. 3.**
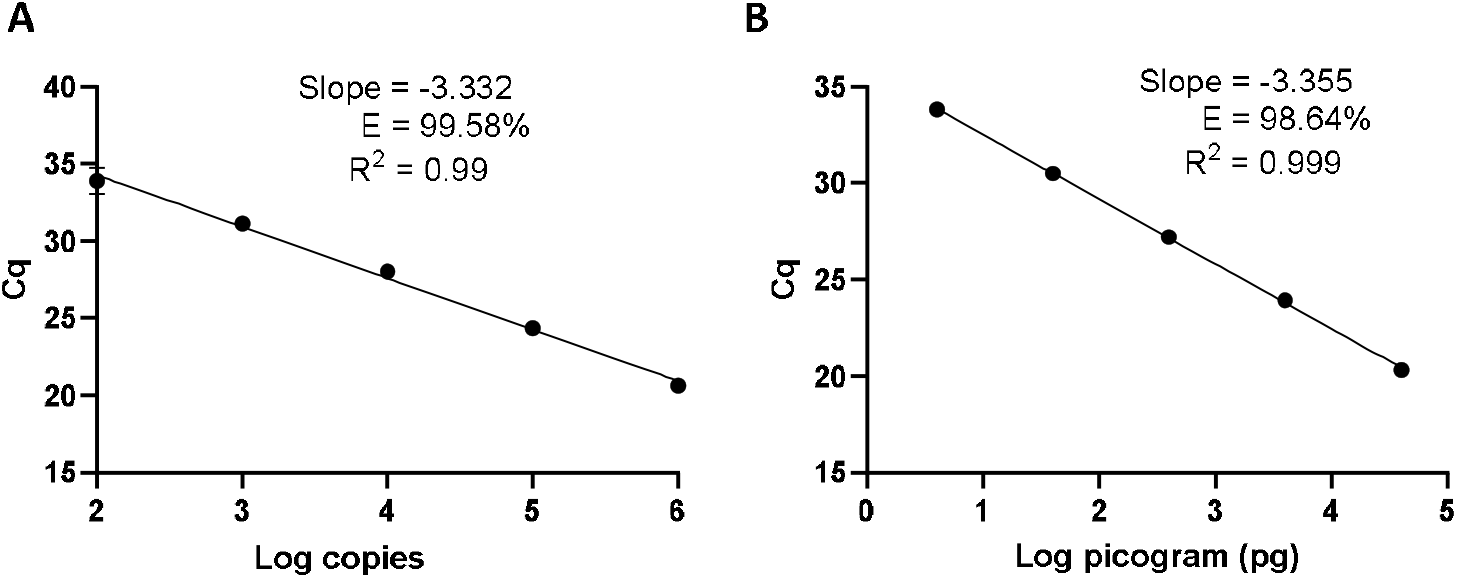
The Ter-qPCR against serial dilutions of the Ter-plasmid and *B. burgdorferi* B31 DNA to measure the LoD and PCR efficiency: (A) Ter-plasmid and (B) B31 DNA. Cq values were plotted against the log values of serial dilutions of the Ter-plasmid and B31 DNA, respectively. Simple regression analysis was carried out using the Graphpad Prism 8.4.3 software. The slope, coefficient of correlation (R^2^) and efficiency of the reaction (E) are shown. Each dot represents the average value from triplicate amplifications, along with the SD.

In this study, we defined the analytical limit of detection (LoD) as the lowest concentration where at least 95% of the technical replicates were positive in the Ter-qPCR^53^. The LoD was calculated from serial dilutions of Ter-plasmids. The proportion of Ter-qPCR positive among replicates was directly correlated with the number of plasmid copies per reaction (Supplementary Fig. 2). For example, one copy of the Ter-plasmid led to two positives out of 10 replicates, while 20 and 40 copies generated 9 and 10 positives out of 10 replicates, respectively. Probit analysis via the SPSS package was used to estimate the LoD and was found to be 22 copies per PCR^54^.

### DNA extraction methods matter: *in vitro* spiked blood samples

To gain an insight into the potential performance of the Ter-qPCR with patients, the Ter-qPCR was applied to *Borrelia*-spiked blood samples. Consistent copy numbers from technical repeats were recovered from samples with ≥1 spike-in *Borrelia* cell (Fig. 4A). This indicates that the Ter-qPCR can potentially detect as low as one *Borrelia* cell from a blood sample. In contrast, the blood sample with 0.1 of a *Borrelia* cell (mimicking the scenario of an extremely low level of *Borrelia* presence in the blood) displayed one copy number out of six repeats (Fig. 4A). It is already established that the number of *Borrelia* cells circulating in the blood is extremely low and is often at the lower end of the detection limit of qPCR^26^. Therefore, it is common to get one PCR amplification out of technical repeats due to stochastic effect when in low concentration^55^. To reflect the low and random distribution nature of *Borrelia* cells in blood, we adopted the following rule for recording copy numbers: the replicate that did not generate a copy number (displayed as ‘failed qPCR’) was scored ‘zero’ ^55,56^. The ‘zero’ *Borrelia* presence in blood was manifested in our later study analysing LD patient samples.

**Fig. 4.**
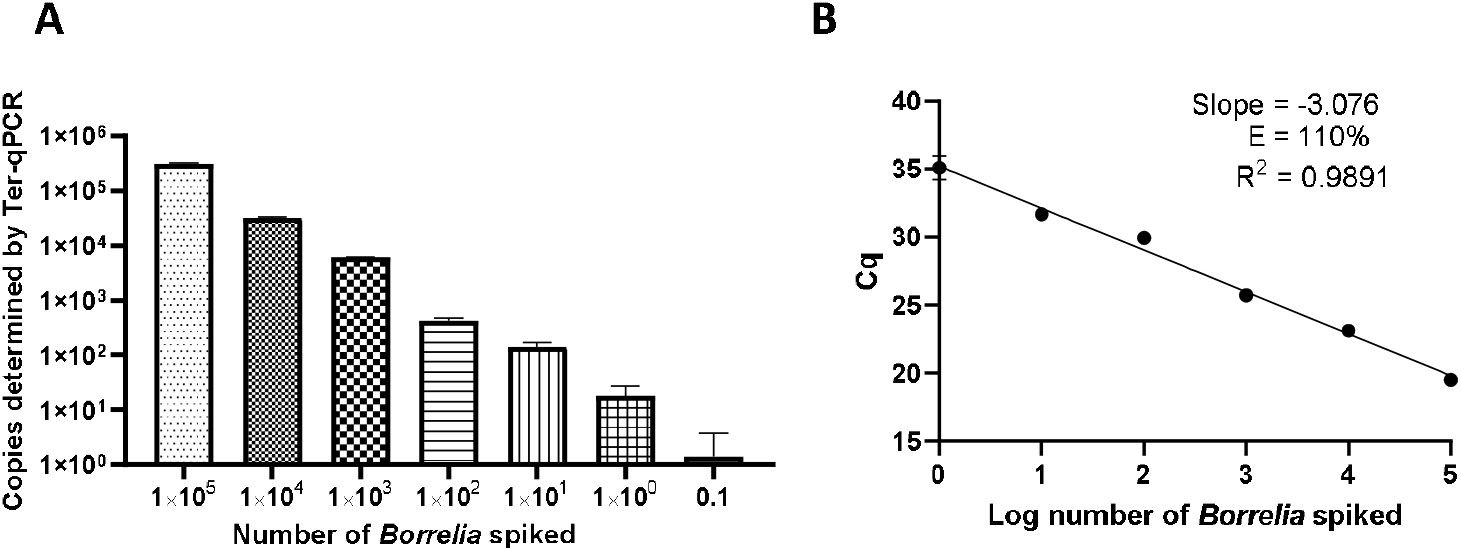
Performance of the Ter-qPCR estimated by examining human blood spiked with tenfold serial dilutions of *Borrelia* cells (10^5^ to 0.1). **A:** bar graph illustrating the number of spike-in *Borrelia* cells and the resulting copy numbers determined by the Ter-qPCR; **B:** Linear regression analysis between the known amount of spiked *Borrelia* cells and the resulting Cq values revealed a strong linear association. The slope, coefficient of correlation (R^2^) and efficiency of the reaction (E) are shown. Each dot represents the average value from triplicate repeats along with SD values obtained from two independent experiments.

To better understand the reliability of the Ter-qPCR, simple linear regression analysis was performed. As seen in Fig. 4B, a linear association (R^2^=0.9891) was observed between the amount of spiked *Borrelia* cells and the resulting Cq values, which demonstrates that the signal intensity of Ter-qPCR correlates with the ‘*Borrelia* load’. In other words, it appears that the Ter-qPCR can detect as low as one *Borrelia* per 300 µl blood, which is equivalent to 3.3 *Borrelia* cells per ml of blood. This is really promising, bearing in mind that evidence from many published studies indicates that *Borrelia* presence in LD patients can range from 1-100 cells/ml^15,25,38,57^. Therefore this single cell sensitivity allows the Ter-qPCR test to revolutionise the way *Borrelia* infection is diagnosed^58^. To put this in context, the current practice to circumvent the bottleneck of low numbers of *Borrelia* circulating in blood is either to culture prior to qPCR or to sample large volumes of blood to artificially increase the amount of PCR templates. Both methods have obvious drawbacks^15,26,59^. Intuitively, by targeting endogenous multi-copy genes, the Ter-qPCR assay offers a much more elegant and reliable way of increasing the amount of PCR template.

We initially chose the phenol method to extract DNA from blood as it is the method of choice when it comes to extracting phage DNA^60,61^. Although the phenol extraction method is in many ways a gold standard, it is cumbersome. To improve the scaling of the assay, we tested commercial DNA extraction kits, such as the DNeasy Blood & Tissue Kit (a column filtration system) and Maxwell RSC Viral Total Nucleic Acid Purification Kit (a magnetic beads-based system) to see whether they could replace the solvent-based method.

Despite a significant effort, including comparisons with bacterial chromosome-targeting 16S qPCR, the phenol extraction method outperformed the other two commercial methods. Thus, the phenol method was used throughout this study. As shown in Fig. 5, regardless of qPCR methods, the phenol approach generated a consistently higher copy number compared to the other two DNA extraction methods. The Ter-qPCR coupled with the phenol method produced significantly higher copy numbers than their counterparts of the 16S qPCR from all spiked samples. Additionally, the Ter-qPCR generated robust copy numbers (three positives out of three replicates) from blood spiked with one *Borrelia* cell (Fig. 5), but the 16S qPCR only displayed amplification once from the triplicate repeats of the same sample. The outstanding sensitivity of Ter-qPCR can also be seen from the markedly lower Cq values (therefore high amounts of PCR target) of the Ter-qPCR than that of the 16S qPCR when both qPCRs were targeting the same B31 genomic DNA (Fig. 6B). The competitive advantage of targeting multicopy genes can also be seen in that the copy numbers determined by the Ter-qPCR were consistently higher than the number of spiked *Borrelia*, while the copy numbers established via the 16S qPCR were numerically about the same as the input *Borrelia* number (Fig. 5).

**Fig. 5.**
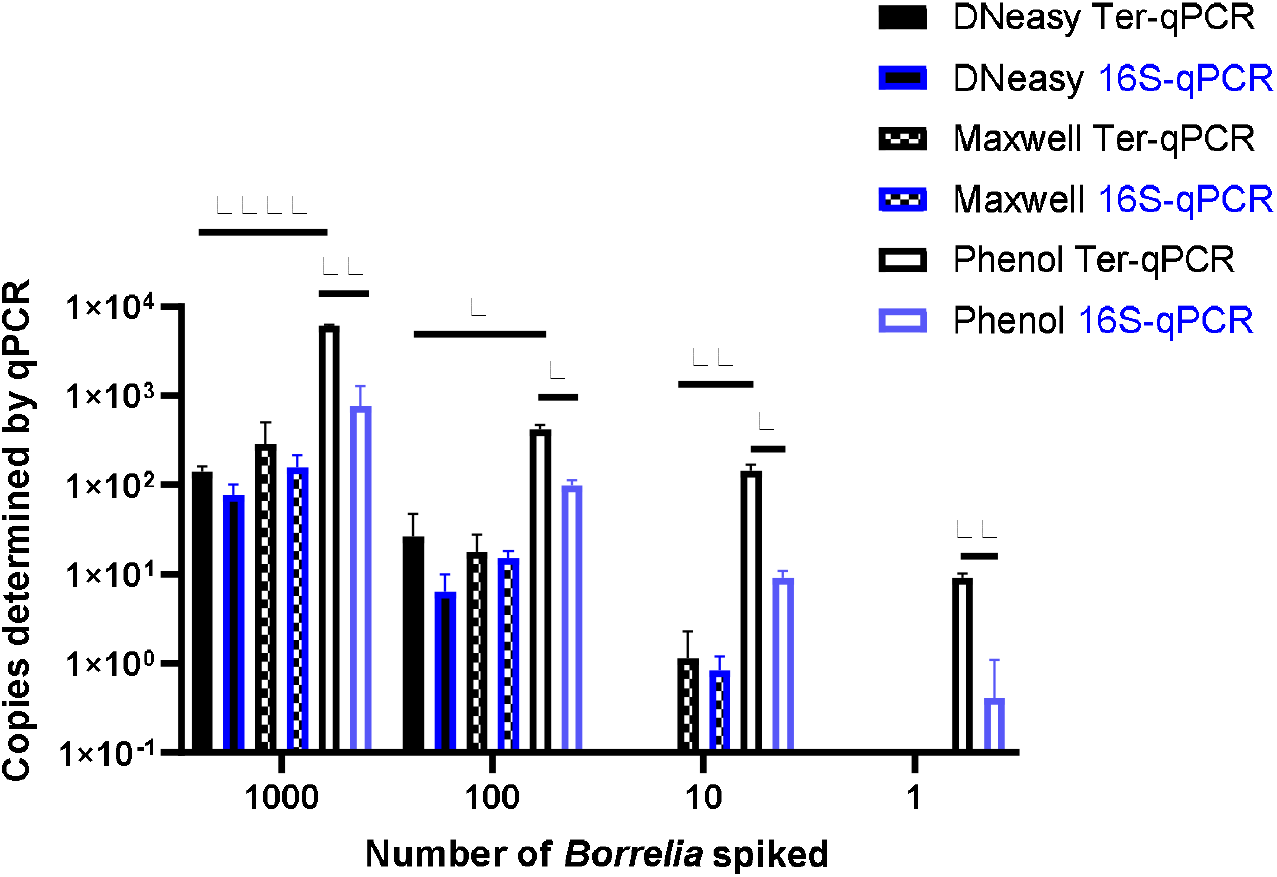
The Ter-qPCR (bars with black border) and 16S qPCR (bars with blue border) against DNA extracted using three methods from human whole blood spiked with tenfold serial dilutions of *Borrelia* cells (10^3^ to 1). The phenol method, DNeasy Blood & Tissue Kit and the Maxwell RSC Viral Total Nucleic Acid Purification Kit were compared for DNA extraction. The Phenol method coupled with the Ter-qPCR displayed consistently and significantly higher quantitation compared to the other two methods. Copy numbers obtained from the Ter-qPCR were significantly higher than those from the 16S qPCR. Values shown are the means from triplicate repeats along with SD values obtained from two independent experiments.

**Fig. 6.**
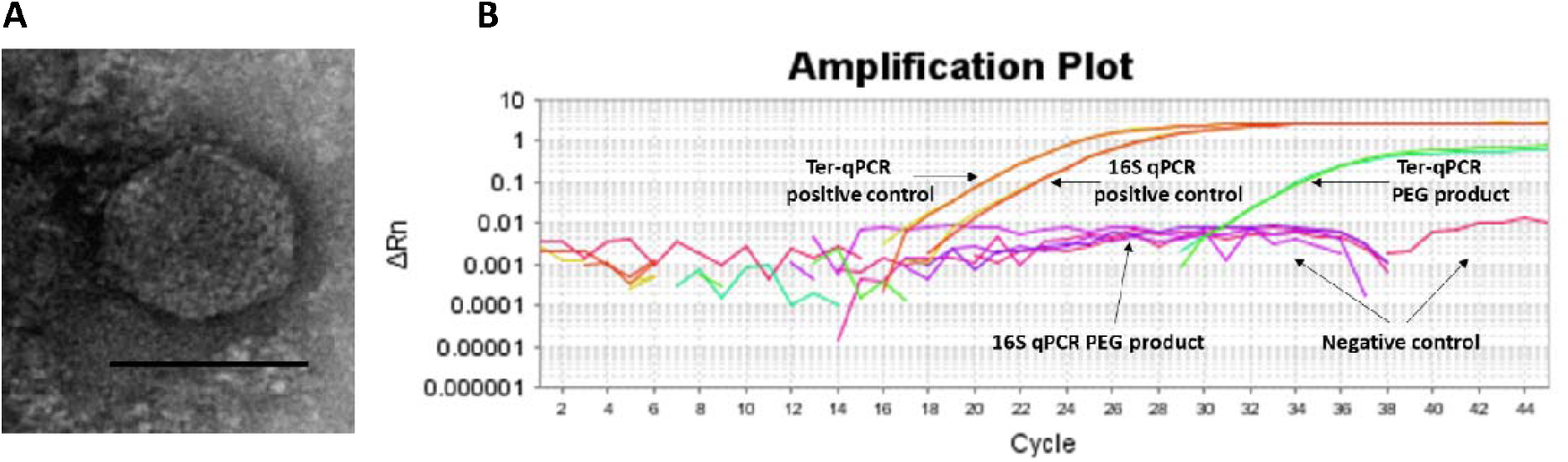
Spontaneous prophage release from *B. burgdorferi* B31 cultures. **A:** Image of the phage-like particles visualised during TEM analysis of the PEG purified cell-free culture filtrates. **B:** DNA extracted from the PEG product tested positive during the Ter-qPCR, but negative to the 16S qPCR, indicating the spontaneous release of cp32 prophages. 1 ng of DNA from B31 was used as a positive control (as annotated in panel B).

### Spontaneous prophage induction provides unique discriminatory power

It is logical that multicopy PCR targets will lead to higher sensitivity when compared to a single-copy PCR target^62^. We determined whether the prophage encoded genes would provide additional sensitivity to detect infections. Since some prophages can escape from their bacterial hosts either by chemical induction or spontaneous prophage induction (SPI), it is entirely possible that multiple *Borrelia* prophages could be released into the blood where they magnify the diagnostic signals and thus can be detected. In the case of *Borrelia, terL*-carrying cp32 prophages have been demonstrated to be susceptible to 1-methyl-3-nitroso-nitroguanidine (MNNG) induction during in vitro culturing^32,35,63^. We hypothesise that *Borrelia* prophages are also capable of SPI which increases the chances of *Borrelia* prophages being induced in the human body.

To prove this SPI hypothesis, we conducted morphological and molecular studies to detect phage presence in cell-free filtrates of *Borrelia* cultures prior and post 36 h incubation. As shown in Fig. 6A, transmission electron microscope (TEM) analysis revealed particles morphologically resembling podoviruses (icosahedral heads and very short tails)^64^ in the polyethylene glycol (PEG)-purified phage fraction derived from *Borrelia* cultures post 36 h incubation. The PEG product was positive to the Ter-qPCR but negative to the *Borrelia* 16S qPCR (Fig. 6B). The Ter-qPCR positive result therefore indicates the presence of cp32 DNA (prophage) in the PEG product. Meanwhile, the PEG product derived from *Borrelia* culture prior incubation showed no phage-like particles and was negative to both the Ter-qPCR and the *Borrelia* 16S qPCR.

Our spontaneous induction data led us to predict that the prophage-based Ter-qPCR would generate a much stronger *Borrelia* signal than bacteria-based qPCR, because LD *Borrelia* species are tissue bound and thus can only circulate in blood transiently and in very low numbers^28^. In contrast, under the scenario of induction, prophages are released into bloodstream, and thus can be a ‘marker’ indicating the presence of *Borrelia*, even though *Borrelia* cells can be hiding and not circulating in the blood. This situation of detecting free phage DNA from human blood bears some resemblance to identifying cell-free circulating DNA (cfDNA) as with cancer diagnosis^65^. Interestingly, the same challenge also stands in terms of the method of choice for isolating cfDNA^66,67^. Phenol chloroform was also highly efficient in recovering cfDNA from clinical samples^67^. Free phages have been discovered from a range of clinical samples, including blood^68,69^. Their biological significance remains to be understood. Considering the fact that metabolic active bacteria can better support phage reproduction^70^, a high level of certain phages in the blood would indicate active infections of their respective bacteria, i.e. a strong *terL* signal would implicate active *Borrelia* infection. Given the fact that most pathogenic bacteria carry many inducible (including SPI) prophages, the rational of diagnosing bacteria by detecting their ‘breakaway’ prophages could revolutionise the current bacteria-focused paradigm of detecting bacterial infections.

### Performance of the Ter-qPCR against clinical samples

As a final important validation step of this work, to determine the potential diagnostic value in a human setting, the Ter-qPCR was applied to 78 individuals belonging to three categories (early LD, late LD patients and healthy volunteers) who were diagnosed by Dr Louis Teulières^6,23,24^. We intended to establish the feasibility of the Ter-qPCR to detect LD. Copy numbers were determined from both blood and serum samples with three technical repeats for each sample type. The technical repeats that did not show detectable copy numbers were scored ‘zero’. The raw data was presented in the Supplementary Table. Visually, there are many more ‘zero’ scores from serum than blood samples, and the mean and median values obtained from blood samples in each category are much higher than those from sera (Supplementary Table). The higher copy numbers determined from the whole blood as compared to the serum samples reflects *Borrelia*’s intracellular life cycle and the fact that Ter-qPCR can detect both prophages that are inside or outside of the *Borrelia* cells (probably due to SPI). Therefore, whole blood is a more robust sample source from which to diagnose LD using PCR assay.

Overall, most of samples showed a low *terL* copy number (<10), which is consistent with the current estimation of a low concentration of *Borrelia* circulating in the blood (Supplementary Table). It is also common to have variation among technical repeats, for example, three technical repeats of the patient No. 25 showed copy numbers of 10, 2.8, and 0, respectively (Supplementary Table). This qPCR variability is due to the stochastic effect of having a low number of PCR templates, and has been observed in PCR detection of low level bacteria in blood before^53,55^. Markedly differences were observed in the mean copy number of early LD (2.4), and late LD (6.9), as well as healthy volunteers (0.8), which suggests a potential positive correlation between the severity of LD and the copy numbers, i.e. higher copy numbers in late LD patients, in contrast to lower copy numbers in early LD patients and healthy volunteers. Also, very encouragingly, the median values from the early (2.0) and late (1.8) LD patients are numerically higher than those of the healthy volunteers (0.7) (Supplementary Table). The fact that some healthy volunteers showed positive with copy numbers, indicates possible asymptomatic *Borrelia* carriage^55^. Most importantly, the apparent marked difference in copy numbers from the three categories is supported by statistical analysis. As shown in Fig. 7, Mann-Whitney *U* test revealed significant differences between early LD, late LD patients and healthy volunteers in terms of *terL* levels determined from whole blood, which offers objective evidence to prove that LD is a fact, and that the early and late LD stages do exist. This statistics-backed difference also suggests the potential application of the Ter-qPCR to distinguish early LD from healthy asymptomatic *Borrelia* carrier.

**Fig. 7.**
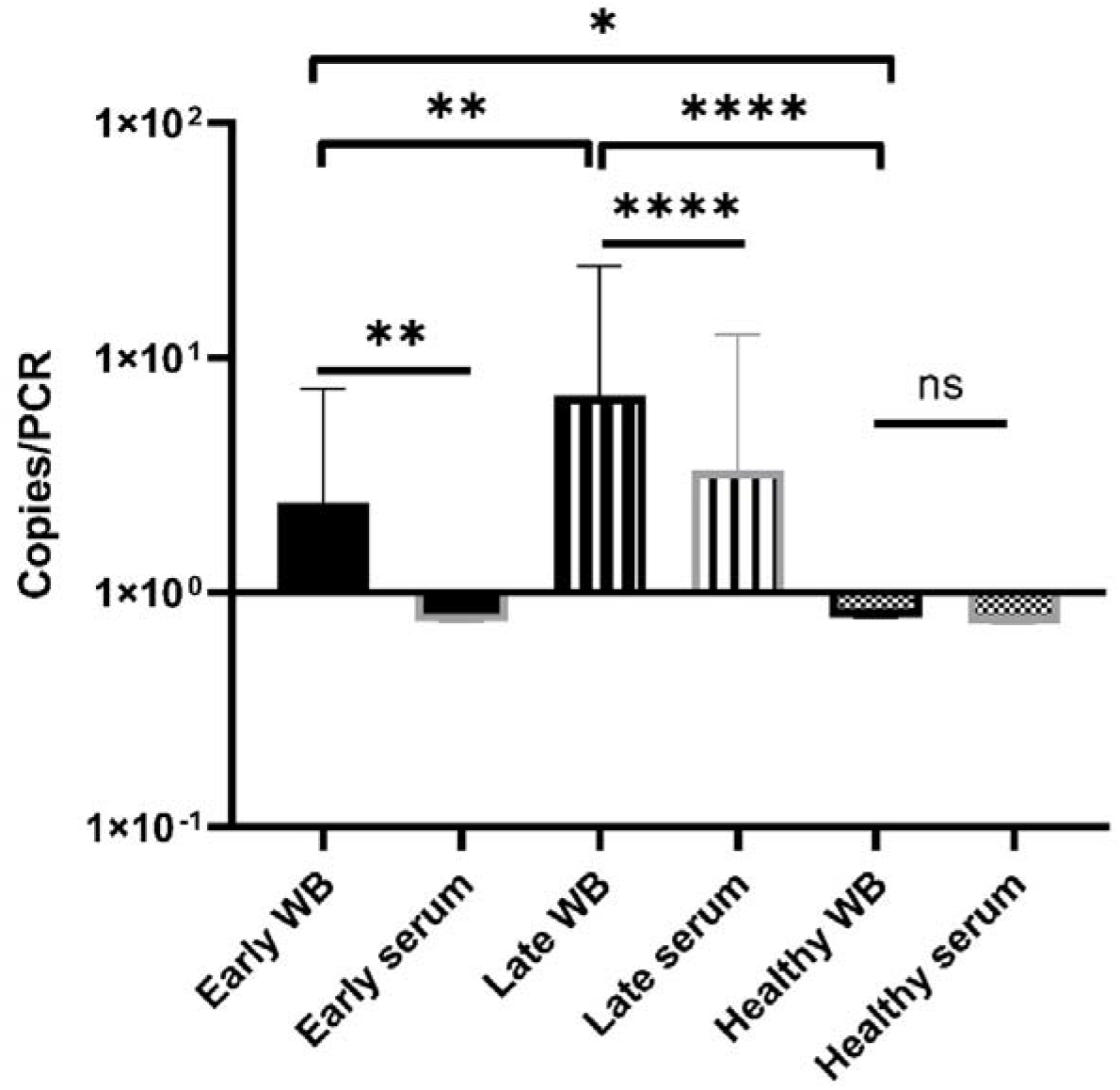
Bar graph displaying the mean *terL* levels (with SD values) of the three respective study groups from whole blood (black border) and serum (grey border) samples. The Mann-Whitney *U* test was used to compare differences between early LD patients, late LD patients, and healthy volunteers. The *terL* copy numbers obtained from the blood samples of late LD patients are significantly higher larger than those of the early LD patients and healthy volunteers, while the *terL* copy numbers of early LD patients are significantly higher larger than those of healthy volunteers. Statistical significance is denoted as ns (p>0.05), * (p<0.05), ** (P<0.01), *** (P<0.001), and **** (P<0.0001).

We conclude that our prophage marker has the ability to differentiate between early and late LD patients. This is the first time such a sensitive and specific test has been developed for LD, and as mentioned, can be greatly beneficial in providing more effective treatments. For example, the Ter-qPCR could be used to monitor LD treatment outcomes, to indicate which treatment option may work best, and to help clinicians measure recovery. We are currently validating a *terL*-based assay targeting RF and *B. miyamotoi*, respectively, working towards a ‘multiplex’ PCR aiming to detect and differentiate LD *Borrelia* spp., RF *Borrelia* spp., and *B. miyamoti* from a single test.

Rapid and accurate detection of microbial pathogens in blood using PCR methods is promising, but hampered by the low-level presence of bacteria in circulating blood, false signals and reduced sensitivity due to unspecific amplification of human DNA. The data presented in this study demonstrates for the first time that targeting phage DNA in blood could offer a rapid diagnosis of bacterial infections and could change the paradigm in the field of PCR detection of bacteria in general.

## Materials and methods

### Phylogenetic analysis

Phylogenetic analysis was constructed using the program Molecular Evolutionary Genetics Analysis (MEGA) 7 according to the previous established method^71,72^. Neighbour-Joining (NJ) with a maximum composite likelihood model and Maximum Likelihood (ML) based on the Tamura-Nei model analyses were conducted on a nucleotide data set of *terL* genes. Support for clades were estimated using a bootstrap analysis implemented in MEGA using 3, 000 replicates. The trees were rooted with phage Lambda (NC_001416) as an outgroup.

### *Borrelia* strains and cultures

The *Borrelia* strains used in this study are listed in Table 1. Ten strains were provided by Professor Sven Bergström, Department of Molecular Biology, Umea University, Sweden. Seven strains were purchased from the Pasteur Institute and DSMZ (German Collection of Microorganisms and Cell Cultures GmbH). Two strains were provided by the Centre for Disease Control and Prevention (CDC), USA, and two by Cecilia Hizo-Teufel from the German National Reference Centre for *Borrelia. Borrelia* cells were grown in 15 ml Falcon™ conical tubes with a culture volume of 14 ml of Barbour-Stoenner-Kelly (BSK) II medium with 7 % rabbit serum (referred to as complete BSKII or c-BSKII) at 35 °C without agitation as previously reported^73^. All culture media was filter sterilised via 0.22 µm pore size filters. Visualisation and counting of *Borrelia* was performed using phase contrast microscopy (Ceti Magnum Trinocular) and a Fuchs Rosenthal Disposable Counting Chamber (C-Chip, NanoEnTek).

### DNA extraction methods

A modified phenol-chloroform method was used to extract DNA from blood and serum samples. In brief, samples were treated with ammonium hydroxide^74^ followed by the classic phenol chloroform DNA extraction method^75^. The resulting DNA pellet was air dried for 5 min, dissolved in 40 µl Tris-Cl (10 mM, pH 8.5), and kept at −20°C. DNeasy Blood & Tissue Kit and Maxwell RSC Viral Total Nucleic Acid Purification Kit were used according to the respective manufacturer instructions. The Thermo Scientific™ NanoDrop™ One Spectrophotometer was used to measure the quantity and quality of DNA samples.

### The Ter-qPCR

Primers and probe were designed using the PrimerQuest® Tool based on an alignment of phage *terL* genes from 13 cp32 plasmids (cp32-1 to cp32-13) carried by *B. burgdorferi* s.l strains. The resulting Ter-qPCR amplified a 147 bp target region^76^. The fluorogenic probe was labelled with 6-carboxyfluorescein (FAM) fluorescent reporter dye at the 5⍰-end, an internal ZEN™ Quencher and an Iowa Black Fluorescent Quencher (IBFQ) to the 3’ (5’FAM/ZEN/3’IBFQ). To rule out PCR inhibition and avoid false negatives, the Ter-qPCR was duplexed with an internal amplification control (IAC) qPCR that generated a 145 bp PCR product^77^. The IAC DNA (accession number FJ357008.1) was synthesised by IDT and added to each Ter-qPCR template. The IAC probe was fluorescently labelled with the fluorescent dye of JOE at the 5⍰-end. The primers and probe targeting the *Borrelia* 16S rRNA gene were adopted from a published paper^78^. All the primers, probes and PrimeTime Gene Expression Master Mix were supplied by IDT.

### Construction of the standard DNA

For absolute quantification, a plasmid carrying the phage *terL* gene fragment (named as Ter-plasmid) was constructed and used as the standard curve. Specifically, a pair of PCR primers was designed using Primer Blast to amplify a 721 bp region of the phage *terL* gene (GenBank accession NC_000948), embracing the 147-bp Ter-qPCR product region. The primers were FTer721:AGACTAAGATGCGGGCAAGA and RTer721:TTGCATCAAGAGCGTCATCA. PCRs were carried out in a LabCycler (SensoQuest GmbH) in a total volume of 50 µl, containing 0.25 mM dNTPs, 3 mM MgCl_2_, 3 µM primers, 50 ng template DNA, 0.5 unit of Taq polymerase (Bioline), and 5 µl 10× Taq buffer (Bioline). Amplification conditions were: 94°C for 2 min, 30 cycles of 94°C for 30 sec, 50°C for 30 sec, 72°C for 1 min, with a final extension of 10 min at 72°C. PCR products were gel-purified using a Qiagen gel extraction kit, and subjected to cloning using the NEB® PCR Cloning Kit according to the manufacturer’s instructions. The recombinant Ter-plasmid DNA carrying the 721-bp terL gene was purified using the Qiagen Plasmid Kit from positive clones. The concentration of the Ter-plasmid was converted into DNA copy number^79^.

### PCR setup

The Ter-qPCR duplexed with the IAC assay was conducted in a 20⍰µl final reaction volume containing 10⍰µl 2X PrimeTime Master Mix, with each primer and each probe at a final concentration of 0.5 and 0.25 μM, respectively, 4⍰µl template DNA, 2 µl IAC (200 genome copies) and nuclease-free water. Standard thermal cycling conditions (Applied Biosystems™ 7500 Fast Real-Time PCR System) were followed with an initial step of 3⍰min at 95°C (polymerase activation), 45 cycles of 15 sec at 95⍰°C (denaturation) and 1 min at 60°C (annealing/extension). The *Borrelia* 16S qPCR setup was carried out according to the previous report^78^

A non-template control (NTC), a positive control of 10 ng *B. burgdorferi* B31 (labelled as B31) DNA and a standard curve made of a series of five tenfold dilutions of the Ter-plasmid DNA (10^6^-10^2^) were included in each run. The qPCR result was analysed and quantified according to the standard curve using FAST7500 software v2.3. For the Ter-qPCR to be valid, the IAC signal should always be produced regardless of the presence or absence of template DNA. All samples were tested in triplicate.

### Analytical specificity and sensitivity

The analytical specificity of the Ter-qPCR assay was determined using both in silico and in vitro analyses. In silico analysis of the primer and probe set was carried out using BLAST and Primer-BLAST^80^, and UCSC In-Silico PCR (https://genome.ucsc.edu/cgi-bin/hgPcr). For in vitro analysis, the Ter-qPCR was applied to DNA extracted from a panel of *Borrelia* strains that cause LD and RF (Table 1), and microbial species that have been used in the lab, including *Clostridium difficile, Clostridium perfringens, Escherichia coli, Pseudomonas aeruginosa, Streptococcus pneumoniae, Staphylococcus aureus, Burkholderia thailandensis, Burkholderia pseudomallei, Haemophilus influenzae* and *Salmonella enterica*. Additionally, human female and male DNA (Promega, G1521 and G1471), and DNA extracted using the phenol method from human whole blood (Cat#: SER-WB10ML from Cambridge Biosciences) were also tested using the Ter-qPCR.

The analytical sensitivity of the Ter-qPCR was firstly evaluated with five tenfold dilutions of the Ter-plasmid (10^6^ to 10^2^) and B31 DNA (40 ng to 4 pg), respectively. Each dilution was tested with three replicates to determine the PCR linearity and amplification efficiency. The limit of detection (LoD) was then estimated by testing Ter-plasmid dilutions from 1000 to 100, 80, 60, 40, 20, 10, 5, and 1 copies/PCR. Ten replicates were used for each dilution. To mimic the real situation of analysing DNA extracted from human samples, all Ter-plasmid serial dilution experiments were conducted with the presence of background human DNA 125 ng per PCR. Probit analysis via the SPSS software was performed to calculate the LoD with 95% probability^81^.

### Spiked blood samples

Actively growing B31 cultures in the early exponential phase (around 10^6^ spirochaetes/ml) was used to spike human whole blood (in duplicate) to generate a final amount of 10^5^, 10^4^, 10^3^, 10^2^, 10, 1, and 0.1 spike-in B31 cells in 300 µl of blood. DNA extraction was carried out using the modified phenol-chloroform method. To compare the different DNA extraction methods, a subset of the B31-spiked blood samples (corresponding to 10^3^, 10^2^, 10, and 1 B31 cells per 300 µl of blood) were also used for DNeasy and Maxwell DNA extractions, respectively. All the resulting DNAs were analysed by both the Ter-qPCR and the *Borrelia* 16S qPCR.

### Spontaneous phage induction and TEM

14 ml of early exponential phase, actively growing B31 cultures (7×10^5^ −10^6^ cells/ml, dominated by free spirochaetal forms) were spun down (6 000 g for 20 min) and washed twice with sterile PBS. The resulting pellets were resuspended in 14 ml c-BSKII and incubated for 36 h at 35 °C. Portions of the B31 suspension prior and post 36 h incubation were centrifuged down, the resulting supernatants were filtered through 0.1 µm pore filters. Extraction of phages from the filtrates were carried out using PEG precipitation^61^. The PEG-purified phage product was used for DNA extraction^82^ and TEM at the Core Biotechnology Services at the University of Leicester^72^. The resulting DNA was examined by the Ter-qPCR and the *Borrelia* 16S qPCR assays.

### Ethic statement and clinical samples

This study was carried out in accordance with protocols reviewed and approved by the Ethics Committee, Comités de protection des personnes (CPP) with investigator reference of Etude Phelix 01617 V1 and CPP reference of 17031. All patients were diagnosed by Dr Louis Teulières according to the ILADS guidelines. A total of 312 samples (156 whole blood and 156 serum samples) were collected from 78 individuals between April-June 2017 (23 healthy volunteers with no ‘identifiable’ LD symptoms, 13 early stage and 42 late stage LD patients). For everyone involved, two tubes of serum and two tubes of Ethylenediaminetetraacetic acid (EDTA)-treated whole blood (approximately one ml in each tube) were provided. Samples were provided in a coded, de-identified manner to preserve patient anonymity. Informed consent was obtained from all participants. The modified phenol chloroform DNA extraction method was applied to all the samples in a duplicate manner. Therefore, there were four DNA samples generated from one individual, two from the whole blood and two from the serum. Triplicate qPCR was applied to each DNA sample, which led to 12 Ter-qPCR data (six from whole blood, six from serum) expressed in copy numbers according to the standard curve for any one individual.

### Statistical analysis

Graphpad Prism 8.4.3. was used for statistical analysis. Descriptive statistics and the D’Agostino-Pearson normality test were used to assess the data distribution. Mann-Whitney U tests were used to determine the significance of the difference between early, late LD patients and healthy volunteers. Furthermore, Probit analysis was carried out via the SPSS software suite (IBM SPSS Statistics 25) to estimate the LoD. The differences were not considered to be significant when the p-values were greater than 0.05. Linear regressions were used to establish correlations between the serial dilution of Ter-plasmids/B31 DNAs and Cq values.

## Data availability

The Ter-qPCR assay includes a set of oligonucleotide primers and Taqman® probes and plasmid DNA as the standard for *in vitro* quantitative detection of *Borrelia* species causing LD. All primers and probes are described in the patent application Ref. P184103.EP.01/T. Other relevant data supporting the findings of the study are available in this article and its Supplementary Information files, or from the corresponding author upon request.

## Supporting information

Summary of copy numbers

Bioinformatic analyses

## Data Availability

The Ter-qPCR assay includes a set of oligonucleotide primers and Taqman probes and plasmid DNA as the standard for in vitro quantitative detection of Borrelia species causing LD. All primers and probes are described in the patent application Ref. P184103.EP.01/T. Other relevant data supporting the findings of the study are available in this article and its Supplementary Information files, or from the corresponding author upon request.

## Acknowledgements

We would like to thank Dr Andrew Millard and Stacy Guiock for their help in proofreading the manuscript. We gratefully acknowledge the main funding received towards the study from the Phelix Research and Development (Phelix R&D, 37 Langton Street, SW10 0JL London, UK, the Charity Number 1156666), the ‘Gift’ funding from Lymefonds, the Netherlands (ANBI-number: 858578438), and the University of Leicester Drug Discovery and Diagnostics (LD3) spring fund 2018.

## Author contributions

JS and YJ contributed to this work equally. JS and MRJC co-conceived the initial idea. JS expanded the initial idea into a coherent scientific project, designed primers and probes and performed in-depth data analysis. YJ was responsible for experimentation and optimisation, data collecting and initial data interpretation. JS and YJ co-wrote the manuscript. MRJC proofread the manuscript and provided valuable comments and suggestions. FP helped with bioinformatic and phylogenetic analyses. LT carried out the ethical application, clinical samples selection and some data analysis.

