## Supplementary material for "Targeting multicopy prophage genes for the differential diagnosis of Lyme disease": Bioinformatic analyses

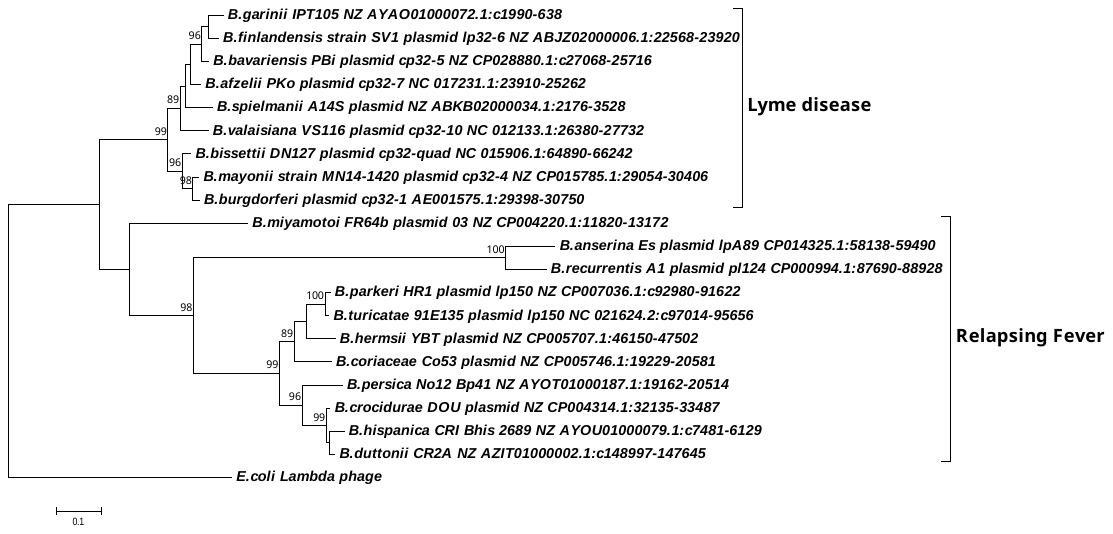

**Supplementary Fig. 1:** A phylogenetic tree constructed based on the *terL* nucleotide sequence showing that the LD, RF *Borrelia* species and *B. miyamotoi* are separated to three clades. Molecular Evolutionary Genetics Analysis (MEGA) package version 7 with maximum likelihood method was used. Scale bar represents the units of the number of base substitutions per site. Support for clades was estimated using a bootstrap analysis in MEGA with 3,000 replicates, values are indicated at the nodes (only values greater than 75 were displayed). The *terL* sequence from phage lambda was used as an outgroup to root the tree.

| **Copy number/PCR** | **Number of replicates** | **Number of PCR positive replicates (% of positive)** |
| --- | --- | --- |
| 1000 | 10 | 10 (100%) |
| 100 | 10 | 10 (100%) |
| 80 | 10 | 10 (100%) |
| 60 | 10 | 10 (100%) |
| 40 | 10 | 10 (100%) |
| 20 | 10 | 9 (90%) |
| 10 | 10 | 7 (70%) |
| 5 | 10 | 4 (40%) |
| 1 | 10 | 2 (20%) |

**Supplementary Fig. 2** **Estimation of LOD of the Ter-qPCR using diluted Ter-plasmid DNA.** The Ter-plasmid DNA was diluted from 1000 copies to 100, 80, 60, 40, 20, 10, 5, and 1 copies/PCR with a human DNA background of 125 ng per PCR. Ten replicates were used for each dilution. The LOD was calculated as 22 copies according to Probit analysis in SPSS with 95% probability. Positive qPCR has typical PCR amplification curve with a Cq<38.

**Supplementary information 1: *Borrelia burgdorferi* B31 phage terminase large subunit (*terL*) nucleotide sequences**

Within the *Borrelia burgdorferi* B31 genome, there are 10 copies of the *terL* gene homologs that are identified from seven circular plasmids (cp32 serials) and three linear plasmids (lp56, lp54 and lp28-2). Eight out of 10 *terL* genes, except those located on lp54 and lp28-2, are highly similar to each other as revealed by alignment below.

```
>CP32-1 - phage terminase large subunit – NC_000948.1 (29398..30750)
GTGAACTTATATCAAACAAAACTTTTTACAACACTACAAAAGGAATACAAAAATAAATATGGAGTTGATATATCACAATTTGTAAAGCTAACAAATTCTTCAATTAATTTTGATAAGTTTGAAGAAGAACAGTTAACTTTAAAACAAAAAAATGTGATAAAAAGCATTAAAAAGAATAATGAAAAGAAGATTATACTCAGCGGAGGCATAGCTAGTGGCAAAACGTATCTTGCATGTTATCTTTTTCTAAAAAGTTTAATTGAAATTAAAAAGTTATACTCTAGTGATACTAATAATTTCATTATAGGGAATTCACAACGTTCAGTTGAAGTTAATGTTTTGGGGCAATTTGAAAAGCTATGTAAACTTCTTAAAATTCCTTATATTCCAAGACATACAAATAATTCATATATTCTGATTGATTCACTACGTATTAATCTATATGGAGGAGATAAGGCAAGTGATTTTGAAAGATTTAGGGGAAGTAATTCGGCACTTATTTTTGTTAATGAGGCTACAACTTTACACAAGCAAACTTTAGAGGAAGTCTTAAAAAGACTAAGATGCGGGCAAGAAACTATTATTTTTGATACTAATCCTGATCATCCAGAACACTATTTTAAAACCGATTATATTGATAATATAGCGACCTTTAAGACATATAAGTTTACAACTTATGATAATGTGCTACTTAGTAAAGGATTTGTCGAAACACAAGAAAAGCTATATAAAGATATACCATCATATAAAGCAAGAGTTTTGTTAGGTGAGTGGATAGCAAGCACTGATTCAATTTTTACACAAATAAATATTACTGATGATTATGTATTTACTAGCCCGATAGCATATTTAGACCCAGCATTTAGTGTTGGCGGGGATAACACTGCATTATGTGTTATGGAGCGAGTTGATGATAAGTATTATGCTTTTGTATTTCAAGACCAAAGACCAGCTAATGATCCTTATATTATGAATATGGTAAAGACTGTTATAGAAAATTTCAATGTGCATACACTGTATTTAGAGGATAGAGATAATACAAAAGGTGCTGGTGGATTGACCCGTGAATACATCTTGCTAAGAAGTAATATAAGCCAATATTTTAGAATTGTTCCAGTTAAGCCAAAGTCTAATAAATTTAGCAGAATAACAACGTTAATTACGCCGTTTACTTACAAAAAACTTTATATTACAAAGTACAGTAGTTCTTCCGTATTTAATGATATTTATTCGTATAAGGGGGATAATAAAACCCATGATGACGCTCTTGATGCAATATCTGCAGCATATTTGATGTTGTCTTTAGGATATAGAGAGCGAAGTGTTCACTTTGGCAATCAAAGATTTTTGTAA

>CP32-3 - phage terminase large subunit – 28871-30223
GTGAACTTATATCAAACAAAACTTTTTACAACACTACAAAAGGAATACAAAAATAAATATGGAGTTGATATATCACAATTTGTAAAGCTAACAAATTCTTCAATTAATTTTGATAAGTTTGAAGAAAAACAGTTAACTTTAAAACAAAAAAATGTGATAAAAAGTATTAAAAAGAATAATGAAAAAAAGATTATATTCAGCGGCGGCATAGCTAGCGGCAAAACGTATCTTGCATGCTATCTTTTTCTCAAAAGTTTAATTGAAAATAAAAAGTTATATTCTAGCGATACGAATAATTTTATTATTGGGAATTCACAACGCTCAGTTGAAGTTAATGTTTTGGGACAATTTGAAAAGCTATGTAAACTTCTTAAAATTCCTTATATTCCAAGACATACAAATAATTCATATATTCTGATTGATTCACTACGTATTAATCTATATGGTGGAGATAAGGCAAGTGATTTTGAAAGATTTAGGGGAAGTAATTCGGCACTTATTTTTGTTAATGAGGCTACAACTTTACACAAGCAAACTTTAGAGGAGGTCCTAAAAAGACTAAGATGCGGGCAAGAAACTATTATTTTTGATACTAATCCTGATCATCCAGAACACTATTTTAAAACCGATTATATTGATAATATAGCGACATTTAAGACATATAATTTTACAACTTATGATAATGTTCTACTTAGTAAAGGATTTATCGAAACACAAGAAAAACTCTATAAAGATATACCATCATATAAAGCAAGAGTTTTGCTAGGTGAGTGGATAGCAAGCACTGATTCAATTTTTACACAAATAAATATTACTGATGATTATGTATTTACTAGTCCAATAGCATATTTAGACCCAGCATTTAGTGTTGGAGGAGATAACACTGCATTATGTGTTATGGAGCGAGTTGATGATAAGTATTATGCTTTTGTATTTCAAGACCAACGACCAGCCAATGATCCTTATATTATGAATATGGTAAAGACCGTTATAGAAAATTTCAATGTGCATACACTGTATTTAGAGGATAGAGATAATACAAAAGGTGCTGGTGGATTGACCCGCGAATACATCTTACTAAGAAATAATATAAGCCAATATTTTAGAATTGTTCCAGTTAAGCCAAAGTCTAATAAATTTAGCAGAATAACAACGTTAATTACGCCGTTTACTTATAAGAAACTTTACATTACAAAGTACAGTAGTTCTTCTGTATTTAATGATATTTATTCGTATAAGGGGGATAGCAAAACCCATGATGATGCTCTTGATGCAATGTCTGCAGCATATTTGATGTTGTCTTTAGGATATAGAGAGCGAAGTGTTCACTTTGGCAATCAAAGATTTTTGTAA

>CP32-4 - phage terminase large subunit – 28947-30299
GTGAACTTATATCAAACAAAACTTTTTACAACACTACAAAAGGAATACAAAAATAAATATGGAGTTGACATATCACAATTTGTAAAGCTAACAAATTCTTCAATTAATTTTGATAAGTTTGAAAAAGAACAGTTAACCATAAAACAAAAAAATGTTATAAAAAGTATTCAAAAAAATAATGAAAAGAAGATTATACTCAGCGGCGGCATAGCTAGCGGCAAAACGTATCTTGCATGTTATCTTTTTCTCAAAAGTTTAATTGAAAATAAAAAGTTATACTCTGGTGATATTAATAATTTTATTATTGGGAATTCACAACGCTCAGTTGAAGTTAATGTTTTGGGACAATTTGAAAAGCTATGTAAACTTCTTAAAATTCCTTATATCCCAAGACATACAAATAATTCATATATATTAATTGATTCACTTCGTATTAATCTATATGGAGGAGATAAGGCAAGTGATTTTGAAAGATTTAGAGGCAGTAATTCGGCACTTATTTTTGTTAATGAGGCTACTACTTTACACAAGCAAACTTTAGAGGAGGTCTTAAAAAGACTTAGGTGCGGACAAGAAACTATTATTTTTGATACTAATCCTGATCATCCAGAACACTATTTTAAAACCGATTATATTGATAATATAGCGACATTTAAGACATATAATTTTACAACTTATGATAATGTGCTACTTAGTAAAGGATTTATCGAAACACAAGAAAAACTCTATAAAGATATACCATCATATAAAGCAAGAGTTTTGCTAGGTGAGTGGATAGCAAGCACCGATTCAATTTTTACACAAATAAATATTACTAATGATTATGTATTTACTAGCCCGATAGCATATTTAGACCCAGCATTTAGTGTTGGAGGGGATAACACTGCATTATGTGTTATGGAGCGAGTTGATGATAAGTATTATGCTTTTGTATTTCAAGACCAACGACCAGCCAATGATCCTTATATTATGAATATGGTAAAGACCGTTATAGAAAATTTCAATGTGCATACACTGTATTTAGAGGATAGAGATAATACAAAAGGTGCTGGTGGATTGACCCGCGAATACATCTTGCTAAGAAATAATATAAGCCAATATTTTAGAATTGTTCCAGTTAAGCCAAAGTCTAATAAATTTAGCAGAATAACAACGGTAATTACGCCGTTTACTTATAAGAAACTTTACATTACAAAGTACAGTAGTTCTTCTGTATTTAATGATATTTATTCGTATAAGGGGGATAGCAAAACCCATGATGATGCTCTTGATGCAATGTCTGCAGCATATTTGATGTTGTCTTTAGGATATAGAGAGCGAAGTGTTCACTTTGGCAATCAAAGATTTTTGTAA

>CP32-6 - phage terminase large subunit – 28486-29838
GTGAACTTATATCAAACAAAACTTTTTACAACACTACAAAAGGAATACAAAAATAAATATGGAGTTGATATATCACAATTTGTAAAGCTAACAAATTCTTCAATTAATTTTGATAAGTTTGAAGAAAAACAGTTAACTTTAAAACAAAAAAATGTGATAAAAAGTATTAAAAAGAATAATGAAAAAAAGATTATACTCAGCGGCGGCATAGCTAGCGGCAAAACGTATCTTGCATGCTATCTTTTTCTCAAAAGTTTAATTGAAAATAAAAAGTTATATTCTAGCGATACGAATAATTTTATTATTGGGAATTCACAACGCTCAGTTGAAGTTAATGTTTTGGGACAATTTGAAAAGCTATGTAAACTTCTTAAAATTCCTTATATTCCAAGACATACAAATAATTCATATATTCTGATTGATTCACTACGTATTAATCTATATGGTGGAGATAAGGCAAGTGATTTTGAAAGATTTAGGGGAAGTAATTCGGCACTTATTTTTGTTAATGAGGCTACAACTTTACACAAGCAAACTTTAGAGGAGGTCTTAAAAAGACTAAGATGCGGGCAAGAAACTATTATTTTTGATACTAATCCCGATCATCCAGAACACTATTTTAAAACCGATTATATTGATAATATAGCGACCTTTAAGACATATAATTTTACAACTTATGATAATGTGCTACTTAGTAAAGGATTTGTCGAAACACAAGAAAAGCTATATAAAGATATACCATCATATAAAGCAAGAGTTTTGCTAGGTGAGTGGATAGCAAGCACTGATTCAATTTTTACACAAATAAATATTACTGATGATTATGTATTTACTAGCCCGATAGCATATTTAGACCCAGCATTTAGTGTTGGCGGGGATAACACTGCATTATGTGTTATGGAGCGAGTTGATGATAAGTATTATGCTTTTGTATTTCAAGACCAAAGACCAGCTAATGATCCTTATATTATGAATATGGTAAAGACTGTTATAGAAAATTTCAATGTGCATACACTGTATTTAGAGGATAGAGATAATACAAAAGGTGCTGGTGGATTGACCCGTGAATACATCTTGCTAAGAAGTAATATAAGCCAATATTTTAGAATTGTTCCAGTTAAGCCAAAGTCTAATAAATTTAGCAGAATAACAACGTTAATTACGCCGTTTACTTACAAAAAACTTTACATTACAAAGTACAGTAGTTCTTCTGTATTTAATGATATTTATTCGTATAAGGGGGATAGCAAAACCCATGATGATGCTCTTGATGCAATGTCTGCAGCATATTTGATGTTGTCTTTAGGATATAGAGAGCGAAGTGTTCACTTTGGCAATCAAAGATTTTTGTAA

>CP32-7 - phage terminase large subunit – 29448-30800
GTGAACTTATATCAAACAAAACTTTTTACAACACTACAAAAGGAATACAAAAATAAATATGGAGTTGATATATCGCAATTTATAAAGCTAACAAATTCTTCAATTAATTTTGATAAGTTTGAAGAAGAACAGCTAACTTTAAAACAAAAAAATGTGATAAAAAGCATTAAAAAGAATAATGAAAAGAAGATTATACTCAGTGGAGGTATAGCTAGCGGCAAAACGTATCTTGCATGTTATCTTTTTCTCAAAAGTTTAATTGAAAATAAAAAGCTATATTCTAGCGATACGAATAATTTCATTATAGGGAATTCACAACGTTCAGTTGAAGTTAATGTTTTGGGACAATTTGAAAAGCTATGTAAACTTCTTAAAATTCCTTATATTCCAAGACATACAAATAATTCATATATTTTGATTGATTCACTACGTATTAATCTATATGGTGGAGATAAGGCAAGTGATTTTGAAAGATTTAGGGGAAGTAATTCAGCACTTATTTTTGTGAATGAGGCTACAACTTTACACAAGCAAACTTTAGAGGAAGTCTTAAAAAGACTAAGGTGTGGGCAAGAAACTATTATTTTTGATACTAACCCCGATCATCCGGAACACTATTTTAAAACCGATTATATTGATAATATAGCAACTTTTAAGACATATAATTTTACAACTTATGATAATGTTCTACTTAGCAAAGGATTTATCGAAACTCAAGAAAAGCTATATAAAGATATACCATCATATAAAGCAAGAGTTTTGCTAGGTGAATGGATAGCAAGCACCGATTCAATTTTTACACAAATAAATATTACTAATGATTATGTATTTACTAGCCCGATAGCATATTTAGACCCAGCATTTAGTGTTGGAGGGGATAACACTGCATTATGTGTTATGGAGCGAGTTGATGATAAGTATTATGCTTTTGTATTTCAAGACCAAAGACCAGCCAATGACCCGTATATTATGAATATGGTTAAGACCGTTTTAGAGAATTTTAATGTACATACACTTTATTTAGAAGATAGAGACAATACAAAAGGTGCTGGTGGATTGACTCGTGAATACATGTTGCTAAGAAATAATATGGGTCAATATTTTAGAATTGTTCCAGTTAAGCCAAAGTCTAATAAATTTAGCAGAATAACAACGTTAATTACGCCGTTTACTTATAAGAAACTTTACATTACAAAGTACAGCAGTTCTTCTGTATTTAATGATATTTATTCGTATAAAGGAGATAACAAAACCCATGATGATGCTCTTGATGCAATATCTGCAGCATATTTGATGTTGTCTTTAGGGTATAGAGAGAGAAGTGTTCACTTTGGCAATCAAAGATTTTTGTAA

>CP32-8 - phage terminase large subunit – 29533-30885
GTGAACTTATATCAAACAAAACTTTTTACAACACTACAAAAGGAATACAAAAATAAATATGGAGTTGATATATCACAATTTGTAAAGCTAACAAATTCTTCAATTAATTTTGATAAGTTTGAAGAAGAACAGTTAACTTTAAAACAAAAAAATGTGATAAAAAGCATTAAAAAGAATAATGAAAAGAAGATTATACTCAGCGGAGGCATAGCTAGTGGCAAAACGTATCTTGCATGTTATCTTTTTCTAAAAAGTTTAATTGAAAATAAAAAGTTATACTCTAGTGATACTAATAATTTCATTATAGGGAATTCACAACGTTCAGTTGAAGTTAATGTTTTGGGGCAATTTGAAAAGCTATGTAAACTTCTTAAAATTCCTTATATTCCAAGACATACAAATAATTCATATATTCTGATTGATTCACTACGTATTAATCTATATGGAGGAGATAAGGCAAGTGATTTTGAAAGATTTAGGGGAAGTAATTCGGCACTTATTTTTGTTAATGAGGCTACAACTTTACACAAGCAAACTTTAGAGGAAGTCTTAAAAAGACTAAGATGCGGGCAAGAAACTATTATTTTTGATACTAATCCTGATCATCCAGAACACTATTTTAAAACCGATTATATTGATAATATAGCGACCTTTAAGACATATAATTTACAACTTATGATAATGTGCTACTTAGTAAAGGATTTGTCGAAACACAAGAAAAGCTATATAAAGATATACCATCATATAAAGCAAGAGTTTTGTTAGGTGAGTGGATAGCAAGCACTGATTCAATTTTTACACAAATAAATATTACTGATGATTATGTATTTACTAGCCCGATAGCATATTTAGACCCAGCATTTAGTGTTGGAGGGGATAACACTGCATTATGTGTTATGGAGCGAGTTGATGATAAGTATTATGCTTTTGTATTTCAAGACCAAAGACCAGCTAATGATCCTTATATTATGAATATGGTAAAGACTGTTATAGAAAATTTCAATGTGCATACACTGTATTTAGAGGATAGAGATAATACAAAAGGTGCTGGTGGATTGACCCGTGAATACATCTTGCTAAGAAGTAATATAAGCCAATATTTTAGAATTGTTCCAGTTAAGCCAAAGTCTAATAAATTTAGCAGAATAACAACGTTAATTACGCCGTTTACTTACAAAAAACTTTATATTACAAAGTACAGTAGTTCTTCCGTATTTAATGATATTTATTCGTATAAGGGGGATAATAAAACCCATGATGACGCTCTTGATGCAATATCTGCAGCATATTTGATGTTGTCTTTAGGATATAGAGAGCGAAGTGTTCACT
TTGGCAATCAAAGATTTTTGTAA

>CP32-9 - phage terminase large subunit – 29299-30651
GTGAACTTATATCAAACAAAACTTTTTACAACACTACAAAAGGAATACAAAAATAAATATGGAGTTGACATATCACAATTTGTAAAGCTAGCAAATTCTTCAATTAATTTTGATAAGTTTGAAGAAAAACAGTTAACTTTAAAACAAAAAAATGTGATAAAAAGTATTAAAAAGAATAATGAAAAAAAGATTATACTCAGCGGAGGCATAGCTAGTGGCAAAACGTATCTTGCATGTTATCTTTTTCTAAAAAGTTTAATTGAAAATAAAAAGTTATACTCTAGTGATACTAATAATTTCATTATTGGGAATTCACAACGCTCAGTTGAAGTTAATGTTTTAGGACAATTTGAAAAGCTATGTAAACTTCTTAAAATTCCTTATATCCCAAGACATACAAATAATTCATATATTCTGATTGATTCACTACGTATTAATCTATATGGTGGAGATAAGGCAAGTGATTTTGAAAGATTTAGGGGAAGTAATTCAGCACTTATTTTTGTTAATGAGGCTACAACTTTACACAAGCAAACTTTAGAGGAGGTCTTAAAAAGACTAAGATGCGGGCAAGAAACTATTATTTTTGATACTAATCCTGATCATCCAGAACACTATTTTAAAACCGATTATATTGATAATATAGCGACATTTAAGATATATAATTTTACAACTTATGATAATGTGCTACTTAGTAAAGGATTTATCGAAACACAAGAAAAACTCTATAAAGATATACCATCATATAAAGCAAGAGTTTTGCTAGGTGAGTGGATAGCAAGCACTGATTCAATTTTTACACAAATAAATATTACTGATGATTATATATTTACTAGCCCGATAGCATATTTAGACCCAGCATTTAGTGTTGGAGGGGATAACACTGCATTATGTGTTATGGAGCGAGTTGATGATAAGTATTATGCTTTTGTATTTCAAGACCAACGACCAGCCAATGATCCTTATATTATGAATATGGTAAAGACCGTTATAGAAAATTTCAATGTGCATACACTGTATTTAGAGGATAGAGATAATACAAAAGGTGCTGGTGGATTGACCCGCGAATACATCTTGCTAAGAAATAATATAAGCCAATATTTTAGAATTGTTCCAGTTAAGCCAAAGTCTAATAAATTTAGCAGAATAACAATGTTAATTACGCCGTTTACTTACAAAAAACTTTATATTACAAAGTACAGTAGTTCTTCTGTATTTAATGATATTTATTCGTATAAGGGGGATAGCAAAACCCATGATGATGCTCTTGATGCAATATCTGCAGCATATTTGATGTTGTCTTTAGGATATAGAGAGCGAAGTGTTCACTTTGGCAATCAAAGATTTTTGTAA

>LP56 - phage terminase large subunit - 32455-33807
GTGAACTTATATCAAACAAAACTTTTTACAATACTACAAAAGGAATACAAAAATAAATATGGAGTTGATATATCACAATTTGTAAAGCTAACAAATTCTTCAATTAATTTTGATAAGTTTGAAGAAGAGCAGCTAACTTTAAAACAAAAAAATGTGATAAAAAGCATTAAAAAGAATAATGAAAAAAAGATTATACTCAGCGGCGGCATAGCTAGCGGCAAAACGTATCTTGCATGTTATCTTTTTATAAAAAGTTTAATTGAAAATAAAAAGCTATATTCTAGCGATACGAATAATTTCATTATAGGGAATTCACAACGCTCAGTTGAAGTTAATGTTTTGGGGCAATTTGAAAAGCTATGCAAACTGCTTAAAATTCCTTATATTCCAAGACATACAAATAATTCATATATTCTGATTGATTCACTGCGAATTAATCTACATGGTGGAGATAAGGCAAGTGATTTTGAAAGATTTAGGGGAAGTAATTCAGCACTTATTTTTGTTAATGAGGCTACAACTTTACACAAGCAAACTTTAGAGGAAGTCTTAAAAAGATTAAGATGCGGGCAAGAAACTATTATTTTTGATACTAATCCTGATCATCCAGAACACTATTTTAAAACCGATTATATTGATAATATAGCGACCTTTAAGACATATAATTTTACAACTTATGATAATGTTCTACTTAGTAAAGGATTTATCGAAACACAAGAAAAGCTCTATAAAGATATACCATCATATAAAGCAAGAGTTTTGCTAGGGGAGTGGATAGCAAGCACCGATTCAATTTTTACACAAATAAATATTACTAATGATTATGTATTTACTAGCCCGATAGCATATTTAGACCCAGCATTTAGTGTTGGCGGGGATAACACTGCATTATGTGTTATGGAGCGGGTTGATGATAAGTATTATGCTTTTGTATTTCAAGACCAACGACCAGCCAATGACCCGTATATTATGAATATGGTTAAGACCGTTTTAGAAAATTTTAATGTACATACATTTTATTTAGAAGATAGAGACAATACAAAAGGTGCTGGTGGATTGACCCGCGAATACATCTTGCTAAGAAATAATATGGGTCAATATTTTAGAATTGTTCCAGTTAAGCCAAAGTCTAATAAATTTAGCAGAATAACAGCGTTAATTACGCCGTTTATTTATAAGAAACTGTACATTACGAAGTACAGCAGTTCTTCTGTATTTAATGATATTTATTCGTATAAAGGAGATAACAAAACCCATGATGATGCTCTTGATGCAATATCTGCAGCATATTTGATGTTGTCTTTAGGGTATAGAGAGAGAAGTGTTCACTTTGGCAATCAAAGATTTTTGTAA

>LP54 - pbsx family phage terminase – NC_001857.2 (18725..20077) (this one is only distantly similar to the above eight terminase sequences, therefore not possible to include this one in the primers and probe design)
TTGAAATTCTTAAGATCATTAGCGTTTCTCAATCTTCAGAAAAAGTTTAAAAATAAGTTTAATATTAACATTTTGGATTATATTAAGCCAAAACCAACCAAAATTTGTTTTAAAGATTTTGAAAATAAATACTTAACTGCTAAGCAAAAGGAAGTTCTTTTTGACATAGAAAGTAACAATTATTCAAAAGTAATATTTAGCGGTGGGATTGCAAGTGGTAAGACGTTTTTAGCTTCATATTTGCTTGTTAAAAAGCTTATTGAAAACAAATCTTTTTATGAGCAAGATACCAATAATTTTATTATAGGCAACTCAATTGGTTTGTTGATGACAAATACTGTAAAGCAAATAGAGAAAATTTGCAGCTTGCTGGGAATTGATTACGAGAAAAAGAAAAGCGGGCAGTCTTTTTGTAAGATAGCAGGTCTTAAGCTTAATATTTACGGGGGTAAAAACAGAGATGCTTTTTCCAAGATTCGTGGGGGCAATAGTGCAATAATTTATGTAAATGAAGCAACAGTAATTCACAGAGAAACTTTACTTGAAGTAATAAAAAGGCTTAGAAAGGGTAAAGAAATTATTATTTTTGATACAAATCCGGAAAGCCCTGCGCATTATTTTAAAACCGATTATATTGAAAATACAGATGTTTTTAAGACATATAATTTTACAACTTATGACAATCCTTTAAATTCAGCAGATTTTATTCAAACTCAGGAAAAACTTTACAGGCGTTTTCCCGCCTATAGGGCTCGTGTGCTTTACGGGGAGTGGATTTTAAATGAATCTACGCTATTTAATGAGATGATTTTCAATCAAGATTATGAATTTAAAAGCCCAATAATGTACATTGATCCTGCATTCTCAGTGGGTGGAGACAATACAGCTATTTGTGTTTTAGAGCGCACTTTTGAGAAGTTTTATGCATATATTTATCAAGACCAAAAACCAGTAAGTGATAGTTTAATGCTTGCTTCCATTCAAGTTTTAATAGAAAATTTCAATGTAAATACCGTTTACATTGAGGAGAGAGACAGTACCAAGGGGGATGGAATTTTAACTAAAACAATTCTTTTCTTGAGAAACAAAAGCAGTCACTATTTTAAAGTTGCACCAATAAAACCTTTAAGCAATAAATTTAAAAGAATATGCGCGCTTATTCCTTTGTTTGAAAGCCGCAAAATAGAATTTTTAAAAATAATAAGTAAAAATGTAATATCTGATATTTACAGCTACAAAGGAGATGGCAAAACAAAAGACGATGCGCTTGATTCTCTTGCAAATGCGTATCTTCTTTTAACGCTAAATTATAAAGAAAAATTATTTCATTTCGGCAGGTTTAAATATTTATAA

>LP28-2 – Hypothetical protein  – NC_001852.1 (16494..17684, complement) (this one is not annotated as termiase gene)
ATGAGACTAAGGCGACTTCCAGTATATGTTGATGCCTACAAAGAAAAGCCCAATGCTGAAATTTTCATATACTACTCAAGTAGGGGAACTGGTAAAACCTACGACATTGCAACTGTTAATTTAGAAAGAAAATTTAGTGCTGATGGCGGAGATACCCTTGCAATTAGAAAAAAGAAAAACAAAACAACACAATCAATACACAAAGAAATTTTAGAACTTTTAAGCATATACAACTTAAGAAAATTTTTCAATATAAGCAAAGCAAAAATTGAAAGTAAGAGTTTGATTTTTGGGAAAAAACGTGCTTTTGTTTTTGAAGGGGGGCATGATACAAGAGATTTAAAATCTTATGCGCATTTTAAGGACTTATGGCTGGAAGAAGCCAATCAGTTTAGCGCTGATGATATAGAAATGCTTGTCCCTACAATGAGAGAACAAGGCGGCAGAATCTATATGTCAAGCAATCCGGTGCCTAAATCACATTGGCTATACAAAAGGTACTTATCAAATCAAGACAATCCTGCTGTGTGTATAATCAAAAGCACTTACCGTGACAACCCATTTTTAAATGGTGGAGACGTACAAGCTTGGCTTGAAAAACAAAGACTTGCATATCATGGCAATGACATTGGTTTTAGAATTGAGGTTTTAGGAGAAGAGTTTGATTTTGGTACAGCAAGGCTAATTAAAAAATTTAATGTGTGTGGTCCCGAAATTCTTTCCAGAGCTAATGGAAGCTATTATACAGGAATACACGTTAAAGGCAACAGAATTTGTTTTTTAGAAATTCTTGTTGGAAGAATTTCCTATCTTCCAGTTGTAATTATTACAAACGCATGTAGTAAAGTTTTACTATCAAAAACTGATTACCAATCCGAAATTAATAAATTTAAAGGGGTTTTTGTATTGCCAACAGCAAGAGAAGAACTCAAATATGTATTTTCTCGTTTTGGTAGAGGTACTTTGCTTGCAAAAAAACGAAACTTGTATTCACTCTCAGACTATTTGATTCCGTCTAATCTTAATGTGGTAAACAAACCCGAAACCACTGATGTAATTTCAGAGTTCAACGAAACTGAATATTATTATGATGAATCTAGTGCAGAAGATAGCGAAGTTACAAATTTTGTTATGCAAAAAGATTTGGTATACATCCCGGCATTTCTCAATGCCATATCGGTTTTTAGCTAA

NT Alignment of phage terminase large subunit from all CP32 Plasmids & LP56.

CP32-1          GTGAACTTATATCAAACAAAACTTTTTACAACACTACAAAAGGAATACAAAAATAAATAT 60
CP32-8          GTGAACTTATATCAAACAAAACTTTTTACAACACTACAAAAGGAATACAAAAATAAATAT 60
CP32-6          GTGAACTTATATCAAACAAAACTTTTTACAACACTACAAAAGGAATACAAAAATAAATAT 60
CP32-4          GTGAACTTATATCAAACAAAACTTTTTACAACACTACAAAAGGAATACAAAAATAAATAT 60
CP32-9          GTGAACTTATATCAAACAAAACTTTTTACAACACTACAAAAGGAATACAAAAATAAATAT 60
CP32-3          GTGAACTTATATCAAACAAAACTTTTTACAACACTACAAAAGGAATACAAAAATAAATAT 60
CP32-7          GTGAACTTATATCAAACAAAACTTTTTACAACACTACAAAAGGAATACAAAAATAAATAT 60
LP56            GTGAACTTATATCAAACAAAACTTTTTACAATACTACAAAAGGAATACAAAAATAAATAT 60
                ******************************* ****************************

CP32-1          GGAGTTGATATATCACAATTTGTAAAGCTAACAAATTCTTCAATTAATTTTGATAAGTTT 120
CP32-8          GGAGTTGATATATCACAATTTGTAAAGCTAACAAATTCTTCAATTAATTTTGATAAGTTT 120
CP32-6          GGAGTTGATATATCACAATTTGTAAAGCTAACAAATTCTTCAATTAATTTTGATAAGTTT 120
CP32-4          GGAGTTGACATATCACAATTTGTAAAGCTAACAAATTCTTCAATTAATTTTGATAAGTTT 120
CP32-9          GGAGTTGACATATCACAATTTGTAAAGCTAGCAAATTCTTCAATTAATTTTGATAAGTTT 120
CP32-3          GGAGTTGATATATCACAATTTGTAAAGCTAACAAATTCTTCAATTAATTTTGATAAGTTT 120
CP32-7          GGAGTTGATATATCGCAATTTATAAAGCTAACAAATTCTTCAATTAATTTTGATAAGTTT 120
LP56            GGAGTTGATATATCACAATTTGTAAAGCTAACAAATTCTTCAATTAATTTTGATAAGTTT 120
                ******** ***** ****** ******** *****************************

CP32-1          GAAGAAGAACAGTTAACTTTAAAACAAAAAAATGTGATAAAAAGCATTAAAAAGAATAAT 180
CP32-8          GAAGAAGAACAGTTAACTTTAAAACAAAAAAATGTGATAAAAAGCATTAAAAAGAATAAT 180
CP32-6          GAAGAAAAACAGTTAACTTTAAAACAAAAAAATGTGATAAAAAGTATTAAAAAGAATAAT 180
CP32-4          GAAAAAGAACAGTTAACCATAAAACAAAAAAATGTTATAAAAAGTATTCAAAAAAATAAT 180
CP32-9          GAAGAAAAACAGTTAACTTTAAAACAAAAAAATGTGATAAAAAGTATTAAAAAGAATAAT 180
CP32-3          GAAGAAAAACAGTTAACTTTAAAACAAAAAAATGTGATAAAAAGTATTAAAAAGAATAAT 180
CP32-7          GAAGAAGAACAGCTAACTTTAAAACAAAAAAATGTGATAAAAAGCATTAAAAAGAATAAT 180
LP56            GAAGAAGAGCAGCTAACTTTAAAACAAAAAAATGTGATAAAAAGCATTAAAAAGAATAAT 180
                *** ** * *** ****  **************** ******** *** **** ******

CP32-1          GAAAAGAAGATTATACTCAGCGGAGGCATAGCTAGTGGCAAAACGTATCTTGCATGTTAT 240
CP32-8          GAAAAGAAGATTATACTCAGCGGAGGCATAGCTAGTGGCAAAACGTATCTTGCATGTTAT 240
CP32-6          GAAAAAAAGATTATACTCAGCGGCGGCATAGCTAGCGGCAAAACGTATCTTGCATGCTAT 240
CP32-4          GAAAAGAAGATTATACTCAGCGGCGGCATAGCTAGCGGCAAAACGTATCTTGCATGTTAT 240
CP32-9          GAAAAAAAGATTATACTCAGCGGAGGCATAGCTAGTGGCAAAACGTATCTTGCATGTTAT 240
CP32-3          GAAAAAAAGATTATATTCAGCGGCGGCATAGCTAGCGGCAAAACGTATCTTGCATGCTAT 240
CP32-7          GAAAAGAAGATTATACTCAGTGGAGGTATAGCTAGCGGCAAAACGTATCTTGCATGTTAT 240
LP56            GAAAAAAAGATTATACTCAGCGGCGGCATAGCTAGCGGCAAAACGTATCTTGCATGTTAT 240
                ***** ********* **** ** ** ******** ******************** ***

CP32-1          CTTTTTCTAAAAAGTTTAATTGAAATTAAAAAGTTATACTCTAGTGATACTAATAATTTC 300
CP32-8          CTTTTTCTAAAAAGTTTAATTGAAAATAAAAAGTTATACTCTAGTGATACTAATAATTTC 300
CP32-6          CTTTTTCTCAAAAGTTTAATTGAAAATAAAAAGTTATATTCTAGCGATACGAATAATTTT 300
CP32-4          CTTTTTCTCAAAAGTTTAATTGAAAATAAAAAGTTATACTCTGGTGATATTAATAATTTT 300
CP32-9          CTTTTTCTAAAAAGTTTAATTGAAAATAAAAAGTTATACTCTAGTGATACTAATAATTTC 300
CP32-3          CTTTTTCTCAAAAGTTTAATTGAAAATAAAAAGTTATATTCTAGCGATACGAATAATTTT 300
CP32-7          CTTTTTCTCAAAAGTTTAATTGAAAATAAAAAGCTATATTCTAGCGATACGAATAATTTC 300
LP56            CTTTTTATAAAAAGTTTAATTGAAAATAAAAAGCTATATTCTAGCGATACGAATAATTTC 300
                ****** * **************** ******* **** *** * ****  ********

CP32-1          ATTATAGGGAATTCACAACGTTCAGTTGAAGTTAATGTTTTGGGGCAATTTGAAAAGCTA 360
CP32-8          ATTATAGGGAATTCACAACGTTCAGTTGAAGTTAATGTTTTGGGGCAATTTGAAAAGCTA 360
CP32-6          ATTATTGGGAATTCACAACGCTCAGTTGAAGTTAATGTTTTGGGACAATTTGAAAAGCTA 360
CP32-4          ATTATTGGGAATTCACAACGCTCAGTTGAAGTTAATGTTTTGGGACAATTTGAAAAGCTA 360
CP32-9          ATTATTGGGAATTCACAACGCTCAGTTGAAGTTAATGTTTTAGGACAATTTGAAAAGCTA 360
CP32-3          ATTATTGGGAATTCACAACGCTCAGTTGAAGTTAATGTTTTGGGACAATTTGAAAAGCTA 360
CP32-7          ATTATAGGGAATTCACAACGTTCAGTTGAAGTTAATGTTTTGGGACAATTTGAAAAGCTA 360
LP56            ATTATAGGGAATTCACAACGCTCAGTTGAAGTTAATGTTTTGGGGCAATTTGAAAAGCTA 360
                ***** ************** ******************** ** ***************

CP32-1          TGTAAACTTCTTAAAATTCCTTATATTCCAAGACATACAAATAATTCATATATTCTGATT 420
CP32-8          TGTAAACTTCTTAAAATTCCTTATATTCCAAGACATACAAATAATTCATATATTCTGATT 420
CP32-6          TGTAAACTTCTTAAAATTCCTTATATTCCAAGACATACAAATAATTCATATATTCTGATT 420
CP32-4          TGTAAACTTCTTAAAATTCCTTATATCCCAAGACATACAAATAATTCATATATATTAATT 420
CP32-9          TGTAAACTTCTTAAAATTCCTTATATCCCAAGACATACAAATAATTCATATATTCTGATT 420
CP32-3          TGTAAACTTCTTAAAATTCCTTATATTCCAAGACATACAAATAATTCATATATTCTGATT 420
CP32-7          TGTAAACTTCTTAAAATTCCTTATATTCCAAGACATACAAATAATTCATATATTTTGATT 420
LP56            TGCAAACTGCTTAAAATTCCTTATATTCCAAGACATACAAATAATTCATATATTCTGATT 420
                ** ***** ***************** **************************  * ***

CP32-1          GATTCACTACGTATTAATCTATATGGAGGAGATAAGGCAAGTGATTTTGAAAGATTTAGG 480
CP32-8          GATTCACTACGTATTAATCTATATGGAGGAGATAAGGCAAGTGATTTTGAAAGATTTAGG 480
CP32-6          GATTCACTACGTATTAATCTATATGGTGGAGATAAGGCAAGTGATTTTGAAAGATTTAGG 480
CP32-4          GATTCACTTCGTATTAATCTATATGGAGGAGATAAGGCAAGTGATTTTGAAAGATTTAGA 480
CP32-9          GATTCACTACGTATTAATCTATATGGTGGAGATAAGGCAAGTGATTTTGAAAGATTTAGG 480
CP32-3          GATTCACTACGTATTAATCTATATGGTGGAGATAAGGCAAGTGATTTTGAAAGATTTAGG 480
CP32-7          GATTCACTACGTATTAATCTATATGGTGGAGATAAGGCAAGTGATTTTGAAAGATTTAGG 480
LP56            GATTCACTGCGAATTAATCTACATGGTGGAGATAAGGCAAGTGATTTTGAAAGATTTAGG 480
                ******** ** ********* **** ********************************

CP32-1          GGAAGTAATTCGGCACTTATTTTTGTTAATGAGGCTACAACTTTACACAAGCAAACTTTA 540
CP32-8          GGAAGTAATTCGGCACTTATTTTTGTTAATGAGGCTACAACTTTACACAAGCAAACTTTA 540
CP32-6          GGAAGTAATTCGGCACTTATTTTTGTTAATGAGGCTACAACTTTACACAAGCAAACTTTA 540
CP32-4          GGCAGTAATTCGGCACTTATTTTTGTTAATGAGGCTACTACTTTACACAAGCAAACTTTA 540
CP32-9          GGAAGTAATTCAGCACTTATTTTTGTTAATGAGGCTACAACTTTACACAAGCAAACTTTA 540
CP32-3          GGAAGTAATTCGGCACTTATTTTTGTTAATGAGGCTACAACTTTACACAAGCAAACTTTA 540
CP32-7          GGAAGTAATTCAGCACTTATTTTTGTGAATGAGGCTACAACTTTACACAAGCAAACTTTA 540
LP56            GGAAGTAATTCAGCACTTATTTTTGTTAATGAGGCTACAACTTTACACAAGCAAACTTTA 540
                ** ******** ************** *********** *********************

CP32-1          GAGGAAGTCTTAAAAAGACTAAGATGCGGGCAAGAAACTATTATTTTTGATACTAATCCT 600
CP32-8          GAGGAAGTCTTAAAAAGACTAAGATGCGGGCAAGAAACTATTATTTTTGATACTAATCCT 600
CP32-6          GAGGAGGTCTTAAAAAGACTAAGATGCGGGCAAGAAACTATTATTTTTGATACTAATCCC 600
CP32-4          GAGGAGGTCTTAAAAAGACTTAGGTGCGGACAAGAAACTATTATTTTTGATACTAATCCT 600
CP32-9          GAGGAGGTCTTAAAAAGACTAAGATGCGGGCAAGAAACTATTATTTTTGATACTAATCCT 600
CP32-3          GAGGAGGTCCTAAAAAGACTAAGATGCGGGCAAGAAACTATTATTTTTGATACTAATCCT 600
CP32-7          GAGGAAGTCTTAAAAAGACTAAGGTGTGGGCAAGAAACTATTATTTTTGATACTAACCCC 600
LP56            GAGGAAGTCTTAAAAAGATTAAGATGCGGGCAAGAAACTATTATTTTTGATACTAATCCT 600
                ***** *** ******** * ** ** ** ************************** **

CP32-1          GATCATCCAGAACACTATTTTAAAACCGATTATATTGATAATATAGCGACCTTTAAGACA 660
CP32-8          GATCATCCAGAACACTATTTTAAAACCGATTATATTGATAATATAGCGACCTTTAAGACA 660
CP32-6          GATCATCCAGAACACTATTTTAAAACCGATTATATTGATAATATAGCGACCTTTAAGACA 660
CP32-4          GATCATCCAGAACACTATTTTAAAACCGATTATATTGATAATATAGCGACATTTAAGACA 660
CP32-9          GATCATCCAGAACACTATTTTAAAACCGATTATATTGATAATATAGCGACATTTAAGATA 660
CP32-3          GATCATCCAGAACACTATTTTAAAACCGATTATATTGATAATATAGCGACATTTAAGACA 660
CP32-7          GATCATCCGGAACACTATTTTAAAACCGATTATATTGATAATATAGCAACTTTTAAGACA 660
LP56            GATCATCCAGAACACTATTTTAAAACCGATTATATTGATAATATAGCGACCTTTAAGACA 660
                ******** ************************************** ** ******* *

CP32-1          TATAAGTTTACAACTTATGATAATGTGCTACTTAGTAAAGGATTTGTCGAAACACAAGAA 720
CP32-8          TATAA-TTTACAACTTATGATAATGTGCTACTTAGTAAAGGATTTGTCGAAACACAAGAA 719
CP32-6          TATAATTTTACAACTTATGATAATGTGCTACTTAGTAAAGGATTTGTCGAAACACAAGAA 720
CP32-4          TATAATTTTACAACTTATGATAATGTGCTACTTAGTAAAGGATTTATCGAAACACAAGAA 720
CP32-9          TATAATTTTACAACTTATGATAATGTGCTACTTAGTAAAGGATTTATCGAAACACAAGAA 720
CP32-3          TATAATTTTACAACTTATGATAATGTTCTACTTAGTAAAGGATTTATCGAAACACAAGAA 720
CP32-7          TATAATTTTACAACTTATGATAATGTTCTACTTAGCAAAGGATTTATCGAAACTCAAGAA 720
LP56            TATAATTTTACAACTTATGATAATGTTCTACTTAGTAAAGGATTTATCGAAACACAAGAA 720
                ***** ******************** ******** ********* ******* ******

CP32-1          AAGCTATATAAAGATATACCATCATATAAAGCAAGAGTTTTGTTAGGTGAGTGGATAGCA 780
CP32-8          AAGCTATATAAAGATATACCATCATATAAAGCAAGAGTTTTGTTAGGTGAGTGGATAGCA 779
CP32-6          AAGCTATATAAAGATATACCATCATATAAAGCAAGAGTTTTGCTAGGTGAGTGGATAGCA 780
CP32-4          AAACTCTATAAAGATATACCATCATATAAAGCAAGAGTTTTGCTAGGTGAGTGGATAGCA 780
CP32-9          AAACTCTATAAAGATATACCATCATATAAAGCAAGAGTTTTGCTAGGTGAGTGGATAGCA 780
CP32-3          AAACTCTATAAAGATATACCATCATATAAAGCAAGAGTTTTGCTAGGTGAGTGGATAGCA 780
CP32-7          AAGCTATATAAAGATATACCATCATATAAAGCAAGAGTTTTGCTAGGTGAATGGATAGCA 780
LP56            AAGCTCTATAAAGATATACCATCATATAAAGCAAGAGTTTTGCTAGGGGAGTGGATAGCA 780
                ** ** ************************************ **** ** *********

CP32-1          AGCACTGATTCAATTTTTACACAAATAAATATTACTGATGATTATGTATTTACTAGCCCG 840
CP32-8          AGCACTGATTCAATTTTTACACAAATAAATATTACTGATGATTATGTATTTACTAGCCCG 839
CP32-6          AGCACTGATTCAATTTTTACACAAATAAATATTACTGATGATTATGTATTTACTAGCCCG 840
CP32-4          AGCACCGATTCAATTTTTACACAAATAAATATTACTAATGATTATGTATTTACTAGCCCG 840
CP32-9          AGCACTGATTCAATTTTTACACAAATAAATATTACTGATGATTATATATTTACTAGCCCG 840
CP32-3          AGCACTGATTCAATTTTTACACAAATAAATATTACTGATGATTATGTATTTACTAGTCCA 840
CP32-7          AGCACCGATTCAATTTTTACACAAATAAATATTACTAATGATTATGTATTTACTAGCCCG 840
LP56            AGCACCGATTCAATTTTTACACAAATAAATATTACTAATGATTATGTATTTACTAGCCCG 840
                ***** ****************************** ******** ********** **

CP32-1          ATAGCATATTTAGACCCAGCATTTAGTGTTGGCGGGGATAACACTGCATTATGTGTTATG 900
CP32-8          ATAGCATATTTAGACCCAGCATTTAGTGTTGGAGGGGATAACACTGCATTATGTGTTATG 899
CP32-6          ATAGCATATTTAGACCCAGCATTTAGTGTTGGCGGGGATAACACTGCATTATGTGTTATG 900
CP32-4          ATAGCATATTTAGACCCAGCATTTAGTGTTGGAGGGGATAACACTGCATTATGTGTTATG 900
CP32-9          ATAGCATATTTAGACCCAGCATTTAGTGTTGGAGGGGATAACACTGCATTATGTGTTATG 900
CP32-3          ATAGCATATTTAGACCCAGCATTTAGTGTTGGAGGAGATAACACTGCATTATGTGTTATG 900
CP32-7          ATAGCATATTTAGACCCAGCATTTAGTGTTGGAGGGGATAACACTGCATTATGTGTTATG 900
LP56            ATAGCATATTTAGACCCAGCATTTAGTGTTGGCGGGGATAACACTGCATTATGTGTTATG 900
                ******************************** ** ************************

CP32-1          GAGCGAGTTGATGATAAGTATTATGCTTTTGTATTTCAAGACCAAAGACCAGCTAATGAT 960
CP32-8          GAGCGAGTTGATGATAAGTATTATGCTTTTGTATTTCAAGACCAAAGACCAGCTAATGAT 959
CP32-6          GAGCGAGTTGATGATAAGTATTATGCTTTTGTATTTCAAGACCAAAGACCAGCTAATGAT 960
CP32-4          GAGCGAGTTGATGATAAGTATTATGCTTTTGTATTTCAAGACCAACGACCAGCCAATGAT 960
CP32-9          GAGCGAGTTGATGATAAGTATTATGCTTTTGTATTTCAAGACCAACGACCAGCCAATGAT 960
CP32-3          GAGCGAGTTGATGATAAGTATTATGCTTTTGTATTTCAAGACCAACGACCAGCCAATGAT 960
CP32-7          GAGCGAGTTGATGATAAGTATTATGCTTTTGTATTTCAAGACCAAAGACCAGCCAATGAC 960
LP56            GAGCGGGTTGATGATAAGTATTATGCTTTTGTATTTCAAGACCAACGACCAGCCAATGAC 960
                ***** *************************************** ******* *****

CP32-1          CCTTATATTATGAATATGGTAAAGACTGTTATAGAAAATTTCAATGTGCATACACTGTAT 1020
CP32-8          CCTTATATTATGAATATGGTAAAGACTGTTATAGAAAATTTCAATGTGCATACACTGTAT 1019
CP32-6          CCTTATATTATGAATATGGTAAAGACTGTTATAGAAAATTTCAATGTGCATACACTGTAT 1020
CP32-4          CCTTATATTATGAATATGGTAAAGACCGTTATAGAAAATTTCAATGTGCATACACTGTAT 1020
CP32-9          CCTTATATTATGAATATGGTAAAGACCGTTATAGAAAATTTCAATGTGCATACACTGTAT 1020
CP32-3          CCTTATATTATGAATATGGTAAAGACCGTTATAGAAAATTTCAATGTGCATACACTGTAT 1020
CP32-7          CCGTATATTATGAATATGGTTAAGACCGTTTTAGAGAATTTTAATGTACATACACTTTAT 1020
LP56            CCGTATATTATGAATATGGTTAAGACCGTTTTAGAAAATTTTAATGTACATACATTTTAT 1020
                ** ***************** ***** *** **** ***** ***** ****** * ***

CP32-1          TTAGAGGATAGAGATAATACAAAAGGTGCTGGTGGATTGACCCGTGAATACATCTTGCTA 1080
CP32-8          TTAGAGGATAGAGATAATACAAAAGGTGCTGGTGGATTGACCCGTGAATACATCTTGCTA 1079
CP32-6          TTAGAGGATAGAGATAATACAAAAGGTGCTGGTGGATTGACCCGTGAATACATCTTGCTA 1080
CP32-4          TTAGAGGATAGAGATAATACAAAAGGTGCTGGTGGATTGACCCGCGAATACATCTTGCTA 1080
CP32-9          TTAGAGGATAGAGATAATACAAAAGGTGCTGGTGGATTGACCCGCGAATACATCTTGCTA 1080
CP32-3          TTAGAGGATAGAGATAATACAAAAGGTGCTGGTGGATTGACCCGCGAATACATCTTACTA 1080
CP32-7          TTAGAAGATAGAGACAATACAAAAGGTGCTGGTGGATTGACTCGTGAATACATGTTGCTA 1080
LP56            TTAGAAGATAGAGACAATACAAAAGGTGCTGGTGGATTGACCCGCGAATACATCTTGCTA 1080
                ***** ******** ************************** ** ******** ** ***

CP32-1          AGAAGTAATATAAGCCAATATTTTAGAATTGTTCCAGTTAAGCCAAAGTCTAATAAATTT 1140
CP32-8          AGAAGTAATATAAGCCAATATTTTAGAATTGTTCCAGTTAAGCCAAAGTCTAATAAATTT 1139
CP32-6          AGAAGTAATATAAGCCAATATTTTAGAATTGTTCCAGTTAAGCCAAAGTCTAATAAATTT 1140
CP32-4          AGAAATAATATAAGCCAATATTTTAGAATTGTTCCAGTTAAGCCAAAGTCTAATAAATTT 1140
CP32-9          AGAAATAATATAAGCCAATATTTTAGAATTGTTCCAGTTAAGCCAAAGTCTAATAAATTT 1140
CP32-3          AGAAATAATATAAGCCAATATTTTAGAATTGTTCCAGTTAAGCCAAAGTCTAATAAATTT 1140
CP32-7          AGAAATAATATGGGTCAATATTTTAGAATTGTTCCAGTTAAGCCAAAGTCTAATAAATTT 1140
LP56            AGAAATAATATGGGTCAATATTTTAGAATTGTTCCAGTTAAGCCAAAGTCTAATAAATTT 1140
                **** ******  * *********************************************

CP32-1          AGCAGAATAACAACGTTAATTACGCCGTTTACTTACAAAAAACTTTATATTACAAAGTAC 1200
CP32-8          AGCAGAATAACAACGTTAATTACGCCGTTTACTTACAAAAAACTTTATATTACAAAGTAC 1199
CP32-6          AGCAGAATAACAACGTTAATTACGCCGTTTACTTACAAAAAACTTTACATTACAAAGTAC 1200
CP32-4          AGCAGAATAACAACGGTAATTACGCCGTTTACTTATAAGAAACTTTACATTACAAAGTAC 1200
CP32-9          AGCAGAATAACAATGTTAATTACGCCGTTTACTTACAAAAAACTTTATATTACAAAGTAC 1200
CP32-3          AGCAGAATAACAACGTTAATTACGCCGTTTACTTATAAGAAACTTTACATTACAAAGTAC 1200
CP32-7          AGCAGAATAACAACGTTAATTACGCCGTTTACTTATAAGAAACTTTACATTACAAAGTAC 1200
LP56            AGCAGAATAACAGCGTTAATTACGCCGTTTATTTATAAGAAACTGTACATTACGAAGTAC 1200
                ************  * *************** *** ** ***** ** ***** ******

CP32-1          AGTAGTTCTTCCGTATTTAATGATATTTATTCGTATAAGGGGGATAATAAAACCCATGAT 1260
CP32-8          AGTAGTTCTTCCGTATTTAATGATATTTATTCGTATAAGGGGGATAATAAAACCCATGAT 1259
CP32-6          AGTAGTTCTTCTGTATTTAATGATATTTATTCGTATAAGGGGGATAGCAAAACCCATGAT 1260
CP32-4          AGTAGTTCTTCTGTATTTAATGATATTTATTCGTATAAGGGGGATAGCAAAACCCATGAT 1260
CP32-9          AGTAGTTCTTCTGTATTTAATGATATTTATTCGTATAAGGGGGATAGCAAAACCCATGAT 1260
CP32-3          AGTAGTTCTTCTGTATTTAATGATATTTATTCGTATAAGGGGGATAGCAAAACCCATGAT 1260
CP32-7          AGCAGTTCTTCTGTATTTAATGATATTTATTCGTATAAAGGAGATAACAAAACCCATGAT 1260
LP56            AGCAGTTCTTCTGTATTTAATGATATTTATTCGTATAAAGGAGATAACAAAACCCATGAT 1260
                ** ******** ************************** ** ****  ************

CP32-1          GACGCTCTTGATGCAATATCTGCAGCATATTTGATGTTGTCTTTAGGATATAGAGAGCGA 1320
CP32-8          GACGCTCTTGATGCAATATCTGCAGCATATTTGATGTTGTCTTTAGGATATAGAGAGCGA 1319
CP32-6          GATGCTCTTGATGCAATGTCTGCAGCATATTTGATGTTGTCTTTAGGATATAGAGAGCGA 1320
CP32-4          GATGCTCTTGATGCAATGTCTGCAGCATATTTGATGTTGTCTTTAGGATATAGAGAGCGA 1320
CP32-9          GATGCTCTTGATGCAATATCTGCAGCATATTTGATGTTGTCTTTAGGATATAGAGAGCGA 1320
CP32-3          GATGCTCTTGATGCAATGTCTGCAGCATATTTGATGTTGTCTTTAGGATATAGAGAGCGA 1320
CP32-7          GATGCTCTTGATGCAATATCTGCAGCATATTTGATGTTGTCTTTAGGGTATAGAGAGAGA 1320
LP56            GATGCTCTTGATGCAATATCTGCAGCATATTTGATGTTGTCTTTAGGGTATAGAGAGAGA 1320
                ** ************** ***************************** ********* **

CP32-1          AGTGTTCACTTTGGCAATCAAAGATTTTTGTAA 1353
CP32-8          AGTGTTCACTTTGGCAATCAAAGATTTTTGTAA 1352
CP32-6          AGTGTTCACTTTGGCAATCAAAGATTTTTGTAA 1353
CP32-4          AGTGTTCACTTTGGCAATCAAAGATTTTTGTAA 1353
CP32-9          AGTGTTCACTTTGGCAATCAAAGATTTTTGTAA 1353
CP32-3          AGTGTTCACTTTGGCAATCAAAGATTTTTGTAA 1353
CP32-7          AGTGTTCACTTTGGCAATCAAAGATTTTTGTAA 1353
LP56            AGTGTTCACTTTGGCAATCAAAGATTTTTGTAA 1353
                *********************************
```

**Supplementary information 2: the *terL* nucleotide sequence from 13 cp32 plasmids present within *Borrelia burgdorferi* species**

13 versions of the *terL* genes are discovered from 13 cp32 plasmids. The alignment shows that they are similar to each other, which allows primers (yellow and green) and probe (purple) to be designed in the conserved regions of nine out of 13 terminase genes (except cp32-2, cp32-3, cp32-6 and cp32-13) with the help of PrimerQuest® Tool (IDT).

```
>cp32-1 AE001575.1:29398-30750 Borrelia burgdorferi B31 plasmid cp32-1, complete sequence
GTGAACTTATATCAAACAAAACTTTTTACAACACTACAAAAGGAATACAAAAATAAATATGGAGTTGATATATCACAATT
TGTAAAGCTAACAAATTCTTCAATTAATTTTGATAAGTTTGAAGAAGAACAGTTAACTTTAAAACAAAAAAATGTGATAA
AAAGCATTAAAAAGAATAATGAAAAGAAGATTATACTCAGCGGAGGCATAGCTAGTGGCAAAACGTATCTTGCATGTTAT
CTTTTTCTAAAAAGTTTAATTGAAATTAAAAAGTTATACTCTAGTGATACTAATAATTTCATTATAGGGAATTCACAACG
TTCAGTTGAAGTTAATGTTTTGGGGCAATTTGAAAAGCTATGTAAACTTCTTAAAATTCCTTATATTCCAAGACATACAA
ATAATTCATATATTCTGATTGATTCACTACGTATTAATCTATATGGAGGAGATAAGGCAAGTGATTTTGAAAGATTTAGG
GGAAGTAATTCGGCACTTATTTTTGTTAATGAGGCTACAACTTTACACAAGCAAACTTTAGAGGAAGTCTTAAAAAGACT
AAGATGCGGGCAAGAAACTATTATTTTTGATACTAATCCTGATCATCCAGAACACTATTTTAAAACCGATTATATTGATA
ATATAGCGACCTTTAAGACATATAAGTTTACAACTTATGATAATGTGCTACTTAGTAAAGGATTTGTCGAAACACAAGAA
AAGCTATATAAAGATATACCATCATATAAAGCAAGAGTTTTGTTAGGTGAGTGGATAGCAAGCACTGATTCAATTTTTAC
ACAAATAAATATTACTGATGATTATGTATTTACTAGCCCGATAGCATATTTAGACCCAGCATTTAGTGTTGGCGGGGATA
ACACTGCATTATGTGTTATGGAGCGAGTTGATGATAAGTATTATGCTTTTGTATTTCAAGACCAAAGACCAGCTAATGAT
CCTTATATTATGAATATGGTAAAGACTGTTATAGAAAATTTCAATGTGCATACACTGTATTTAGAGGATAGAGATAATAC
AAAAGGTGCTGGTGGATTGACCCGTGAATACATCTTGCTAAGAAGTAATATAAGCCAATATTTTAGAATTGTTCCAGTTA
AGCCAAAGTCTAATAAATTTAGCAGAATAACAACGTTAATTACGCCGTTTACTTACAAAAAACTTTATATTACAAAGTAC
AGTAGTTCTTCCGTATTTAATGATATTTATTCGTATAAGGGGGATAATAAAACCCATGATGACGCTCTTGATGCAATATC
TGCAGCATATTTGATGTTGTCTTTAGGATATAGAGAGCGAAGTGTTCACTTTGGCAATCAAAGATTTTTGTAA

>cp32-5 CP019849.1:29322-30674 Borreliella burgdorferi plasmid cp32-5, complete sequence
GTGAACTTATATCAAACAAAACTTTTTACAACACTACAAAAGGAATACAAAAATAAATATGGAGTTGATATATCACAATT
TGTAAAGCTAACAAATTCTTCAATTAATTTTGATAAGTTTGAAGAAGAACAGTTAACTTTAAAACAAAAAAATGTGATAA
AAAGCATTAAAAAGAATAATGAAAAGAAGATTATACTCAGCGGAGGCATAGCTAGTGGCAAAACGTATCTTGCATGTTAT
CTTTTTCTAAAAAGTTTAATTGAAAATAAAAAGTTATACTCTAGTGATACTAATAATTTCATTATAGGGAATTCACAACG
TTCAGTTGAAGTTAATGTTTTGGGGCAATTTGAAAAGCTATGTAAACTTCTTAAAATTCCTTATATTCCAAGACATACAA
ATAATTCATATATTCTGATTGATTCACTACGTATTAATCTATATGGAGGAGATAAGGCAAGTGATTTTGAAAGATTTAGG
GGAAGTAATTCGGCACTTATTTTTGTTAATGAGGCTACAACTTTACACAAGCAAACTTTAGAGGAAGTCTTAAAAAGACT
AAGATGCGGGCAAGAAACTATTATTTTTGATACTAATCCTGATCATCCAGAACACTATTTTAAAACCGATTATATTGATA
ATATAGCGACCTTTAAGACATATAAGTTTACAACTTATGATAATGTGCTACTTAGTAAAGGATTTGTCGAAACACAAGAA
AAGCTATATAAAGATATACCATCATATAAAGCAAGAGTTTTGTTAGGTGAGTGGATAGCAAGCACTGATTCAATTTTTAC
ACAAATAAATATTACTGATGATTATGTATTTACTAGCCCGATAGCATATTTAGACCCAGCATTTAGTGTTGGCGGGGATA
ACACTGCATTATGTGTTATGGAGCGAGTTGATGATAAGTATTATGCTTTTGTATTTCAAGACCAAAGACCAGCTAATGAT
CCTTATATTATGAATATGGTAAAGACTGTTATAGAAAATTTCAATGTGCATACACTGTATTTAGAGGATAGAGATAATAC
AAAAGGTGCTGGTGGATTGACCCGTGAATACATCTTGCTAAGAAGTAATATAAGCCAATATTTTAGAATTGTTCCAGTTA
AGCCAAAGTCTAATAAATTTAGCAGAATAACAACGTTAATTACGCCGTTTACTTACAAAAAACTTTATATTACAAAGTAC
AGTAGTTCTTCCGTATTTAATGATATTTATTCGTATAAGGGGGATAATAAAACCCATGATGACGCTCTTGATGCAATATC
TGCAGCATATTTGATGTTGTCTTTAGGATATAGAGAGCGAAGTGTTCACTTTGGCAATCAAAGATTTTTGTAA

>cp32-8 AE001580.1:29533-30885 Borrelia burgdorferi B31 plasmid cp32-8, complete sequence
GTGAACTTATATCAAACAAAACTTTTTACAACACTACAAAAGGAATACAAAAATAAATATGGAGTTGATATATCACAATT
TGTAAAGCTAACAAATTCTTCAATTAATTTTGATAAGTTTGAAGAAGAACAGTTAACTTTAAAACAAAAAAATGTGATAA
AAAGCATTAAAAAGAATAATGAAAAGAAGATTATACTCAGCGGAGGCATAGCTAGTGGCAAAACGTATCTTGCATGTTAT
CTTTTTCTAAAAAGTTTAATTGAAAATAAAAAGTTATACTCTAGTGATACTAATAATTTCATTATAGGGAATTCACAACG
TTCAGTTGAAGTTAATGTTTTGGGGCAATTTGAAAAGCTATGTAAACTTCTTAAAATTCCTTATATTCCAAGACATACAA
ATAATTCATATATTCTGATTGATTCACTACGTATTAATCTATATGGAGGAGATAAGGCAAGTGATTTTGAAAGATTTAGG
GGAAGTAATTCGGCACTTATTTTTGTTAATGAGGCTACAACTTTACACAAGCAAACTTTAGAGGAAGTCTTAAAAAGACT
AAGATGCGGGCAAGAAACTATTATTTTTGATACTAATCCTGATCATCCAGAACACTATTTTAAAACCGATTATATTGATA
ATATAGCGACCTTTAAGACATATAAGTTTACAACTTATGATAATGTGCTACTTAGTAAAGGATTTGTCGAAACACAAGAA
AAGCTATATAAAGATATACCATCATATAAAGCAAGAGTTTTGTTAGGTGAGTGGATAGCAAGCACTGATTCAATTTTTAC
ACAAATAAATATTACTGATGATTATGTATTTACTAGCCCGATAGCATATTTAGACCCAGCATTTAGTGTTGGAGGGGATA
ACACTGCATTATGTGTTATGGAGCGAGTTGATGATAAGTATTATGCTTTTGTATTTCAAGACCAAAGACCAGCTAATGAT
CCTTATATTATGAATATGGTAAAGACTGTTATAGAAAATTTCAATGTGCATACACTGTATTTAGAGGATAGAGATAATAC
AAAAGGTGCTGGTGGATTGACCCGTGAATACATCTTGCTAAGAAGTAATATAAGCCAATATTTTAGAATTGTTCCAGTTA
AGCCAAAGTCTAATAAATTTAGCAGAATAACAACGTTAATTACGCCGTTTACTTACAAAAAACTTTATATTACAAAGTAC
AGTAGTTCTTCCGTATTTAATGATATTTATTCGTATAAGGGGGATAATAAAACCCATGATGACGCTCTTGATGCAATATC
TGCAGCATATTTGATGTTGTCTTTAGGATATAGAGAGCGAAGTGTTCACTTTGGCAATCAAAGATTTTTGTAA

>cp32-3 CP002269.1:28910-30262 Borrelia burgdorferi 297 plasmid 297_cp32-3, complete sequence
GTGAACTTATATCAAACAAAACTTTTTACAACACTACAAAAGGAATATAAAAATAAATATGGAGTTGACATATCACAATT
TGTAAAGCTAACAAATTCTTCAATTAATTTTGATAAGTTTGAAGAAAAACAGTTAACTTTAAAACAAAAAAATGTGATAA
AAAGCATTAAAAAGAATAATGAAAAGAAGATTATACTCAGCGGAGGCATAGCTAGTGGCAAAACGTATCTTGCATGTTAT
CTTTTTCTAAAAAGTTTAATTGAAAATAAAAAGTTATACTCTAGTGATACTAATAATTTTATTATTGGGAATTCACAACG
CTCAGTTGAAGTTAATGTTTTGGGACAATTTGAAAAGCTATGTAAACTTCTTAAAATTCCTTATATCCCAAGACATACAA
ATAATTCATATATTCTGATTGATTCACTACGTATTAATCTATATGGTGGAGATAAGGCAAGTGATTTTGAAAGATTTAGG
GGAAGTAATTCGGCACTTATTTTTGTTAATGAGGCTACAACTTTACACAAGCAAACTTTAGAGGAGGTCTTAAAAAGACT
AAGATGCGGGCAAGAAACTATTATTTTTGATACTAATCCTGATCATCCAGAACACTATTTTAAAACCGATTATATTGATA
ATATAGCGACCTTTAAGACATATAATTTTACAACTTATGATAATGTGCTACTTAGTAAAGGATTTGTCGAAACACAAGAA
AAGCTATATAAAGATATACCATCATATAAAGCAAGAGTTTTGTTAGGTGAGTGGATAGCAAGCACCGATTCAATTTTTAC
ACAAATAAATATTACTGATGATTATGTATTTACTAGCCCGATAGCATATTTAGACCCAGCATTTAGTGTTGGCGGGGATA
ACACTGCATTATGTGTTATGGAGCGAGTTGATGATAAGTATTATGCTTTTGTATTTCAAGACCAAAGACCAGCTAATGAT
CCTTATATTATGAATATGGTAAAGACTGTTATAGAAAATTTCAATGTGCATACACTGTATTTAGAGGATAGAGATAATAC
AAAAGGTGCTGGTGGATTGACCCGTGAATACATCTTGCTAAGAAGTAATATAAGCCAATATTTTAGAATTGTTCCAGTTA
AGCCAAAGTCTAATAAATTTAGCAGAATAACAACGTTAATTACGCCGTTTACTTACAAAAAACTTTATATTACAAAGTAC
AGTAGTTCTTCCGTATTTAATGATATTTATTCGTATAAGGGGGATAATAAAACCCATGATGATGCTCTTGATGCAATGTC
TGCAGCATATTTGATGTTGTCTTTAGGATATAGAGAGCGAAGTGTTCACTTTGGCAATCAAAGATTTTTGTAA

>cp32-4 CP001208.1:29612-30964 Borrelia burgdorferi ZS7 plasmid ZS7_cp32-4, complete sequence
GTGAACTTATATCAAACAAAACTTTTTACAACACTACAAAAGGAATATAAAAATAAATATGGAGTTGACATATCACAATT
TGTAAAGCTAACAAATTCTTCAATTAATTTTGATAAGTTTGAAGAAAAACAGTTAACTTTAAAACAAAAAAATGTGATAA
AAAGCATTCAAAAGAATAATGAAAAGAAGATTATACTCAGCGGAGGCATAGCTAGTGGCAAAACGTATCTTGCATGTTAT
CTTTTTCTAAAAAGTTTAATTGAAAATAAAAAGTTATACTCTAGTGATACTAATAATTTTATTATTGGGAATTCACAACG
CTCAGTTGAAGTTAATGTTTTGGGACAATTTGAAAAGCTATGTAAACTTCTTAAAATTCCTTATATCCCAAGACATACAA
ATAATTCATATATTCTGATTGATTCACTACGTATTAATCTATATGGTGGAGATAAGGCAAGTGATTTTGAAAGATTTAGG
GGAAGTAATTCGGCACTTATTTTTGTTAATGAGGCTACAACTTTACACAAGCAAACTTTAGAGGAAGTCTTAAAAAGACT
AAGATGCGGGCAAGAAACTATTATTTTTGATACTAATCCTGATCATCCAGAACACTATTTTAAAACCGATTATATTGATA
ATATAGCGACCTTTAAGACATATAAGTTTACAACTTATGATAATGTGCTACTTAGTAAAGGATTTGTCGAAACACAAGAA
AAGCTATATAAAGATATACCATCATATAAAGCAAGAGTTTTGTTAGGTGAGTGGATAGCAAGCACTGATTCAATTTTTAC
ACAAATAAATATTACTGATGATTATGTATTTACTAGCCCGATAGCATATTTAGACCCAGCATTTAGTGTTGGCGGGGATA
ACACTGCATTATGTGTTATGGAGCGAGTTGATGATAAGTATTATGCTTTTGTATTTCAAGACCAAAGACCAGCTAATGAT
CCTTATATTATGAATATGGTAAAGACTGTTATAGAAAATTTCAATGTGCATACACTGTATTTAGAGGATAGAGATAATAC
AAAAGGTGCTGGTGGATTGACCCGTGAATACATCTTGCTAAGAAGTAATATAAGCCAATATTTTAGAATTGTTCCAGTTA
AGCCAAAGTCTAATAAATTTAGCAGAATAACAACGTTAATTACGCCGTTTACTTACAAAAAACTTTATATTACAAAGTAC
AGTAGTTCTTCTGTATTTAATGATATTTATTCGTATAAGGGGGATAGCAAAACCCATGATGATGCTCTTGATGCAATGTC
TGCAGCATATTTGATGTTGTCTTTAGGATATAGAGAGCGAAGTGTTCACTTTGGCAATCAAAGATTTTTGTAA

>cp32-7 CP017209.1:28702-30054 Borreliella burgdorferi strain B331 plasmid B331_cp32_7
GTGAACTTATATCAAACAAAACTTTTTACAACACTACAAAAGGAATACAAAAATAAATATGGAGTTGACATATCACAATT
TGTAAAGCTAACAAATTCTTCAATTAATTTTGATAAGTTTGAAGAAAAACAGTTAACTTTAAAACAAAAAAATGTGATAA
AAAGCATTCAAAAGAATAATGAAAAGAAGATTATACTCAGCGGAGGCATAGCTAGTGGCAAAACGTATCTTGCATGTTAT
CTTTTTCTAAAAAGTTTAATTGAAAATAAAAAGTTATACTCTAGTGATACTAATAATTTTATTATTGGGAATTCACAACG
CTCAGTTGAAGTTAATGTTTTGGGACAATTTGAAAAGCTATGTAAACTTCTTAAAATTCCTTATATCCCAAGACATACAA
ATAATTCATATATTCTGATTGATTCACTACGTATTAATCTATATGGTGGAGATAAGGCAAGTGATTTTGAAAGATTTAGG
GGAAGTAATTCAGCACTTATTTTTGTTAATGAGGCTACAACTTTACACAAGCAAACTTTAGGGGAGGTCTTAAAAAGACT
AAGATGCGGGCAAGAAACTATTATTTTTGATACCAATCCTGATCATCCAGAACACTATTTTAAAACCGATTATATTGATA
ATATAGCGACCTTTAAGACATATAATTTTACAACTTATGATAATGTGCTACTTAGTAAAGGATTTGTCGAAACACAAGAA
AAGCTATATAAAGATATACCATCATATAAAGCAAGAGTTTTGCTAGGTGAGTGGATAGCAAGCACTGATTCAATTTTTAC
ACAAATAAATATTACTGATGATTATGTATTTACTAGCCCAATAGCATATTTAGACCCAGCATTTAGTGTTGGAGGAGATA
ACACTGCATTATGTGTTATGGAGCGAGTTGATGATAAGTATTATGCTTTTGTATTTCAAGACCAACGACCGGCCAATGAT
CCTTATATTATGAATATGGTAAAGACTGTTATAGAAAATTTCAATGTGCATACACTGTATTTAGAGGATAGAGATAATAC
AAAAGGTGCTGGTGGATTGACCCGTGAATACATCTTGCTAAGAAGTAATATAAGCCAATATTTTAGAATTGTTCCAGTTA
AGCCAAAGTCTAATAAATTTAGCAGAATAACAACGTTAATTACGCCGTTTACTTACAAAAAACTTTATATTACAAAGTAC
AGTAGTTCTTCTGTATTTAATGATATTTATTCGTATAAGGGGGATAGCAAAACCCATGATGATGCTCTTGATGCAATGTC
TGCAGCATATTTGATGTTGTCTTTAGGATATAGAGAGCGAAGTGTTCACTTTGGCAATCAAAGATTTTTGTAA

>cp32-11 CP001426.1:28908-30260 Borrelia burgdorferi 64b plasmid 64b_cp32-11, complete sequence
GTGAACTTATATCAAACAAAACTTTTTACAACACTACAAAAGGAATACAAAAATAAATATGGAGTTGATATATCACAATT
TGTAAAGCTAACAAATTCTTCAATTAATTTTGATAAGTTTGAAGAAGAACAGTTAACTTTAAAACAAAAAAATGTGATAA
AAAGCATTAAAAAGAATAATGAAAAGAAGATTATACTCAGTGGAGGTATAGCTAGTGGCAAAACGTATCTTGCATGTTAT
CTTTTTCTAAAAAGTTTAATTAAAAATAAAAAGTTATACTCTAGTGATACTAATAATTTCATTATAGGGAATTCACAACG
TTCAGTTGAAGTTAATGTTTTGGGGCAATTTGAAAAGCTATGTAAACTTCTTAAAATTCCTTATATTCCAAGACATACAA
ATAATTCATATATTCTGATTGATTCACTGCGAATTAATCTATATGGTGGAGATAAGGCAAGTGATTTTGAAAGATTTAGG
GGAAGTAATTCAGCACTTATTTTTGTTAATGAGGCTACAACTTTACACAAGCAAACTTTAGAGGAGGTCTTAAAAAGACT
AAGATGCGGGCAAGAAACTATTATTTTTGATACTAATCCCGATCATCCAGAACACTATTTTAAAACCGATTATATTGATA
ATATAGCGACCTTTAAGACATATAATTTTACAACTTATGATAATGTGCTACTTAGTAAAGGATTTGTCGAAACACAAGAA
AAGCTATATAAAGATATACCATCATATAAAGCAAGAGTTTTGCTAGGTGAGTGGATAGCAAGCACTGATTCAATTTTTAC
ACAAATAAATATTACTGATGATTATGTATTTACTAGCCCGATAGCATATTTAGACCCAGCATTTAGTGTTGGCGGGGATA
ACACTGCATTATGTGTTATGGAGCGAGTTGATGATAAGTATTATGCTTTTGTATTTCAAGACCAAAGACCAGCTAATGAT
CCTTATATTATGAATATGGTAAAGACCGTTATAGAAAATTTCAATGTGCATACACTGTATTTAGAGGATAGAGATAATAC
AAAAGGTGCTGGTGGATTGACCCGTGAATACATCTTGCTAAGAAGTAATATAAGCCAATATTTTAGAATTGTTCCAGTTA
AGCCAAAGTCTAATAAATTTAGCAGAATAACAACGTTAATTACGCCGTTTACTTACAAAAAACTTTATATTACAAAGTAC
AGTAGTTCTTCTGTATTTAATGATATTTATTCGTATAAGGGGGATAGCAAAACCCATGATGATGCTCTTGATGCAATGTC
TGCAGCATATTTGATGTTGTCTTTAGGATATAGAGAGCGAAGTGTTCACTTTGGCAATCAAAGATTTTTGTAA

>cp32-6 CP001427.1:29403-30755 Borrelia burgdorferi 64b plasmid 64b_cp32-6, complete sequence
GTGAACTTATATCAAACAAAACTTTTTACAACACTACAAAAGGAATATAAAAATAAATATGGAGTTGACATATCACAATT
TGTAAAGCTAACAAATTCTTCAATTAATTTTGATAAGTTTGAAGAAAAACAGTTAACTTTAAAACAAAAAAATGTGATAA
AAAGCATTCAAAAGAATAATGAAAAGAAGATTATACTCAGCGGAGGCATAGCTAGTGGCAAAACGTATCTTGCATGTTAT
CTTTTTCTAAAAAGTTTAATTGAAAATAAAAAGTTATACTCTAGTGATACTAATAATTTCATTATAGGGAATTCACAACG
TTCAGTTGAAGTTAATGTTTTGGGACAATTTGAAAAGCTATGTAAACTTCTTAAAATTCCTTATATCCCAAGACATACAA
ATAATTCATATATTCTGATTGATTCACTACGTATTAATCTATATGGTGGAGATAAGGCAAGTGATTTTGAAAGATTTAGG
GGAAGTAATTCGGCACTTATTTTTGTTAATGAGGCTACAACTTTACACAAGCAAACTTTAGAGGAAGTCTTAAAAAGACT
AAGATGCGGGCAAGAAACTATTATTTTTGATACTAATCCTGATCATCCAGAACACTATTTTAAAACCGATTATATTGATA
ATATAGCGACCTTTAAGACATATAATTTTACAACTTATGATAATGTGCTACTTAGTAAAGGATTTGTCGAAACACAAGAA
AAGCTATATAAAGATATACCATCATATAAAGCAAGAGTTTTGTTAGGTGAGTGGATAGCAAGCACTGATTCAATTTTTAC
ACAAATAAATATTACTGATGATTATGTATTTACTAGCCCAATAGCATATTTAGACCCAGCATTTAGTGTTGGAGGAGATA
ACACTGCATTATGTGTTATGGAGCGAGTTGATGATAAGTATTATGCTTTTGTATTTCAAGACCAACGACCAGCCAATGAT
CCTTATATTATGAATATCGTAAAGACTGTTATAGAAAATTTCAATGTGCATACACTGTATTTAGAGGATAGAGATAATAC
AAAAGGTGCTGGTGGATTGACCCGTGAATACATCTTACTAAGAAGTAATATAAGCCAATATTTTAGAATTGTTCCAGTTA
AGCCAAAGTCTAATAAATTTAGCAGAATAACAACGTTAATTACGCCGTTTACTTACAAAAAACTTTACATTACAAAGTAC
AGTAGTTCTTCTGTATTTAATGATATTTATTCGTATAAGGGGGATAATAAAACCCATGATGATGCTCTTGATGCAATGTC
TGCAGCATATTTGATGTTGTCTTTAGGATATAGAGAGCGAAGTGTTCACTTTGGCAATCAAAGATTTTTGTAA

>cp32-2 CP019919.1:29893-31245 Borreliella burgdorferi strain PAbe plasmid p_cp32-2, complete sequence
GTGAACTTATATCAAACAAAACTTTTTACAACACTACAAAAGGAATACAAAAATAAATATGGAGTTGATATATCACAATT
TGTAAAGCTAACAAATTCTTCAATTAATTTTGATAAGTTTGAAGAAGAACAGTTAACTTTAAAACAGAAAAATGTGATAA
AAAGCATTGAAAAGAATAATGAAAAGAAGATTATACTCAGTGGAGGTATAGCTAGCGGCAAAACGTATCTTGCATGTTAT
CTTTTTCTAAAAAGTTTAATTGAAAATAAAAAGCTATATTCTAGCGATACGAATAATTTCATTATAGGGAATTCACAACG
TTCAGTTGAAGTTAATGTTTTGGGACAATTTGAAAAGCTATGTAAACTTCTTAAAATTCCTTATATTCCAAGACATACAA
ATAATTCATATATTCTGATTGATTCACTGCGAATTAATCTATATGGTGGAGATAAGGCAAGTGTTTTTGAAAGATTTAGG
GGAAGTAATTCAGCACTTATTTTTGTGAATGAGGCTACAACTTTACACAAGCAAACTTTAGAGGAGGTCTTAAAAAGACT
AAGATGCGGGCAAGAAACTATTATTTTTGATACTAATCCTGATCATCCAGAACACTATTTTAAAACCGATTATATTGATA
ATATAGCGACCTTTAAGACATATAATTTCACAACTTATGATAATGTGCTACTTAGTAAAGGATTTGTCGAAACACAAGAA
AAGCTATATAAAGATATACCATCATATAAAGCAAGAGTTTTGCTAGGTGAATGGATAGCAAGCACCGATTCAATTTTTAC
ACAAATAAATATTACTGATGATTATGTATTTACTAGCCCGATAGCATATTTAGACCCAGCATTTAGTGTTGGAGGGGATA
ACACTGCATTATGTGTTATGGAGCGAATTGATGATAAGTATTATGCTTTTGTATTTCAAGACCAACGACCAGCCAATGAT
CCTTATATTATGAATATGGTAAAGACCGTTATAGAAAATTTCAATGTGCATACACTGTATTTAGAGGATAGAGATAATAC
AAAAGGTGCTGGTGGATTGACCCGTGAATACATCTTGCTAAGAAATAATACGAGTCAATATTTTAGAATTGTTCCAGTTA
AGCCAAAGTCTAATAAATTTAGCAGAATAACAACGTTAATTACGCCGTTTACTTATAAGAAACTTTACATTACAAAGTAC
AGTAGTTCTTCTGTATTTAATGATATTTATTCGTATAAGGGGGATAACAAAACCCATGATGATGCTCTTGATGCAATGTC
TGCAGCATATTTGATGTTGTCTTTAGGATATAGAGAGCGAAGTGTTCACTTTGGCAATCAAAGATTTTTGTAA

>cp32-12 CP001565.1:30081-31433 Borrelia burgdorferi Bol26 plasmid Bol26_cp32-12, complete sequence
GTGAACTTATATCAAACAAAACTTTTTACAACACTACAAAAGGAATACAAAAATAAATATGGAGTTGACATATCACAATT
TGTAAAGCTAACAAATTCTTCAATTAATTTTGATAAGTTTGAAGAAAAACAGTTAACTTTAAAACAAAAAAATGTGATAA
AAAGCATTAAAAAGAATAATGAAAAAAAGATTATACTCAGCGGCGGCATAGCTAGCGGCAAAACGTATCTTGCCTGTTAT
CTTTTTCTAAAAAGTTTAATTGCAAATAAGAATTTATATTCTAGTGATACGAATAATTTCATTATTGGCAATTCGCAGCG
CTCGGTTGAAGTTAATGTTTTGGGGCAATTTGAAAAGCTATGCAAACGGCTTAAAATTCCTTATATTCCAAGACATACAA
ATAATTCATATATTCTGATTGATTCACTACGTATTAATCTATATGGTGGAGATAAGGCAAGTGATTTTGAAAGATTTAGG
GGAAGTAATTCAGCGCTTATTTTTGTTAATGAGGCTACAACTTTACACAGGCAAACTTTAGAGGAAGTCTTAAAAAGACT
AAGGTGTGGGCAAGAAACTATTATTTTTGATACTAACCCCGATCATCCGGAACACTATTTTAAAACCGATTATATTGATA
ATATAGCAACTTTTAAGACATATAATTTTACAACTTATGATAATGTTCTACTTAGTAAAGGATTTATCGAAACACAAGAA
AAACTCTATAAAGATATACCATCATATAAAGCAAGAGTTTTGCTAGGTGAGTGGATAGCAAGCACTGATTCAATTTTTAC
ACAAATAAATATTACTGATGATTATGTATTTACTAGCCCGATAGCATATTTAGACCCAGCATTTAGTGTTGGCGGGGATA
ACACTGCATTATGTGTTATGGAGCGAGTTGATGATAAGTATTATGCTTTTGTATTTCAAGACCAACGACCAGCTAATGAT
CCTTATATTATGAATATGGTAAAGACCGTTATAGAAAATTTCAATGTGCATACACTGTATTTAGAGGATAGAGATAATAC
AAAAGGTGCTGGTGGATTGACCCGTGAATACATCTTGCTAAGAAATAATATAAGCCAATATTTTAGAATTGTTCCAGTTA
AGCCAAAGTCTAATAAATTTAGCAGAATAACAACGTTAATTACGCCGTTTACTTATAAGAAACTTTACATTACAAAGTAC
AGTAGTTCTTCTGTATTTAATGATATTTATTCGTATAAGGGGGATAACAAAACCCATGATGATGCTCTTGATGCAATGTC
TGCAGCATATTTGATGTTGTCTTTAGGATATAGAGAGCGAAGTGTTCACTTTGGCAACCAAAGATTTTTGTAA

>cp32-10 CP001449.1:28404-29756 Borrelia burgdorferi WI91-23 plasmid WI91-23_cp32-10, complete sequence
GTGAACTTATATCAAACAAAACTTTTTACAACACTACAAAAGGAATACAAAAATAAATATGGAGTTGATATATCACAATT
TGTAAAGCTAACAAATTCTTCAATTAATTTTGATAAGTTTGAAGAAGAACAGTTAACTTTAAAACAAAAAAATGTGATAA
AAAGCATTAAAAAGAATAATGAAAAAAAGATTATACTCAGCGGCGGCATAGCTAGCGGCAAAACGTATCTTGCATGTTAT
CTTTTTCTAAAAAGTTTAATTAAAAATAAAAAGTTATATTCTAGCGATACGAATAATTTTATTATTGGGAATTCACAACG
CTCAGTTGAAGTTAATGTTTTGGGACAATTTGAAAAGCTATGTAAACTTCTTAAAATTCCTTATATTCCAAGACATACAA
ATAATTCATATATTCTTATTGATTCACTGAGAATTAATCTATATGGAGGAGATAAGGCAAGTGATTTTGAAAGATTTAGG
GGAAGTAATTCGGCACTTATTTTTGTTAATGAGGCTACAACTTTACACAAGCAAACTTTAGAGGAGGTCTTAAAAAGACT
AAGATGCGGGCAAGAAACTATTATTTTTGATACTAATCCCGATCATCCAGAACACTATTTTAAAACCGATTATATTGATA
ATATAGCGACATTTAAGACATATAATTTTACAACTTATGATAATGTGCTACTTAGTAAAGGATTTGTCGAAACACAAGAA
AAGCTATATAAAGATATACCATCATATAAAGCAAGAGTTTTGCTAGGTGAGTGGATAGCAAGCACTGATTCAATTTTTAC
ACAAATAAATATTACTGATGATTATGTATTTACTAGCCCGATAGCATATTTAGACCCAGCATTTAGTGTTGGCGGGGATA
ACACTGCATTATGTGTTATGGAGCGAGTTGATGATAAGTATTATGCTTTTGTATTTCAAGACCAAAGACCAGCTAATGAT
CCTTATATTATGAATATGGTAAAGACTGTTATAGAAAATTTCAATGTGCATACACTGTATTTAGAGGATAGAGATAATAC
AAAAGGTGCTGGTGGATTGACCCGTGAATACATCTTGCTAAGAAGTAATATAAGCCAATATTTTAGAATTGTTCCAGTTA
AGCCAAAGTCTAATAAATTTAGCAGAATAACAACGTTAATTACGCCGTTTACTTACAAAAAACTTTATATTACAAAGTAC
AGTAGTTCTTCTGTATTTAATGATATTTATTCGTATAAGGGGGATAGCAAAACCCATGATGATGCTCTTGATGCAATGTC
TGCAGCATATTTGATGTTGTCTTTAGGATATAGAGAGCGAAGTGTTCACTTTGGCAATCAAAGATTTTTGTAA

>cp32-9 CP001211.1:29115-30467 Borrelia burgdorferi ZS7 plasmid ZS7_cp32-9, complete sequence
GTGAACTTATATCAAACAAAACTTTTTACAACACTACAAAAGGAATACAAAAATAAATATGGAGTTGACATATCACAATT
TGTAAAGCTAGCAAATTCTTCAATTAATTTTGATAAGTTTGAAGAAAAACAGTTAACTTTAAAACAAAAAAATGTGATAA
AAAGTATTAAAAAGAATAATGAAAAAAAGATTATACTCAGCGGAGGCATAGCTAGTGGCAAAACGTATCTTGCATGTTAT
CTTTTTCTAAAAAGTTTAATTGAAAATAAAAAGTTATACTCTAGTGATACTAATAATTTCATTATTGGGAATTCACAACG
CTCAGTTGAAGTTAATGTTTTAGGACAATTTGAAAAGCTATGTAAACTTCTTAAAATTCCTTATATCCCAAGACATACAA
ATAATTCATATATTCTGATTGATTCACTACGTATTAATCTATATGGTGGAGATAAGGCAAGTGATTTTGAAAGATTTAGG
GGAAGTAATTCAGCACTTATTTTTGTGAATGAGGCTACAACTTTACACAAGCAAACTTTAGAGGAGGTCTTAAAAAGACT
AAGATGCGGGCAAGAAACTATTATTTTTGATACTAATCCTGATCATCCAGAACACTATTTTAAAACCGATTATATTGATA
ATATAGCGACATTTAAGACATATAATTTTACAACTTATGATAATGTGCTACTTAGTAAAGGATTTATCGAAACACAAGAA
AAACTCTATAAAGATATACCATCATATAAAGCAAGAGTTTTGCTAGGTGAGTGGATAGCAAGCACTGATTCAATTTTTAC
ACAAATAAATATTACTGATGATTATATATTTACTAGCCCGATAGCATATTTAGACCCAGCATTTAGTGTTGGAGGGGATA
ACACTGCATTATGTGTTATGGAGCGAGTTGATGATAAGTATTATGCTTTTGTATTTCAAGACCAACGACCAGCCAATGAT
CCTTATATTATGAATATGGTAAAGACCGTTATAGAAAATTTCAATGTGCATACACTGTATTTAGAGGATAGAGATAATAC
AAAAGGTGCTGGTGGATTGACCCGCGAATACATCTTGCTAAGAAATAATATAAGCCAATATTTTAGAATTGTTCCAGTTA
AGCCAAAGTCTAATAAATTTAGCAGAATAACAACGTTAATTACGCCGTTTACTTACAAAAAACTTTATATTACAAAGTAC
AGTAGTTCTTCTGTATTTAATGATATTTATTCGTATAAGGGGGATAGCAAAACCCATGATGATGCTCTTGATGCAATATC
TGCAGCATATTTGATGTTGTCTTTAGGATATAGAGAGCGAAGTGTTCACTTTGGCAATCAAAGATTTTTGTAA

>cp32-13 CP001478.1:28736-30088 Borrelia burgdorferi CA-11.2a plasmid CA-11.2A
GTGAACTTATATCAAACAAAACTTTTTACAACACTACAAAAGGAATACAAAAATAAATATGGAGTTGACATATCACAATT
TGTAAAGCTAACAAATTCTTCAATTAATTTTGATAAGTTTGAAGAAAAACAGTTAACTTTAAAACAAAAAAATGTGATAA
AAAGCATTAAAAAGAATAATGAAAAAAAGATTATACTCAGCGGCGGCATAGCTAGCGGCAAAACGTATCTTGCATGCTAT
CTTTTTCTCAAAAGTTTAATTGAAAATAAAAAGTTATATTCTAGTGATACGAATAATTTTATTATTGGGAATTCACAACG
CTCAGTTGAAGTTAATGTTTTGGGACAATTTGAAAAGCTATGTAAACTTCTTAAAATTCCTTATATTCCAAGACATACAA
ATAATTCATATATTCTGATTGATTCACTACGTATTAATCTATATGGTGGAGATAAGGCAAGTGATTTTGAAAGATTTAGG
GGAAGTAATTCAGCACTTATTTTTGTTAATGAGGCTACAACTTTACACAAGCAAACTTTAGAGGAGGTCTTAAAAAGACT
AAGATGCGGGCAAGAAACTATTATTTTTGATACTAATCCTGATCATCCAGAACACTATTTTAAAACCGATTATATTGATA
ATATAGCGACATTTAAGACATATAATTTTACAACTTATGATAATGTTCTACTTAGTAAAGGATTTATCGAAACACAAGAA
AAACTCTATAAAGATATACCATCATATAAAGCAAGAGTTTTGCTAGGTGAGTGGATAGCAAGCACTGATTCAATTTTTAC
ACAAATAAATATTACTGATGATTATGTATTTACTAGTCCAATAGCATATTTAGACCCAGCATTTAGTGTTGGAGGAGATA
ACACTGCATTATGTGTTATGGAGCGAGTTGATGATAAGTATTATGCTTTTGTATTTCAAGACCAACGACCAGCCAATGAT
CCTTATATTATGAATATGGTAAAGACCGTTATAGAAAATTTCAATGTGCATACACTGTATTTAGAGGATAGAGATAATAC
AAAAGGTGCTGGTGGATTGACCCGCGAATACATCTTGCTAAGAAATAATATAAGCCAATATTTTAGAATTGTTCCAGTTA
AGCCAAAGTCTAATAAATTTAGCAGAATAACAACGTTAATTACGCCGTTTACTTATAAGAAACTTTACATTACAAAGTAC
AGTAGTTCTTCTGTATTTAATGATATTTATTCGTATAAGGGGGATAACAAAACCCATGATGATGCTCTTGATGCAATGTC
TGCAGCATATTTGATGTTGTCTTTAGGATATAGAGAGCGAAGTGTTCACTTTGGCAATCAAAGATTTTTGTAA

Alignment of phage terminase large subunit

cp32-12      GTGAACTTATATCAAACAAAACTTTTTACAACACTACAAAAGGAATACAAAAATAAATAT	60
cp32-9       GTGAACTTATATCAAACAAAACTTTTTACAACACTACAAAAGGAATACAAAAATAAATAT	60
cp32-13      GTGAACTTATATCAAACAAAACTTTTTACAACACTACAAAAGGAATACAAAAATAAATAT	60
cp32-2       GTGAACTTATATCAAACAAAACTTTTTACAACACTACAAAAGGAATACAAAAATAAATAT	60
cp32-10      GTGAACTTATATCAAACAAAACTTTTTACAACACTACAAAAGGAATACAAAAATAAATAT	60
cp32-11      GTGAACTTATATCAAACAAAACTTTTTACAACACTACAAAAGGAATACAAAAATAAATAT	60
cp32-8       GTGAACTTATATCAAACAAAACTTTTTACAACACTACAAAAGGAATACAAAAATAAATAT	60
cp32-1       GTGAACTTATATCAAACAAAACTTTTTACAACACTACAAAAGGAATACAAAAATAAATAT	60
cp32-5       GTGAACTTATATCAAACAAAACTTTTTACAACACTACAAAAGGAATACAAAAATAAATAT	60
cp32-6       GTGAACTTATATCAAACAAAACTTTTTACAACACTACAAAAGGAATATAAAAATAAATAT	60
cp32-3       GTGAACTTATATCAAACAAAACTTTTTACAACACTACAAAAGGAATATAAAAATAAATAT	60
cp32-4       GTGAACTTATATCAAACAAAACTTTTTACAACACTACAAAAGGAATATAAAAATAAATAT	60
cp32-7       GTGAACTTATATCAAACAAAACTTTTTACAACACTACAAAAGGAATATAAAAATAAATAT	60
             *********************************************** ************

cp32-12      GGAGTTGACATATCACAATTTGTAAAGCTAACAAATTCTTCAATTAATTTTGATAAGTTT	120
cp32-9       GGAGTTGACATATCACAATTTGTAAAGCTAGCAAATTCTTCAATTAATTTTGATAAGTTT	120
cp32-13      GGAGTTGACATATCACAATTTGTAAAGCTAACAAATTCTTCAATTAATTTTGATAAGTTT	120
cp32-2       GGAGTTGATATATCACAATTTGTAAAGCTAACAAATTCTTCAATTAATTTTGATAAGTTT	120
cp32-10      GGAGTTGATATATCACAATTTGTAAAGCTAACAAATTCTTCAATTAATTTTGATAAGTTT	120
cp32-11      GGAGTTGATATATCACAATTTGTAAAGCTAACAAATTCTTCAATTAATTTTGATAAGTTT	120
cp32-8       GGAGTTGATATATCACAATTTGTAAAGCTAACAAATTCTTCAATTAATTTTGATAAGTTT	120
cp32-1       GGAGTTGATATATCACAATTTGTAAAGCTAACAAATTCTTCAATTAATTTTGATAAGTTT	120
cp32-5       GGAGTTGATATATCACAATTTGTAAAGCTAACAAATTCTTCAATTAATTTTGATAAGTTT	120
cp32-6       GGAGTTGACATATCACAATTTGTAAAGCTAACAAATTCTTCAATTAATTTTGATAAGTTT	120
cp32-3       GGAGTTGACATATCACAATTTGTAAAGCTAACAAATTCTTCAATTAATTTTGATAAGTTT	120
cp32-4       GGAGTTGACATATCACAATTTGTAAAGCTAACAAATTCTTCAATTAATTTTGATAAGTTT	120
cp32-7       GGAGTTGACATATCACAATTTGTAAAGCTAACAAATTCTTCAATTAATTTTGATAAGTTT	120
             ******** ********************* *****************************

cp32-12      GAAGAAAAACAGTTAACTTTAAAACAAAAAAATGTGATAAAAAGCATTAAAAAGAATAAT	180
cp32-9       GAAGAAAAACAGTTAACTTTAAAACAAAAAAATGTGATAAAAAGTATTAAAAAGAATAAT	180
cp32-13      GAAGAAAAACAGTTAACTTTAAAACAAAAAAATGTGATAAAAAGCATTAAAAAGAATAAT	180
cp32-2       GAAGAAGAACAGTTAACTTTAAAACAGAAAAATGTGATAAAAAGCATTGAAAAGAATAAT	180
cp32-10      GAAGAAGAACAGTTAACTTTAAAACAAAAAAATGTGATAAAAAGCATTAAAAAGAATAAT	180
cp32-11      GAAGAAGAACAGTTAACTTTAAAACAAAAAAATGTGATAAAAAGCATTAAAAAGAATAAT	180
cp32-8       GAAGAAGAACAGTTAACTTTAAAACAAAAAAATGTGATAAAAAGCATTAAAAAGAATAAT	180
cp32-1       GAAGAAGAACAGTTAACTTTAAAACAAAAAAATGTGATAAAAAGCATTAAAAAGAATAAT	180
cp32-5       GAAGAAGAACAGTTAACTTTAAAACAAAAAAATGTGATAAAAAGCATTAAAAAGAATAAT	180
cp32-6       GAAGAAAAACAGTTAACTTTAAAACAAAAAAATGTGATAAAAAGCATTCAAAAGAATAAT	180
cp32-3       GAAGAAAAACAGTTAACTTTAAAACAAAAAAATGTGATAAAAAGCATTAAAAAGAATAAT	180
cp32-4       GAAGAAAAACAGTTAACTTTAAAACAAAAAAATGTGATAAAAAGCATTCAAAAGAATAAT	180
cp32-7       GAAGAAAAACAGTTAACTTTAAAACAAAAAAATGTGATAAAAAGCATTCAAAAGAATAAT	180
             ****** ******************* ***************** *** ***********

cp32-12      GAAAAAAAGATTATACTCAGCGGCGGCATAGCTAGCGGCAAAACGTATCTTGCCTGTTAT	240
cp32-9       GAAAAAAAGATTATACTCAGCGGAGGCATAGCTAGTGGCAAAACGTATCTTGCATGTTAT	240
cp32-13      GAAAAAAAGATTATACTCAGCGGCGGCATAGCTAGCGGCAAAACGTATCTTGCATGCTAT	240
cp32-2       GAAAAGAAGATTATACTCAGTGGAGGTATAGCTAGCGGCAAAACGTATCTTGCATGTTAT	240
cp32-10      GAAAAAAAGATTATACTCAGCGGCGGCATAGCTAGCGGCAAAACGTATCTTGCATGTTAT	240
cp32-11      GAAAAGAAGATTATACTCAGTGGAGGTATAGCTAGTGGCAAAACGTATCTTGCATGTTAT	240
cp32-8       GAAAAGAAGATTATACTCAGCGGAGGCATAGCTAGTGGCAAAACGTATCTTGCATGTTAT	240
cp32-1       GAAAAGAAGATTATACTCAGCGGAGGCATAGCTAGTGGCAAAACGTATCTTGCATGTTAT	240
cp32-5       GAAAAGAAGATTATACTCAGCGGAGGCATAGCTAGTGGCAAAACGTATCTTGCATGTTAT	240
cp32-6       GAAAAGAAGATTATACTCAGCGGAGGCATAGCTAGTGGCAAAACGTATCTTGCATGTTAT	240
cp32-3       GAAAAGAAGATTATACTCAGCGGAGGCATAGCTAGTGGCAAAACGTATCTTGCATGTTAT	240
cp32-4       GAAAAGAAGATTATACTCAGCGGAGGCATAGCTAGTGGCAAAACGTATCTTGCATGTTAT	240
cp32-7       GAAAAGAAGATTATACTCAGCGGAGGCATAGCTAGTGGCAAAACGTATCTTGCATGTTAT	240
             ***** ************** ** ** ******** ***************** ** ***

cp32-12      CTTTTTCTAAAAAGTTTAATTGCAAATAAGAATTTATATTCTAGTGATACGAATAATTTC	300
cp32-9       CTTTTTCTAAAAAGTTTAATTGAAAATAAAAAGTTATACTCTAGTGATACTAATAATTTC	300
cp32-13      CTTTTTCTCAAAAGTTTAATTGAAAATAAAAAGTTATATTCTAGTGATACGAATAATTTT	300
cp32-2       CTTTTTCTAAAAAGTTTAATTGAAAATAAAAAGCTATATTCTAGCGATACGAATAATTTC	300
cp32-10      CTTTTTCTAAAAAGTTTAATTAAAAATAAAAAGTTATATTCTAGCGATACGAATAATTTT	300
cp32-11      CTTTTTCTAAAAAGTTTAATTAAAAATAAAAAGTTATACTCTAGTGATACTAATAATTTC	300
cp32-8       CTTTTTCTAAAAAGTTTAATTGAAAATAAAAAGTTATACTCTAGTGATACTAATAATTTC	300
cp32-1       CTTTTTCTAAAAAGTTTAATTGAAATTAAAAAGTTATACTCTAGTGATACTAATAATTTC	300
cp32-5       CTTTTTCTAAAAAGTTTAATTGAAAATAAAAAGTTATACTCTAGTGATACTAATAATTTC	300
cp32-6       CTTTTTCTAAAAAGTTTAATTGAAAATAAAAAGTTATACTCTAGTGATACTAATAATTTC	300
cp32-3       CTTTTTCTAAAAAGTTTAATTGAAAATAAAAAGTTATACTCTAGTGATACTAATAATTTT	300
cp32-4       CTTTTTCTAAAAAGTTTAATTGAAAATAAAAAGTTATACTCTAGTGATACTAATAATTTT	300
cp32-7       CTTTTTCTAAAAAGTTTAATTGAAAATAAAAAGTTATACTCTAGTGATACTAATAATTTT	300
             ******** ************  ** *** **  **** ***** ***** ********

cp32-12      ATTATTGGCAATTCGCAGCGCTCGGTTGAAGTTAATGTTTTGGGGCAATTTGAAAAGCTA	360
cp32-9       ATTATTGGGAATTCACAACGCTCAGTTGAAGTTAATGTTTTAGGACAATTTGAAAAGCTA	360
cp32-13      ATTATTGGGAATTCACAACGCTCAGTTGAAGTTAATGTTTTGGGACAATTTGAAAAGCTA	360
cp32-2       ATTATAGGGAATTCACAACGTTCAGTTGAAGTTAATGTTTTGGGACAATTTGAAAAGCTA	360
cp32-10      ATTATTGGGAATTCACAACGCTCAGTTGAAGTTAATGTTTTGGGACAATTTGAAAAGCTA	360
cp32-11      ATTATAGGGAATTCACAACGTTCAGTTGAAGTTAATGTTTTGGGGCAATTTGAAAAGCTA	360
cp32-8       ATTATAGGGAATTCACAACGTTCAGTTGAAGTTAATGTTTTGGGGCAATTTGAAAAGCTA	360
cp32-1       ATTATAGGGAATTCACAACGTTCAGTTGAAGTTAATGTTTTGGGGCAATTTGAAAAGCTA	360
cp32-5       ATTATAGGGAATTCACAACGTTCAGTTGAAGTTAATGTTTTGGGGCAATTTGAAAAGCTA	360
cp32-6       ATTATAGGGAATTCACAACGTTCAGTTGAAGTTAATGTTTTGGGACAATTTGAAAAGCTA	360
cp32-3       ATTATTGGGAATTCACAACGCTCAGTTGAAGTTAATGTTTTGGGACAATTTGAAAAGCTA	360
cp32-4       ATTATTGGGAATTCACAACGCTCAGTTGAAGTTAATGTTTTGGGACAATTTGAAAAGCTA	360
cp32-7       ATTATTGGGAATTCACAACGCTCAGTTGAAGTTAATGTTTTGGGACAATTTGAAAAGCTA	360
             ***** ** ***** ** ** ** ***************** ** ***************

cp32-12      TGCAAACGGCTTAAAATTCCTTATATTCCAAGACATACAAATAATTCATATATTCTGATT	420
cp32-9       TGTAAACTTCTTAAAATTCCTTATATCCCAAGACATACAAATAATTCATATATTCTGATT	420
cp32-13      TGTAAACTTCTTAAAATTCCTTATATTCCAAGACATACAAATAATTCATATATTCTGATT	420
cp32-2       TGTAAACTTCTTAAAATTCCTTATATTCCAAGACATACAAATAATTCATATATTCTGATT	420
cp32-10      TGTAAACTTCTTAAAATTCCTTATATTCCAAGACATACAAATAATTCATATATTCTTATT	420
cp32-11      TGTAAACTTCTTAAAATTCCTTATATTCCAAGACATACAAATAATTCATATATTCTGATT	420
cp32-8       TGTAAACTTCTTAAAATTCCTTATATTCCAAGACATACAAATAATTCATATATTCTGATT	420
cp32-1       TGTAAACTTCTTAAAATTCCTTATATTCCAAGACATACAAATAATTCATATATTCTGATT	420
cp32-5       TGTAAACTTCTTAAAATTCCTTATATTCCAAGACATACAAATAATTCATATATTCTGATT	420
cp32-6       TGTAAACTTCTTAAAATTCCTTATATCCCAAGACATACAAATAATTCATATATTCTGATT	420
cp32-3       TGTAAACTTCTTAAAATTCCTTATATCCCAAGACATACAAATAATTCATATATTCTGATT	420
cp32-4       TGTAAACTTCTTAAAATTCCTTATATCCCAAGACATACAAATAATTCATATATTCTGATT	420
cp32-7       TGTAAACTTCTTAAAATTCCTTATATCCCAAGACATACAAATAATTCATATATTCTGATT	420
             ** ****  ***************** ***************************** ***

cp32-12      GATTCACTACGTATTAATCTATATGGTGGAGATAAGGCAAGTGATTTTGAAAGATTTAGG	480
cp32-9       GATTCACTACGTATTAATCTATATGGTGGAGATAAGGCAAGTGATTTTGAAAGATTTAGG	480
cp32-13      GATTCACTACGTATTAATCTATATGGTGGAGATAAGGCAAGTGATTTTGAAAGATTTAGG	480
cp32-2       GATTCACTGCGAATTAATCTATATGGTGGAGATAAGGCAAGTGTTTTTGAAAGATTTAGG	480
cp32-10      GATTCACTGAGAATTAATCTATATGGAGGAGATAAGGCAAGTGATTTTGAAAGATTTAGG	480
cp32-11      GATTCACTGCGAATTAATCTATATGGTGGAGATAAGGCAAGTGATTTTGAAAGATTTAGG	480
cp32-8       GATTCACTACGTATTAATCTATATGGAGGAGATAAGGCAAGTGATTTTGAAAGATTTAGG	480
cp32-1       GATTCACTACGTATTAATCTATATGGAGGAGATAAGGCAAGTGATTTTGAAAGATTTAGG	480
cp32-5       GATTCACTACGTATTAATCTATATGGAGGAGATAAGGCAAGTGATTTTGAAAGATTTAGG	480
cp32-6       GATTCACTACGTATTAATCTATATGGTGGAGATAAGGCAAGTGATTTTGAAAGATTTAGG	480
cp32-3       GATTCACTACGTATTAATCTATATGGTGGAGATAAGGCAAGTGATTTTGAAAGATTTAGG	480
cp32-4       GATTCACTACGTATTAATCTATATGGTGGAGATAAGGCAAGTGATTTTGAAAGATTTAGG	480
cp32-7       GATTCACTACGTATTAATCTATATGGTGGAGATAAGGCAAGTGATTTTGAAAGATTTAGG	480
             ********  * ************** **************** ****************

cp32-12      GGAAGTAATTCAGCGCTTATTTTTGTTAATGAGGCTACAACTTTACACAGGCAAACTTTA	540
cp32-9       GGAAGTAATTCAGCACTTATTTTTGTGAATGAGGCTACAACTTTACACAAGCAAACTTTA	540
cp32-13      GGAAGTAATTCAGCACTTATTTTTGTTAATGAGGCTACAACTTTACACAAGCAAACTTTA	540
cp32-2       GGAAGTAATTCAGCACTTATTTTTGTGAATGAGGCTACAACTTTACACAAGCAAACTTTA	540
cp32-10      GGAAGTAATTCGGCACTTATTTTTGTTAATGAGGCTACAACTTTACACAAGCAAACTTTA	540
cp32-11      GGAAGTAATTCAGCACTTATTTTTGTTAATGAGGCTACAACTTTACACAAGCAAACTTTA	540
cp32-8       GGAAGTAATTCGGCACTTATTTTTGTTAATGAGGCTACAACTTTACACAAGCAAACTTTA	540
cp32-1       GGAAGTAATTCGGCACTTATTTTTGTTAATGAGGCTACAACTTTACACAAGCAAACTTTA	540
cp32-5       GGAAGTAATTCGGCACTTATTTTTGTTAATGAGGCTACAACTTTACACAAGCAAACTTTA	540
cp32-6       GGAAGTAATTCGGCACTTATTTTTGTTAATGAGGCTACAACTTTACACAAGCAAACTTTA	540
cp32-3       GGAAGTAATTCGGCACTTATTTTTGTTAATGAGGCTACAACTTTACACAAGCAAACTTTA	540
cp32-4       GGAAGTAATTCGGCACTTATTTTTGTTAATGAGGCTACAACTTTACACAAGCAAACTTTA	540
cp32-7       GGAAGTAATTCGGCACTTATTTTTGTTAATGAGGCTACAACTTTACACAAGCAAACTTTA	540
             *********** ** *********** ********************** **********

cp32-12      GAGGAAGTCTTAAAAAGACTAAGGTGTGGGCAAGAAACTATTATTTTTGATACTAACCCC	600
cp32-9       GAGGAGGTCTTAAAAAGACTAAGATGCGGGCAAGAAACTATTATTTTTGATACTAATCCT	600
cp32-13      GAGGAGGTCTTAAAAAGACTAAGATGCGGGCAAGAAACTATTATTTTTGATACTAATCCT	600
cp32-2       GAGGAGGTCTTAAAAAGACTAAGATGCGGGCAAGAAACTATTATTTTTGATACTAATCCT	600
cp32-10      GAGGAGGTCTTAAAAAGACTAAGATGCGGGCAAGAAACTATTATTTTTGATACTAATCCC	600
cp32-11      GAGGAGGTCTTAAAAAGACTAAGATGCGGGCAAGAAACTATTATTTTTGATACTAATCCC	600
cp32-8       GAGGAAGTCTTAAAAAGACTAAGATGCGGGCAAGAAACTATTATTTTTGATACTAATCCT	600
cp32-1       GAGGAAGTCTTAAAAAGACTAAGATGCGGGCAAGAAACTATTATTTTTGATACTAATCCT	600
cp32-5       GAGGAAGTCTTAAAAAGACTAAGATGCGGGCAAGAAACTATTATTTTTGATACTAATCCT	600
cp32-6       GAGGAAGTCTTAAAAAGACTAAGATGCGGGCAAGAAACTATTATTTTTGATACTAATCCT	600
cp32-3       GAGGAGGTCTTAAAAAGACTAAGATGCGGGCAAGAAACTATTATTTTTGATACTAATCCT	600
cp32-4       GAGGAAGTCTTAAAAAGACTAAGATGCGGGCAAGAAACTATTATTTTTGATACTAATCCT	600
cp32-7       GAGGAAGTCTTAAAAAGACTAAGATGCGGGCAAGAAACTATTATTTTTGATACTAATCCT	600
             ***** ***************** ** ***************************** **

cp32-12      GATCATCCGGAACACTATTTTAAAACCGATTATATTGATAATATAGCAACTTTTAAGACA	660
cp32-9       GATCATCCAGAACACTATTTTAAAACCGATTATATTGATAATATAGCGACATTTAAGACA	660
cp32-13      GATCATCCAGAACACTATTTTAAAACCGATTATATTGATAATATAGCGACATTTAAGACA	660
cp32-2       GATCATCCAGAACACTATTTTAAAACCGATTATATTGATAATATAGCGACCTTTAAGACA	660
cp32-10      GATCATCCAGAACACTATTTTAAAACCGATTATATTGATAATATAGCGACATTTAAGACA	660
cp32-11      GATCATCCAGAACACTATTTTAAAACCGATTATATTGATAATATAGCGACCTTTAAGACA	660
cp32-8       GATCATCCAGAACACTATTTTAAAACCGATTATATTGATAATATAGCGACCTTTAAGACA	660
cp32-1       GATCATCCAGAACACTATTTTAAAACCGATTATATTGATAATATAGCGACCTTTAAGACA	660
cp32-5       GATCATCCAGAACACTATTTTAAAACCGATTATATTGATAATATAGCGACCTTTAAGACA	660
cp32-6       GATCATCCAGAACACTATTTTAAAACCGATTATATTGATAATATAGCGACCTTTAAGACA	660
cp32-3       GATCATCCAGAACACTATTTTAAAACCGATTATATTGATAATATAGCGACCTTTAAGACA	660
cp32-4       GATCATCCAGAACACTATTTTAAAACCGATTATATTGATAATATAGCGACCTTTAAGACA	660
cp32-7       GATCATCCAGAACACTATTTTAAAACCGATTATATTGATAATATAGCGACCTTTAAGACA	660
             ******** ************************************** ** *********

cp32-12      TATAATTTTACAACTTATGATAATGTTCTACTTAGTAAAGGATTTATCGAAACACAAGAA	720
cp32-9       TATAATTTTACAACTTATGATAATGTGCTACTTAGTAAAGGATTTATCGAAACACAAGAA	720
cp32-13      TATAATTTTACAACTTATGATAATGTTCTACTTAGTAAAGGATTTATCGAAACACAAGAA	720
cp32-2       TATAATTTCACAACTTATGATAATGTGCTACTTAGTAAAGGATTTGTCGAAACACAAGAA	720
cp32-10      TATAATTTTACAACTTATGATAATGTGCTACTTAGTAAAGGATTTGTCGAAACACAAGAA	720
cp32-11      TATAATTTTACAACTTATGATAATGTGCTACTTAGTAAAGGATTTGTCGAAACACAAGAA	720
cp32-8       TATAAGTTTACAACTTATGATAATGTGCTACTTAGTAAAGGATTTGTCGAAACACAAGAA	720
cp32-1       TATAAGTTTACAACTTATGATAATGTGCTACTTAGTAAAGGATTTGTCGAAACACAAGAA	720
cp32-5       TATAAGTTTACAACTTATGATAATGTGCTACTTAGTAAAGGATTTGTCGAAACACAAGAA	720
cp32-6       TATAATTTTACAACTTATGATAATGTGCTACTTAGTAAAGGATTTGTCGAAACACAAGAA	720
cp32-3       TATAATTTTACAACTTATGATAATGTGCTACTTAGTAAAGGATTTGTCGAAACACAAGAA	720
cp32-4       TATAAGTTTACAACTTATGATAATGTGCTACTTAGTAAAGGATTTGTCGAAACACAAGAA	720
cp32-7       TATAATTTTACAACTTATGATAATGTGCTACTTAGTAAAGGATTTGTCGAAACACAAGAA	720
             ***** ** ***************** ****************** **************


cp32-12      AAACTCTATAAAGATATACCATCATATAAAGCAAGAGTTTTGCTAGGTGAGTGGATAGCA	780
cp32-9       AAACTCTATAAAGATATACCATCATATAAAGCAAGAGTTTTGCTAGGTGAGTGGATAGCA	780
cp32-13      AAACTCTATAAAGATATACCATCATATAAAGCAAGAGTTTTGCTAGGTGAGTGGATAGCA	780
cp32-2       AAGCTATATAAAGATATACCATCATATAAAGCAAGAGTTTTGCTAGGTGAATGGATAGCA	780
cp32-10      AAGCTATATAAAGATATACCATCATATAAAGCAAGAGTTTTGCTAGGTGAGTGGATAGCA	780
cp32-11      AAGCTATATAAAGATATACCATCATATAAAGCAAGAGTTTTGCTAGGTGAGTGGATAGCA	780
cp32-8       AAGCTATATAAAGATATACCATCATATAAAGCAAGAGTTTTGTTAGGTGAGTGGATAGCA	780
cp32-1       AAGCTATATAAAGATATACCATCATATAAAGCAAGAGTTTTGTTAGGTGAGTGGATAGCA	780
cp32-5       AAGCTATATAAAGATATACCATCATATAAAGCAAGAGTTTTGTTAGGTGAGTGGATAGCA	780
cp32-6       AAGCTATATAAAGATATACCATCATATAAAGCAAGAGTTTTGTTAGGTGAGTGGATAGCA	780
cp32-3       AAGCTATATAAAGATATACCATCATATAAAGCAAGAGTTTTGTTAGGTGAGTGGATAGCA	780
cp32-4       AAGCTATATAAAGATATACCATCATATAAAGCAAGAGTTTTGTTAGGTGAGTGGATAGCA	780
cp32-7       AAGCTATATAAAGATATACCATCATATAAAGCAAGAGTTTTGTTAGGTGAGTGGATAGCA	780
             ** ** ************************************ ******* *********

cp32-12      AGCACTGATTCAATTTTTACACAAATAAATATTACTGATGATTATGTATTTACTAGCCCG	840
cp32-9       AGCACTGATTCAATTTTTACACAAATAAATATTACTGATGATTATATATTTACTAGCCCG	840
cp32-13      AGCACTGATTCAATTTTTACACAAATAAATATTACTGATGATTATGTATTTACTAGTCCA	840
cp32-2       AGCACCGATTCAATTTTTACACAAATAAATATTACTGATGATTATGTATTTACTAGCCCG	840
cp32-10      AGCACTGATTCAATTTTTACACAAATAAATATTACTGATGATTATGTATTTACTAGCCCG	840
cp32-11      AGCACTGATTCAATTTTTACACAAATAAATATTACTGATGATTATGTATTTACTAGCCCG	840
cp32-8       AGCACTGATTCAATTTTTACACAAATAAATATTACTGATGATTATGTATTTACTAGCCCG	840
cp32-1       AGCACTGATTCAATTTTTACACAAATAAATATTACTGATGATTATGTATTTACTAGCCCG	840
cp32-5       AGCACTGATTCAATTTTTACACAAATAAATATTACTGATGATTATGTATTTACTAGCCCG	840
cp32-6       AGCACTGATTCAATTTTTACACAAATAAATATTACTGATGATTATGTATTTACTAGCCCA	840
cp32-3       AGCACCGATTCAATTTTTACACAAATAAATATTACTGATGATTATGTATTTACTAGCCCG	840
cp32-4       AGCACTGATTCAATTTTTACACAAATAAATATTACTGATGATTATGTATTTACTAGCCCG	840
cp32-7       AGCACTGATTCAATTTTTACACAAATAAATATTACTGATGATTATGTATTTACTAGCCCG	840
             ***** *************************************** ********** **

cp32-12      ATAGCATATTTAGACCCAGCATTTAGTGTTGGCGGGGATAACACTGCATTATGTGTTATG	900
cp32-9       ATAGCATATTTAGACCCAGCATTTAGTGTTGGAGGGGATAACACTGCATTATGTGTTATG	900
cp32-13      ATAGCATATTTAGACCCAGCATTTAGTGTTGGAGGAGATAACACTGCATTATGTGTTATG	900
cp32-2       ATAGCATATTTAGACCCAGCATTTAGTGTTGGAGGGGATAACACTGCATTATGTGTTATG	900
cp32-10      ATAGCATATTTAGACCCAGCATTTAGTGTTGGCGGGGATAACACTGCATTATGTGTTATG	900
cp32-11      ATAGCATATTTAGACCCAGCATTTAGTGTTGGCGGGGATAACACTGCATTATGTGTTATG	900
cp32-8       ATAGCATATTTAGACCCAGCATTTAGTGTTGGAGGGGATAACACTGCATTATGTGTTATG	900
cp32-1       ATAGCATATTTAGACCCAGCATTTAGTGTTGGCGGGGATAACACTGCATTATGTGTTATG	900
cp32-5       ATAGCATATTTAGACCCAGCATTTAGTGTTGGCGGGGATAACACTGCATTATGTGTTATG	900
cp32-6       ATAGCATATTTAGACCCAGCATTTAGTGTTGGAGGAGATAACACTGCATTATGTGTTATG	900
cp32-3       ATAGCATATTTAGACCCAGCATTTAGTGTTGGCGGGGATAACACTGCATTATGTGTTATG	900
cp32-4       ATAGCATATTTAGACCCAGCATTTAGTGTTGGCGGGGATAACACTGCATTATGTGTTATG	900
cp32-7       ATAGCATATTTAGACCCAGCATTTAGTGTTGGCGGGGATAACACTGCATTATGTGTTATG	900
             ******************************** ** ************************

cp32-12      GAGCGAGTTGATGATAAGTATTATGCTTTTGTATTTCAAGACCAACGACCAGCTAATGAT	960
cp32-9       GAGCGAGTTGATGATAAGTATTATGCTTTTGTATTTCAAGACCAACGACCAGCCAATGAT	960
cp32-13      GAGCGAGTTGATGATAAGTATTATGCTTTTGTATTTCAAGACCAACGACCAGCCAATGAT	960
cp32-2       GAGCGAATTGATGATAAGTATTATGCTTTTGTATTTCAAGACCAACGACCAGCCAATGAT	960
cp32-10      GAGCGAGTTGATGATAAGTATTATGCTTTTGTATTTCAAGACCAAAGACCAGCTAATGAT	960
cp32-11      GAGCGAGTTGATGATAAGTATTATGCTTTTGTATTTCAAGACCAAAGACCAGCTAATGAT	960
cp32-8       GAGCGAGTTGATGATAAGTATTATGCTTTTGTATTTCAAGACCAAAGACCAGCTAATGAT	960
cp32-1       GAGCGAGTTGATGATAAGTATTATGCTTTTGTATTTCAAGACCAAAGACCAGCTAATGAT	960
cp32-5       GAGCGAGTTGATGATAAGTATTATGCTTTTGTATTTCAAGACCAAAGACCAGCTAATGAT	960
cp32-6       GAGCGAGTTGATGATAAGTATTATGCTTTTGTATTTCAAGACCAACGACCAGCCAATGAT	960
cp32-3       GAGCGAGTTGATGATAAGTATTATGCTTTTGTATTTCAAGACCAAAGACCAGCTAATGAT	960
cp32-4       GAGCGAGTTGATGATAAGTATTATGCTTTTGTATTTCAAGACCAAAGACCAGCTAATGAT	960
cp32-7       GAGCGAGTTGATGATAAGTATTATGCTTTTGTATTTCAAGACCAAAGACCAGCTAATGAT	960
             ****** ************************************** ******* ******
```

```
cp32-12      CCTTATATTATGAATATGGTAAAGACCGTTATAGAAAATTTCAATGTGCATACACTGTAT	1020
cp32-9       CCTTATATTATGAATATGGTAAAGACCGTTATAGAAAATTTCAATGTGCATACACTGTAT	1020
cp32-13      CCTTATATTATGAATATGGTAAAGACCGTTATAGAAAATTTCAATGTGCATACACTGTAT	1020
cp32-2       CCTTATATTATGAATATGGTAAAGACCGTTATAGAAAATTTCAATGTGCATACACTGTAT	1020
cp32-10      CCTTATATTATGAATATGGTAAAGACTGTTATAGAAAATTTCAATGTGCATACACTGTAT	1020
cp32-11      CCTTATATTATGAATATGGTAAAGACCGTTATAGAAAATTTCAATGTGCATACACTGTAT	1020
cp32-8       CCTTATATTATGAATATGGTAAAGACTGTTATAGAAAATTTCAATGTGCATACACTGTAT	1020
cp32-1       CCTTATATTATGAATATGGTAAAGACTGTTATAGAAAATTTCAATGTGCATACACTGTAT	1020
cp32-5       CCTTATATTATGAATATGGTAAAGACTGTTATAGAAAATTTCAATGTGCATACACTGTAT	1020
cp32-6       CCTTATATTATGAATATCGTAAAGACTGTTATAGAAAATTTCAATGTGCATACACTGTAT	1020
cp32-3       CCTTATATTATGAATATGGTAAAGACTGTTATAGAAAATTTCAATGTGCATACACTGTAT	1020
cp32-4       CCTTATATTATGAATATGGTAAAGACTGTTATAGAAAATTTCAATGTGCATACACTGTAT	1020
cp32-7       CCTTATATTATGAATATGGTAAAGACTGTTATAGAAAATTTCAATGTGCATACACTGTAT	1020
             ***************** ******** *********************************

cp32-12      TTAGAGGATAGAGATAATACAAAAGGTGCTGGTGGATTGACCCGTGAATACATCTTGCTA	1080
cp32-9       TTAGAGGATAGAGATAATACAAAAGGTGCTGGTGGATTGACCCGCGAATACATCTTGCTA	1080
cp32-13      TTAGAGGATAGAGATAATACAAAAGGTGCTGGTGGATTGACCCGCGAATACATCTTGCTA	1080
cp32-2       TTAGAGGATAGAGATAATACAAAAGGTGCTGGTGGATTGACCCGTGAATACATCTTGCTA	1080
cp32-10      TTAGAGGATAGAGATAATACAAAAGGTGCTGGTGGATTGACCCGTGAATACATCTTGCTA	1080
cp32-11      TTAGAGGATAGAGATAATACAAAAGGTGCTGGTGGATTGACCCGTGAATACATCTTGCTA	1080
cp32-8       TTAGAGGATAGAGATAATACAAAAGGTGCTGGTGGATTGACCCGTGAATACATCTTGCTA	1080
cp32-1       TTAGAGGATAGAGATAATACAAAAGGTGCTGGTGGATTGACCCGTGAATACATCTTGCTA	1080
cp32-5       TTAGAGGATAGAGATAATACAAAAGGTGCTGGTGGATTGACCCGTGAATACATCTTGCTA	1080
cp32-6       TTAGAGGATAGAGATAATACAAAAGGTGCTGGTGGATTGACCCGTGAATACATCTTACTA	1080
cp32-3       TTAGAGGATAGAGATAATACAAAAGGTGCTGGTGGATTGACCCGTGAATACATCTTGCTA	1080
cp32-4       TTAGAGGATAGAGATAATACAAAAGGTGCTGGTGGATTGACCCGTGAATACATCTTGCTA	1080
cp32-7       TTAGAGGATAGAGATAATACAAAAGGTGCTGGTGGATTGACCCGTGAATACATCTTGCTA	1080
             ******************************************** *********** ***

cp32-12      AGAAATAATATAAGCCAATATTTTAGAATTGTTCCAGTTAAGCCAAAGTCTAATAAATTT	1140
cp32-9       AGAAATAATATAAGCCAATATTTTAGAATTGTTCCAGTTAAGCCAAAGTCTAATAAATTT	1140
cp32-13      AGAAATAATATAAGCCAATATTTTAGAATTGTTCCAGTTAAGCCAAAGTCTAATAAATTT	1140
cp32-2       AGAAATAATACGAGTCAATATTTTAGAATTGTTCCAGTTAAGCCAAAGTCTAATAAATTT	1140
cp32-10      AGAAGTAATATAAGCCAATATTTTAGAATTGTTCCAGTTAAGCCAAAGTCTAATAAATTT	1140
cp32-11      AGAAGTAATATAAGCCAATATTTTAGAATTGTTCCAGTTAAGCCAAAGTCTAATAAATTT	1140
cp32-8       AGAAGTAATATAAGCCAATATTTTAGAATTGTTCCAGTTAAGCCAAAGTCTAATAAATTT	1140
cp32-1       AGAAGTAATATAAGCCAATATTTTAGAATTGTTCCAGTTAAGCCAAAGTCTAATAAATTT	1140
cp32-5       AGAAGTAATATAAGCCAATATTTTAGAATTGTTCCAGTTAAGCCAAAGTCTAATAAATTT	1140
cp32-6       AGAAGTAATATAAGCCAATATTTTAGAATTGTTCCAGTTAAGCCAAAGTCTAATAAATTT	1140
cp32-3       AGAAGTAATATAAGCCAATATTTTAGAATTGTTCCAGTTAAGCCAAAGTCTAATAAATTT	1140
cp32-4       AGAAGTAATATAAGCCAATATTTTAGAATTGTTCCAGTTAAGCCAAAGTCTAATAAATTT	1140
cp32-7       AGAAGTAATATAAGCCAATATTTTAGAATTGTTCCAGTTAAGCCAAAGTCTAATAAATTT	1140
             **** *****  ** *********************************************

cp32-12      AGCAGAATAACAACGTTAATTACGCCGTTTACTTATAAGAAACTTTACATTACAAAGTAC	1200
cp32-9       AGCAGAATAACAACGTTAATTACGCCGTTTACTTACAAAAAACTTTATATTACAAAGTAC	1200
cp32-13      AGCAGAATAACAACGTTAATTACGCCGTTTACTTATAAGAAACTTTACATTACAAAGTAC	1200
cp32-2       AGCAGAATAACAACGTTAATTACGCCGTTTACTTATAAGAAACTTTACATTACAAAGTAC	1200
cp32-10      AGCAGAATAACAACGTTAATTACGCCGTTTACTTACAAAAAACTTTATATTACAAAGTAC	1200
cp32-11      AGCAGAATAACAACGTTAATTACGCCGTTTACTTACAAAAAACTTTATATTACAAAGTAC	1200
cp32-8       AGCAGAATAACAACGTTAATTACGCCGTTTACTTACAAAAAACTTTATATTACAAAGTAC	1200
cp32-1       AGCAGAATAACAACGTTAATTACGCCGTTTACTTACAAAAAACTTTATATTACAAAGTAC	1200
cp32-5       AGCAGAATAACAACGTTAATTACGCCGTTTACTTACAAAAAACTTTATATTACAAAGTAC	1200
cp32-6       AGCAGAATAACAACGTTAATTACGCCGTTTACTTACAAAAAACTTTACATTACAAAGTAC	1200
cp32-3       AGCAGAATAACAACGTTAATTACGCCGTTTACTTACAAAAAACTTTATATTACAAAGTAC	1200
cp32-4       AGCAGAATAACAACGTTAATTACGCCGTTTACTTACAAAAAACTTTATATTACAAAGTAC	1200
cp32-7       AGCAGAATAACAACGTTAATTACGCCGTTTACTTACAAAAAACTTTATATTACAAAGTAC	1200
             *********************************** ** ******** ************

cp32-12      AGTAGTTCTTCTGTATTTAATGATATTTATTCGTATAAGGGGGATAACAAAACCCATGAT	1260
cp32-9       AGTAGTTCTTCTGTATTTAATGATATTTATTCGTATAAGGGGGATAGCAAAACCCATGAT	1260
cp32-13      AGTAGTTCTTCTGTATTTAATGATATTTATTCGTATAAGGGGGATAACAAAACCCATGAT	1260
cp32-2       AGTAGTTCTTCTGTATTTAATGATATTTATTCGTATAAGGGGGATAACAAAACCCATGAT	1260
cp32-10      AGTAGTTCTTCTGTATTTAATGATATTTATTCGTATAAGGGGGATAGCAAAACCCATGAT	1260
cp32-11      AGTAGTTCTTCTGTATTTAATGATATTTATTCGTATAAGGGGGATAGCAAAACCCATGAT	1260
cp32-8       AGTAGTTCTTCCGTATTTAATGATATTTATTCGTATAAGGGGGATAATAAAACCCATGAT	1260
cp32-1       AGTAGTTCTTCCGTATTTAATGATATTTATTCGTATAAGGGGGATAATAAAACCCATGAT	1260
cp32-5       AGTAGTTCTTCCGTATTTAATGATATTTATTCGTATAAGGGGGATAATAAAACCCATGAT	1260
cp32-6       AGTAGTTCTTCTGTATTTAATGATATTTATTCGTATAAGGGGGATAATAAAACCCATGAT	1260
cp32-3       AGTAGTTCTTCCGTATTTAATGATATTTATTCGTATAAGGGGGATAATAAAACCCATGAT	1260
cp32-4       AGTAGTTCTTCTGTATTTAATGATATTTATTCGTATAAGGGGGATAGCAAAACCCATGAT	1260
cp32-7       AGTAGTTCTTCTGTATTTAATGATATTTATTCGTATAAGGGGGATAGCAAAACCCATGAT	1260
             *********** **********************************  ************

cp32-12      GATGCTCTTGATGCAATGTCTGCAGCATATTTGATGTTGTCTTTAGGATATAGAGAGCGA	1320
cp32-9       GATGCTCTTGATGCAATATCTGCAGCATATTTGATGTTGTCTTTAGGATATAGAGAGCGA	1320
cp32-13      GATGCTCTTGATGCAATGTCTGCAGCATATTTGATGTTGTCTTTAGGATATAGAGAGCGA	1320
cp32-2       GATGCTCTTGATGCAATGTCTGCAGCATATTTGATGTTGTCTTTAGGATATAGAGAGCGA	1320
cp32-10      GATGCTCTTGATGCAATGTCTGCAGCATATTTGATGTTGTCTTTAGGATATAGAGAGCGA	1320
cp32-11      GATGCTCTTGATGCAATGTCTGCAGCATATTTGATGTTGTCTTTAGGATATAGAGAGCGA	1320
cp32-8       GACGCTCTTGATGCAATATCTGCAGCATATTTGATGTTGTCTTTAGGATATAGAGAGCGA	1320
cp32-1       GACGCTCTTGATGCAATATCTGCAGCATATTTGATGTTGTCTTTAGGATATAGAGAGCGA	1320
cp32-5       GACGCTCTTGATGCAATATCTGCAGCATATTTGATGTTGTCTTTAGGATATAGAGAGCGA	1320
cp32-6       GATGCTCTTGATGCAATGTCTGCAGCATATTTGATGTTGTCTTTAGGATATAGAGAGCGA	1320
cp32-3       GATGCTCTTGATGCAATGTCTGCAGCATATTTGATGTTGTCTTTAGGATATAGAGAGCGA	1320
cp32-4       GATGCTCTTGATGCAATGTCTGCAGCATATTTGATGTTGTCTTTAGGATATAGAGAGCGA	1320
cp32-7       GATGCTCTTGATGCAATGTCTGCAGCATATTTGATGTTGTCTTTAGGATATAGAGAGCGA	1320
             ** ************** ******************************************

cp32-12      AGTGTTCACTTTGGCAACCAAAGATTTTTGTAA	1353
cp32-9       AGTGTTCACTTTGGCAATCAAAGATTTTTGTAA	1353
cp32-13      AGTGTTCACTTTGGCAATCAAAGATTTTTGTAA	1353
cp32-2       AGTGTTCACTTTGGCAATCAAAGATTTTTGTAA	1353
cp32-10      AGTGTTCACTTTGGCAATCAAAGATTTTTGTAA	1353
cp32-11      AGTGTTCACTTTGGCAATCAAAGATTTTTGTAA	1353
cp32-8       AGTGTTCACTTTGGCAATCAAAGATTTTTGTAA	1353
cp32-1       AGTGTTCACTTTGGCAATCAAAGATTTTTGTAA	1353
cp32-5       AGTGTTCACTTTGGCAATCAAAGATTTTTGTAA	1353
cp32-6       AGTGTTCACTTTGGCAATCAAAGATTTTTGTAA	1353
cp32-3       AGTGTTCACTTTGGCAATCAAAGATTTTTGTAA	1353
cp32-4       AGTGTTCACTTTGGCAATCAAAGATTTTTGTAA	1353
cp32-7       AGTGTTCACTTTGGCAATCAAAGATTTTTGTAA	1353
             ***************** ***************
```

**Supplementary information 3**:

To ensure the specificity of the primer/probe set, BLAST analysis using sequences submitted to GenBank was performed. All hits with e-value < 0.01 were Lyme *Borrelia* species dominated by *B. burgdorferi* with multiple hits of the following *Borrelia* strains: *B. mayonii*, *B. garinii*, *B. afzelii*, *B. bisettii*, and *B. valasiana* (BLAST was carried out on the 5^th^ October 2020). According to the discovery of widespread presence of the *terL*-bearing cp32 plasmids, we predicted that the Ter-qPCR would show positive PCR results against Lyme *Borrelia* species except *B. spielmanii*. Indeed, ‘*In silico’* PCR (http://insilico.ehu.es/PCR/) revealed PCR product of the correct size from the plasmid fractions of *B. burgdorferi*, *B. mayonii*, *B. garinii*, *B. afzelii*, *B. bisettii*, *B. valasiana, B. bavariensis, B. finlandensis*. No PCR products were amplified from *B. spielmanii*. Positive PCR results were also obtained from all the LD *Borrelia* strains listed in Table 2, except *B. spielmanii*. No positive PCR results were ever observed from RF *Borrelia* strains including *B. miyamotoi* (Table 1) and other bacterial strains that have been used in the lab, including *Clostridium difficile*, *Clostridium perfringens*, *Escherichia coli*, *Pseudomonas aeruginosa*, *Streptococcus pneumoniae*, *Staphylococcus aureus*, *Burkholderia thailandensis*, *Burkholderia pseudomallei*, *Haemophilus influenzae* and *Salmonella enterica*.

To rule out the possibility of unspecific amplification with human DNA, UCSC In-Silico PCR (<https://genome.ucsc.edu/cgi-bin/hgPcr>) was conducted against human DNA. No PCR product was observed. In addition, commercial human female and male DNA and total DNA extracted from commercial HWB also didn’t display any positive Ter-qPCRs, while reference qPCR targeting human housekeeping genes RNase P (TaqMan™ Copy Number Reference Assay, Catalog number: 4403326) produced positive results. This confirmed that the Ter-qPCR did not amplify human DNA.
